# Loss of function and structural variants in the *PRKRA* synaptic gene link mild cognitive impairment and tinnitus

**DOI:** 10.1101/2025.11.04.25339446

**Authors:** Mai T. Pham, Alberto Bernal-Robledano, Alba Escalera-Balsera, Stefan Schoisswohl, Pablo Cruz-Granados, Paula Robles-Bolivar, Birgit Mazurek, Dimitrios Kikidis, Rilana Cima, Christopher R. Cederroth, Silvano Gallus, Myra Spiliopoulou, Berthold Langguth, Patricia Perez-Carpena, Winfried Schlee, Jose A. Lopez Escamez, the UNITI Consortium

## Abstract

The genetic basis underlying the co-occurrence of tinnitus and mild cognitive impairment (MCI) remains unknown. Adults with chronic tinnitus from a European cohort were classified into MCI (n=75) and non-MCI (n=201) groups, and rare variant distributions were assessed using gene burden analyses. Approximately one-fourth of individuals with chronic tinnitus exhibited MCI in their early fifties. Rare loss-of-function variants in *PRKRA* were identified in 18.6% of MCI patients, including a recurrent ∼3kb deletion in 14 individuals (OR=513.86, 95% CI [224.34-1177.02], adjusted *p*-value = 1.04E-48). These carriers reported tinnitus onset approximately eight years before MCI was observed. Cognitive impairment was linked to tinnitus, but not to hyperacusis or high-frequency hearing loss, among carriers of rare variants in MCI candidate genes, particularly *PRKRA.* The association between *PRKRA* variants and MCI was independent of *APOE* ε4 allele and known Parkinson’s disease dementia risk variants. In silico modelling and docking show increased binding affinity between mutant PACT (*PRKRA*) and PKR, suggesting over-activation of the PKR-eIF2α integrated stress response. Functional validation using a *PRKRA* knockout model identified differentially expressed genes enriched in dementia-related pathways. These findings support *PRKRA* as a potential driver gene in tinnitus-associated MCI.

Clinicaltrial.gov: NCT04663828

## Main

Tinnitus is the perception of sound without an external source, affecting about 10-30% of the population. For some, it becomes chronic and distressing condition (tinnitus disorder),^1^ and is often associated with high-frequency hearing loss (HFHL)^2^, hyperacusis^3^, and psychological comorbidities, including emotional disturbances such as anxiety and depression^4^.

Emerging evidence links tinnitus and hearing loss to cognitive impairment, particularly in older adults, including neurological conditions involving both auditory and non-auditory brain regions^5–7^. Tinnitus has been identified as an early sign of neurodegeneration in central nervous system (CNS), with changes in motor control, memory loss, seizures, and cognitive impairment in Alzheimer’s disease, Parkinson’s disease, and epilepsy^8^. Previous studies have shown the association between mild cognitive impairment (MCI), tinnitus distress, and HFHL^9^. Persistent tinnitus can impair emotional processing, attention, working memory and executive functions due to the chronic salience and distress associated with the perceived sound^10^. Genome-wide association studies have recently supported a distinct genetic architecture between hearing loss and tinnitus, with tinnitus sharing common genetic variants to neuropsychiatric disorders and personality traits supporting pleiotropy^11,12^.

Despite these clinical and neurobiological associations, the molecular mechanisms underlying tinnitus-related cognitive impairment remain poorly understood. A burden of rare missense variants in *ANK2, TCS2* and *AKAP9* have been reported in individuals with severe tinnitus^13^, but their impact on cognition remains unknown. One hypothesis is that specific genetic variants may show horizontal pleiotropy and predispose to both tinnitus and cognitive decline through independent neurobiological pathways. Conversely, genetic variants with vertical or mediated pleiotropy influencing hearing loss or tinnitus, could lead to cognitive impairment, either by sensory deprivation or depletion of cognitive resources. Understanding these genetic interactions could clarify why some tinnitus patients develop cognitive dysfunction while others do not.

Here, we performed rare variant analysis to identify candidate genes and biological pathways underlying the intersection between tinnitus and cognitive impairment. Using whole-genome sequencing data from 294 individuals with chronic tinnitus from the “Unification of Treatments and Interventions for Tinnitus Patients” (UNITI) trial^14^, we stratified participants according to cognitive performance on the Montreal Cognitive Assessment (MoCA) tool. We performed gene burden analysis (GBA) and structural variations in chronic tinnitus patients with MCI, to uncover genetic variants contributing to cognitive performance in tinnitus individuals. We further examined associations with tinnitus severity, hearing loss, hyperacusis, and depression, exploring the functional impact of candidate genes and its variants, providing a foundation for future mechanistic studies and therapeutic exploration in tinnitus-associated MCI.

## Materials and methods

### Participants

Data from adult individuals with chronic subjective tinnitus, who were recruited for the UNITI multicenter randomized clinical trial that compared single vs combination treatment in tinnitus (NCT04663828), were retrieved.^14^ Participants were recruited between April 2021 and December 2022 in five tertiary clinical European centers (Athens, Berlin, Granada, Leuven, and Regensburg). Assessment and treatment procedures were harmonized across centers.

### Ethical approval

The RCT was approved by local ethics committees of each site. The University of Sydney Human Ethics Committee approved the use of genomic de-identified data (Protocol #2023/199). Informed consent was obtained from all the participants prior to participation after clarifying the nature and possible consequences of the study.

### Cognitive and audiological assessment

Clinical and psychometric assessments including the Montreal Cognitive Assessment (MoCA), Tinnitus Handicap Inventory (THI), Hyperacusis questionnaire (GÜF), Patient Health Questionnaire for depressive symptoms (PHQ-9), and hearing thresholds (dB HL) at 500 Hz, 1, 2, 4, 8 kHz were obtained for 294 tinnitus patients in UNITI cohort^9^. MoCA scores (0-30) were used to classify chronic tinnitus patients with mild (18 – 25), moderate (10 – 17, strongly occur with Alzheimer’s disease), and severe (< 10) cognitive impairment^15^. THI, GÜF, and PHQ-9 were categorised into standard severity levels. THI assesses tinnitus-related distress through a questionnaire covering functional, emotional, and catastrophizing dimensions, with five severity levels: very mild (0–16), mild (18–36), moderate (38–56), severe (58 – 76), and catastrophic (78–100). The GÜF assessment consists of 15 questions with four levels handicap: slight (0 – 10), moderate (11–17), severe (18 – 25), and very severe (26 – 45)^16^. PHQ-9 is a self-administered test to assess psychological depressive symptoms, including non-depressive (1–4), mild-depressive (5–9), moderate-depressive (10–14), severe-depressive (15 – 27)^17^.

According to audiometric criteria of the UNITI-RCT protocol^18^, hearing loss (HL) was defined using pure-tone audiometry (PTA) as an average of 500 Hz, 1, and 2 kHz. Patients were classified as having unilateral HL if PTA exceeded 25 dB HL in one ear, or as bilateral HL if both ears were affected. High-frequency HL was defined as thresholds > 25 dB HL at 8 k Hz with a > 15 dB difference from PTA. High-frequency HL was measured as average thresholds at 4 kHz and 8 kHz for both ears.

### Whole genome sequencing

Whole genome sequencing was conducted on 294 tinnitus samples (from UNITI project), obtained from participants who provided additional consent for genetic studies. DNA was obtained from whole blood utilizing the QIAamp DNA Blood Mini Kit (Qiagen, Hilden, Germany), following the manufacturer protocol and quality control procedures, as previously described^19^. The library was sequenced on the Illumina platform, resulting paired-end sequences with a length of 150 bp and a minimum 30X coverage.

### Germline variant calling

Sequenced reads were mapped to the human reference genome GRCh38/hg38 using the Nextflow Sarek v2.7.1 nf-core workflow^20^, following GATK Best Practices for variant discovery^21^. The alignment was done with *BWA-MEM*^22^ and preprocessed following GATK (Genome Analysis Toolkit) best practices, including *picards MarkDuplicates* and *BaseRecalibrator* to correct for systematic biases by sequencing. *HaplotypeCaller* in GATK was then used for per-sample variant calling, followed by GATK *CombineGVCF/GenotypeGVCF* for joint calling with genotypic information for each sample. Variant filtering was then applied using GATK *VariantCalibrator* to obtain high-quality variant candidates with a sensitivity threshold of 90% for single nucleotide variants (SNVs) or small insertions/deletions (indels), based on known variant databases and VQSR model. Any variants marked as ‘PASS’ from VQSR filtering were subsequently filtered using hard-filter parameters, as recommended by GATK best practice to remove low-confident variants. Passed variants were additionally filtered by following gnomAD genotype filtering: Allele balance (AB) ≥ 0.2 and AB ≤ 0.8 (for heterozygous genotypes only), genotype quality (GQ) ≥ 20 and depth (DP) ≥ 10 (5 for haploid genotypes on sex chromosomes)^23^.

### Variant annotation and prioritization

Variant annotation was performed with the *Variant Effect Predictor* (*VEP*) v113^24^. Supplementary annotations included allele frequency (AF) of human genomes from gnomADg v4.1, reported for both non-Finnish European (NFE) and global populations, Combined Annotation Dependent Depletion (CADD) score, and pathogenic classifications based on the 2015 American College Medical Genetics/Association for Molecular Pathology (ACMG/AMP) guidelines^25^. Missense and Loss-of-Function (LoF) variants, including stop gain, splice donor/receptor, and start lost, were retained for downstream analysis. LOFTEE predictions were also used to retain high-confidence LoF variants. Splicing effects of the LoF variants, including splice donor or splice acceptor variants, were predicted with *spliceAI*^26^, using the GRCh38/hg38 GENCODE annotation and a maximum distance of 500 bp, and further validated by Human Splicing Finder Pro^27^. Variants with CADD scores < 20 or AF ≥ 0.05 in external control populations (gnomAD NFE/global) were excluded. Only genes containing at least one variant observed in more than one individual were retained for downstream analyses.

### Gene burden analysis and gene prioritization

Gene burden analysis (GBA) was conducted in individuals with MCI (18 ≤ MoCA ≤ 25) and different levels of tinnitus, hyperacusis, and depression symptoms. At gene level, aggregated allele frequencies of replicated variants were computed in the case cohort, compared with the NFE gnomAD population as an external control. GBA in MCI individuals were also conducted against the non-MCI group (MoCA > 25) as an internal control. Gene burden statistics including odd ratio (OR) at 95% confidence intervals, (CI), etiological fraction (FE), and *p*-values adjusted for multiple correction using the Bonferroni method were reported. Genes were considered statistically enriched if they exhibited adjusted *p*-value < 0.05 in one or more of the control datasets. Frequently mutated genes were excluded^28^.

We searched for target genes associated with brain diseases in SynaptomeDB v0.99.17^29^, which includes proteomic datasets of synaptic compartments (hereafter referred to synaptic genes) and human brain disease annotation based on Human Disease Ontology^30^, hereafter referred to brain-disease-associated genes (BD-associated genes). Alzheimer’s disease genes with consistently altered protein expression were also retrieved from consolidated Alzheimer’s disease protein change resources^31^.

### Structural variant analysis

*CNVkit* v0.9.8^32^, *Manta* v1.6.0^33^, and *TIDDIT* v2.3.1^34^ were employed to detect SVs. After per-sample variant calling for each caller, SVs were underwent a quality-filtering process using *SVDB* v2.8.1 (https://github.com/J35P312/SVDB), categorising those identified by at least two SV callers with ≥ 60% reciprocal overlap and those exclusively detected by either of the tools. SVs were subsequently annotated using *annotSV* v3.4.6 with default parameters^35^.

Rare SVs (AF < 0.05 in gnomAD-SV v4.1 NFE or global populations) that overlapped genes with significant LoF or missense SNV burdens were retained for downstream analyses. For SVs only detected by one tool, SVs with an exact match were kept for further analysis. For those detected by more than one tool, when an exact match for an SV was not available in the database, we used the allele frequency of the closest overlapping variant (e.g., ≥ 50% reciprocal overlap) as a proxy.

### Ancestry markers in target gene regions

We used the gnomAD NFE population to identify common variants as ancestry-informative markers within target gene regions. Variants with max ΔAF (the largest absolute AF difference observed across all non-NFE populations) ≥ 0.1 were selected as ancestry markers, representing common variants with potential frequency divergence between NFE and other populations. Selected variants were then used to stratify potential population ancestry of rare variants located in target gene regions in the MCI cohort relative to the non-MCI cohort and the gnomAD NFE population.

### APOE genotyping

To further assess the correlation between THI, GÜF, and MoCA scores in patients with late onset of Alzheimer’s disease, we categorized 294 tinnitus samples with *APOE* ε4 genotypes based on the two missense variants (rs429358 and rs4293). Samples were classified as non-*APOE* ε4 (ε2/ε2, ε3/ε3, and ε2/ε3) and *APOE* ε4 (ε2/ε4, ε3/ε4, and ε4/ε4).

### Protein modelling and molecular docking

The PBD file for wild-type protein were retrieved from the AlphaFold2 Protein Structure Database and assessed for quality^36^. For proteins not available in AlphaFold2, amino acid sequences were obtained from the UniProt Database^37^ and used to generate *ab initio* wild-type models using AlphaFold3^38^. The models were evaluated based on the predicted local distance difference test scores for α-helices and β-sheets, and the predicted template modelling score for the overall structure. Models with the highest quality scores were selected for further analysis. Mutant protein models were generated by homology modelling using *MODELLER* v10.6^39^. Amino acid sequences were retrieved from UniProt, and the resulting mutant models were evaluated using *MODELLER*’s built-in quality metrics, including the DOPE and GA341 scores. Protein visualisation was done in PyMOL Open Source (Schrodinger, LLC. 2010. The PyMOL Molecular Graphics System, Version 3.0.x.). *PRODIGY* v2.4 was also used to predict the binding affinity in protein-protein interaction^40^. For SNVs, *Dynamut2* was used to predict the impact of mutations on protein structural stability and to analyse atomic interactions^41^.

Molecular docking was performed using the ClusPro Server^42^. The wild-type and mutant protein models were docked using their respective PDB files. Ligand or receptor protein structures were obtained from the AlphaFold2 database.

### Gene expression in *PRKRA* knockout cell line model

Publicly available bulk RNA-seq FASTQ data from *PRKRA* knockout and control prostate cancer LNCaP cell lines were used^43^. Reads were aligned to the GRCh38 reference genome (GENCODE v48) using *STAR* v2.7.10b^44^, and gene-level read counts were quantified using *RSEM* v1.3.3^45^, following the ENCODE bulk RNA-seq processing pipeline. Differential gene expression analysis between knockout and control samples was performed using *edgeR* v4.8.2^46^ with *p*-values adjusted for multiple testing using the BH method, followed by pathway enrichment analysis using *clusterProfiler*.

### RNA and protein expression of target genes in human brain

RNA expression of target genes in human cortical brain regions, including entorhinal cortex (a part of hippocampal formation), frontal cortex, hippocampus, motor cortex (paracentral and precentral gyrus), sensory cortex (postcentral gyrus), and cingulate gyrus, was retrieved in the Human Protein Atlas^47^. Brain regions showing significantly altered protein expression in Alzheimer’s disease vs. normal brain tissues with false discovery rate (FDR) cutoff of 0.05 were obtained from consolidated Alzheimer’s disease protein change resources^31^.

### Statistical analysis and data visualisation

Protein-protein association networks and functional enrichment analysis were performed using STRING database v12^48^ and *clusterProfiler* v4.18.4^49^, respectively. The two-sided Wilcoxon rank-sum test was used to compare two independent groups without normality consumption, unless otherwise specified. Data visualisation and statistical plots were generated using R v4.5.1. Read alignment against GRCh38/hg38 was visualized using the Integrative Genome Visualisation (IGV) tool v17.0.9.^50^

## Results

### Participant characteristics

The study was analysed from 294 tinnitus patients across five clinical centres (UNITI cohort) with 39.5% females and average baseline THI score of 46.0 (IQR 30.5 to 62.0) at 51.6 ± 12.1 years old (Supplementary Table 1, Supplementary Fig. 1A). The MoCA scores of these participants ranged from 20 to 30, with 25.5% (75 individuals) scoring below 26 classified as MCI (Supplementary Fig. 1B). MCI participants were marginally older were slightly older and exhibited greater HFHL compared to those with the normal MoCA score (54.4 ± 11.0 vs. 50.6 ± 12.3, *p* = 0.02 (median age); 45.5 [27.5, 56.3] dB vs. 37.5 [20.9, 31.6] dB, *p* = 0.03 (HFHL); 19.2 (13.3, 28.3) dB vs. 15.8 (10.0, 24.2) dB, *p* = 0.005 (PTA)). No significant differences were observed in gender distribution, the proportions of tinnitus patients with unilateral or bilateral HL, tinnitus/hyperacusis-related hearing measures (THI/GÜF) or PHQ-9 score for depression.

Correlations between cognitive impairment and tinnitus/hyperacusis showed negative associations in the MCI subgroup, with higher significant correlations between GÜF and MoCA scores (*r* = -0.42, *p*_adj_ = 8.44E-04) than between THI and MoCA scores (*r* = -0.31, *p*_adj_ = 1.10E-02). PHQ-9 was found to be highly correlated with tinnitus (*r* > 0.68, *p*_adj_ < 2.2E-16) and hyperacusis distress (*r* > 0.48, *p*_adj_ < 2.2E-16), regardless of MoCA level. No correlation was observed between HL scores and MoCA, PHQ-9, or THI/GÜF in either MCI or non-MCI patients. Regarding *APOE* genotypes, 18.7% of MCI individuals were observed with *APOE* ε4, which was comparable to 27.9% of the non-MCI group (Fisher’s Exact test, *p* > 0.05).

### Burden of rare variants in the MCI subgroup

The GBA of high-confidence LoF variants identified 15 significant genes (hereafter referred to GBA genes) for the MCI subgroup vs. external controls, with 6 synaptic genes including *PRKRA* (*OR* = 49.81 [17.59–141.03], *p*_adj_ = 3.27E-11), *HK3* (*OR* = 109.06 [22.97–517.92], *p*_adj_ = 6.35E-07), *IGFN1* (*OR* = 54.50 [12.42–239.15], *p*_adj_ = 2.07E-05)*, PIK3C2G* (*OR* = 9.83 [4.03–23.97], *p*_adj_ = 8.91E-05), *GRAMD1A* (*OR* = 12.70 [4.64–34.77], *p*_adj_ = 1.35E-04), and *ENDOG* (*OR* = 10.96 [3.45–34.86], *p*_adj_ = 8.89E-03) (Table 1, Supplementary Table 2). Besides, regions including genes with a burden of rare variants showed similar allele frequencies in common variants selected as ancestry markers between tinnitus patients with and without MCI (*p* = 0.69) and the NFE population in gnomAD (*p* = 0.64) (Supplementary Table 3). These results support that the observed enrichment of rare LoF variants is more likely attributable to MCI-related genetic differences rather than difference population ancestry.

**Table 1.** Synaptic gene burden for rare high-confident loss-of-function variants (CADD ≥ 20) associated with mild cognitive impairment.

| Gene | Number of variants | Number of individuals | gnomADg (NFE)* |  |  | gnomADg (global)<sup>†</sup> |  |  |
| --- | --- | --- | --- | --- | --- | --- | --- | --- |
|  |  |  | OR (95% CI)<sup>‡</sup> | Adjusted-*P* | EF | OR (95% CI)<sup>‡</sup> | Adjusted-*P* | EF |
| *PRKRA* | 2 | 4 | 49.81<br>(17.59 - 141.03) | 3.27E-11 | 0.98 | 85.82<br>(30.67 – 240.15) | 3.98E-15 | 0.99 |
| *HK3* | 1 | 2 | 109.06<br>(22.97 - 517.92) | 6.35E-07 | 0.99 | 138.28<br>(31.15 – 613.79) | 1.60E-08 | 0.99 |
| *IGFN1* | 1 | 2 | 54.50<br>(12.42 - 239.15) | 2.07E-05 | 0.98 | 34.54<br>(8.35 – 142.85) | 1.79E-04 | 0.97 |
| *PIK3C2G* | 3 | 5 | 9.83<br>(4.03 - 23.97) | 8.91E-05 | 0.90 | 14.35<br>(5.90 – 34.87) | 7.31E-07 | 0.93 |
| *GRAMD1A* | 1 | 4 | 12.70<br>(4.64 - 34.77) | 1.35E-04 | 0.92 | 21.54<br>(7.89 – 58.79) | 3.65E-07 | 0.95 |
| *ENDOG* | 1 | 3 | 10.96<br>(3.45 - 34.86) | 8.89E-03 | 0.91 | 9.53<br>(3.02 – 30.06) | 2.12E-02 | 0.90 |

\* Non-Finnish European subpopulation in gnomAD genome v4.1.
<sup>†</sup> All populations in gnomAD genome v4.1.
<sup>‡</sup> Odds ratio with 95% confidence interval.
Abbreviations: NFE, Non-Finnish European; OR, Odds Ratio, CI, Confidence Interval; EF, Etiological Fraction.
*Note:* Genes are ordered by adjusted *P*-values (Bonferroni correction) of gnomADg (NFE).

Similarly, the GBA of missense variants found 37 significant GBA genes, with the 5 most significant synaptic genes in MCI vs. external controls (*OR* ≥ 10, *p*_adj_ < 0.01), including *EIF4G1*, *ERBB3*, *EP400*, *ANKRD52*, and *ATP6V1E2* (Table 2, Supplementary Table 4). No significant differences in allele frequencies of ancestry-informative markers were observed within genes with a burden of rare missense variants between the MCI and non-MCI tinnitus cohorts (*p* = 0.63), or MCI tinnitus cohort and gnomAD-NFE used as external controls (*p* = 0.73) (Supplementary Table 5).

**Table 2.** Synaptic gene burden for rare missense variants (CADD ≥ 20) associated with mild cognitive impairment (MCI).

| Gene | Number of variants | Number of individuals | gnomADg (NFE)* |  |  | gnomADg (global)<sup>†</sup> |  |  |
| --- | --- | --- | --- | --- | --- | --- | --- | --- |
|  |  |  | OR (95% CI)<sup>‡</sup> | Adjusted-*P* | EF | OR (95% CI)<sup>‡</sup> | Adjusted-*P* | EF |
| *EIF4G1* | 1 | 4 | 154.39<br>(48.60 - 490.47) | 3.74E-14 | 0.99 | 102.33<br>(36.01 - 290.75) | 1.09E-14 | 0.99 |
| *ERBB3* | 1 | 2 | 436.09<br>(61.02 - 3116.56) | 4.04E-06 | 1 | 32.23<br>(7.81 - 133.08) | 4.63E-03 | 0.97 |
| *ATP12A* | 3 | 14 | 4.48<br>(2.67 - 7.51) | 3.90E-05 | 0.78 | 4.08<br>(2.44 - 6.83) | 2.66E-04 | 0.75 |
| *EP400* | 1 | 2 | 852.91<br>(76.92 - 9457.48) | 1.12E-04 | 1 | 467.54<br>(84.98 - 2572.24) | 4.64E-09 | 1 |
| *PRX* | 1 | 2 | 62.30<br>(14.03 - 276.52) | 1.61E-04 | 0.98 | 0.65<br>(0.16 - 2.64) | *ns* | -0.53 |
| *KIAA1328* | 1 | 2 | 39.65<br>(9.24 - 170.14) | 2.14E-03 | 0.97 | 0.40<br>(0.10 - 1.63) | *ns* | -1.48 |
| *ANO2* | 1 | 2 | 37.93<br>(8.86 - 162.32) | 2.78E-03 | 0.97 | 3.12<br>(0.77 - 12.62) | *ns* | 0.68 |
| *ANKRD52* | 1 | 3 | 17.32<br>(5.40 - 55.53) | 4.69E-03 | 0.94 | 25.63<br>(8.05 - 81.58) | 1.64E-04 | 0.96 |
| *ATP6V1E2* | 1 | 4 | 11.17<br>(4.09 - 30.53) | 7.45E-03 | 0.91 | 13.79<br>(5.07 - 37.49) | 7.94E-04 | 0.93 |
| *SCN5A* | 1 | 3 | 16.25<br>(5.07 - 52.02) | 7.77E-03 | 0.94 | 0.66<br>(0.21 - 2.06) | *ns* | 0.53 |
| *REEP6* | 1 | 2 | 27.25<br>(6.47 - 114.76) | 1.93E-02 | 0.96 | 19.55<br>(4.78 - 80.00) | *ns* | 0.95 |
| *AGAP3* | 2 | 5 | 7.57<br>(3.10 - 18.45) | 2.52E-02 | 0.87 | 8.89<br>(3.65 - 21.58) | 4.19E-03 | 0.89 |
| *TBCC* | 1 | 3 | 13.29<br>(4.17 - 42.40) | 3.60E-02 | 0.92 | 17.19<br>(5.43 - 54.45) | 3.88E-03 | 0.94 |
| *TMED3* | 1 | 2 | 24.23<br>(5.78 - 101.55) | 3.80E-02 | 0.96 | 1.97<br>(0.49 - 7.96) | *ns* | 0.49 |
| *TNKS1BP1* | 2 | 12 | 3.60<br>(2.01 - 6.42) | 4.41E-02 | 0.72 | 4.37<br>(2.45 - 7.79) | 1.74E-03 | 0.77 |

\* Non-Finnish European subpopulation in gnomAD genome v4.1.
<sup>†</sup> All populations in gnomAD genome v4.1.
<sup>‡</sup> Odds ratio with 95% confidence interval.
Abbreviations: NFE, Non-Finnish European; OR, Odds Ratio, CI, Confidence Interval; EF, Etiological Fraction; *ns*, non-significant.
Note: Genes are ordered by adjusted *P*-values (Bonferroni correction) of gnomADg (NFE).

Of note, the burden of LoF variants *PRKRA* was not observed in the group of tinnitus individuals without MCI (Supplementary Figs. 2A, 3).

### Rare structural variants in the MCI subgroup

We found a total of 36,800 SVs in the MCI subgroup, of which approximately 7% were detected by more than one tool with at least 60% reciprocal overlap. After filtering, four high-quality SVs (including three intronic and one loss-of-function) overlapped genes previously identified as enriched for rare SNVs and showed a reciprocal overlap of at least 50% with rare SVs in the gnomAD SV v4.1 reference dataset (Supplementary Table 6). Notably, a loss-of-function deletion of ∼3 kb (2:178444501-178447508del) in *PRKRA* was found in 14 MCI individuals compared to external and internal controls (*OR* = 513.86 [224.34–1177.02], *p*_adj_ = 1.04E-48 (gnomAD-NFE); *OR* = 6.79 [2.56–18.03], *p*_adj_ = 3.58E-03 (non-MCI) (Supplementary Figs. 2B, 4-6). Common variants used as NFE ancestry markers close to the deletion region (+/-10kB) showed no differences in AF between the MCI cohort and reference controls (two-sided Wilcoxon test, *p* = 0.49 (non-MCI); *p* = 0.66 (gnomAD-NFE), Supplementary Table 3), suggesting no potential differences driven by genetic population ancestry in this region. Variant carriers were recruited from multiple centers, indicating no apparent site-specific bias (Supplementary Fig. 7). Notably, the age of tinnitus onset in these individuals differed significantly from the age at clinical trial inclusion, with a mean difference of 8 years (*p* = 8.1E-04). Other two intronic deletions, 2:46524048-46524172del and 11:57304632-57304828del, were found in synaptic genes *ATP6V1E2* and *TNKS1BP1*, respectively, both of which also exhibited an enrichment of missense variants.

### Genes associated with MCI and tinnitus/hyperacusis annoyance

We identified 52 GBA genes from the MCI subgroup, including 21 synaptic genes and 12 BD-associated synaptic genes, two of which showing significant downregulation at the protein level in Alzheimer’s disease vs. controls across multiple studies of bulk tissue (Figs. 1A-B, Supplementary Table 7).

**Figure 1.**
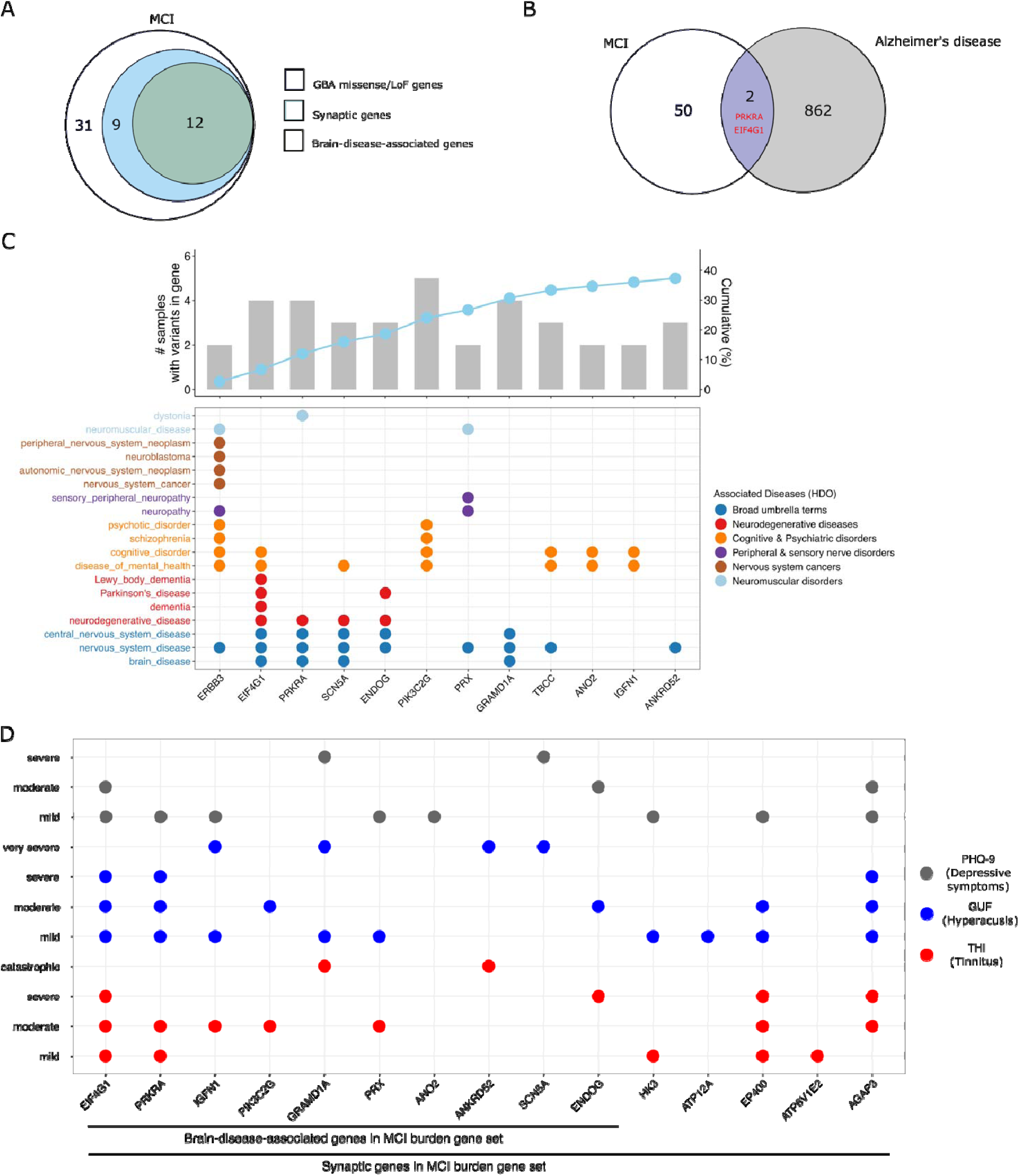
Genes significantly associated to tinnitus and mild cognitive impairment (MCI), severity levels of tinnitus, hyperacusis, depressive symptoms, and neurological diseases, based on gene burden analysis (GBA) and consolidated studies. **(A)** Venn diagram showing 52 GBA genes identified in the MCI subgroup, of which 21 were synaptic and 12 were associated with brain diseases. **(B)** Venn diagram showing the overlap between the MCI gene burden set and Alzheimer’s disease genes. Brain-disease-associated genes are highlighted in red. **(C)** Dot plot illustrating the distribution of 12 brain-disease-associated genes enriched in MCI (x-axis) across brain diseases (y-axis), based on Human Disease Ontology (HDO) and synaptomeDB. The accompanying bar plot shows cumulative proportion (%) of 75 MCI samples (y-axis) explained by variants in these genes (x-axis). Genes are ranked by their occurrence across brain diseases. **(D)** Dot plot showing enrichment of synaptic genes associated with MCI (x-axis) across severity levels of tinnitus, hyperacusis, and depressive symptoms (y-axis). MCI, mild cognitive impairment.

Figure 1C presents the brain disease annotations, according to HDO database, for the 12 BD-associated genes enriched in the MCI subgroup, variants of which were found in about 40% of the 75 MCI samples. Four of these genes, *EIF4G1*, *PRKRA*, *SCN5A*, and *ENDOG*, were strongly linked to neurodegenerative diseases. Notably, *EIF4G1* and *ENDOG* have been associated with dementia subtypes, including Parkinson’s disease and Lewy body dementia, while *PRKRA* is commonly linked to dystonia-parkinsonism. However, LoF variants in PRKRA among MCI individuals were not significantly associated with risk variants in *TMEM175*, a gene linked to Parkinson’s disease dementia (Supplementary Fig. 8). Besides *EIF4G1*, other MCI burden genes such as *ERBB3*, *PIK3C2G*, *TBCC*, *ANO2*, and *IGFN1* were associated with MCI. Functional enrichment analysis revealed a predominance of transmembrane channel-related pathways, particularly those involving potassium ion channels (Supplementary Fig. 9, Supplementary Table 8). No dementia-related gene networks were significantly enriched in the MCI gene burden set. Instead, *PRKRA* and *EIF4G1* were explicitly found in the protein association network of parkinsonism in UniProt (KW-0908) and a database of disease-gene associations (DOID: 14330) (Supplementary Fig. 10).

To assess the association between MCI and tinnitus, as well as its comorbidities, gene burden set of different severity levels of tinnitus, hyperacusis and depressive symptoms in the UNITI cohort was further overlapped with genes enriched in the MCI subgroup (Fig. 1D, Supplementary Tables 9-19). Approximately half of the BD-associated genes enriched in the MCI subgroup overlapped with genes associated with specific levels of tinnitus annoyance. *PRKRA*, *IGFN1*, *PIK3C2G* and *PRX* were associated with mild-to-moderate tinnitus, while *GRAMD1A*, *ANKRD52* and *ENDOG* were enriched in patients with severe-to-catastrophic tinnitus. The latter genes were also found to be associated with moderate-to-severe hyperacusis and/or depressive symptoms. Other synaptic genes, *HK3* and *ATP6V1E2*, were enriched in individuals with mild tinnitus, but not in severe tinnitus. In contrast, a broader set of non-synaptic gene (*NCKAP5*, *LGALS12*, *PSKH1*, *KCNK12*, *UPK2*, *CDHR4* and *PLEKHA3*) were exclusively found in patients with severe-to-catastrophic tinnitus annoyance (Supplementary Fig. 11).

### Association between cognitive performance, auditory phenotypes and *APOE* genotypes in MCI individuals

Although MoCA and THI scores were correlated in the MCI subgroup (*r^2^* = 0.09, *p* = 0.007; Fig. 2A), we observed stronger correlations in MCI individuals carrying rare missense and LoF variants in GBA genes (*r^2^* = 0.14, *p* = 0.002; Fig. 2B). Of these, BD-associated synaptic genes showed moderate correlations between MoCA and THI/GÜF scores (*r^2^* = 0.33, *p* = 0.001 (THI); *r^2^* = 0.31, *p* = 0.003 (GÜF); Fig. 2D), which were higher than those of synaptic genes without brain disease records (*r^2^* = 0.17, *p* = 0.002 (THI); *r^2^* = 0.28, *p* < 0.001 (GÜF); Fig. 2C).

**Figure 2.**
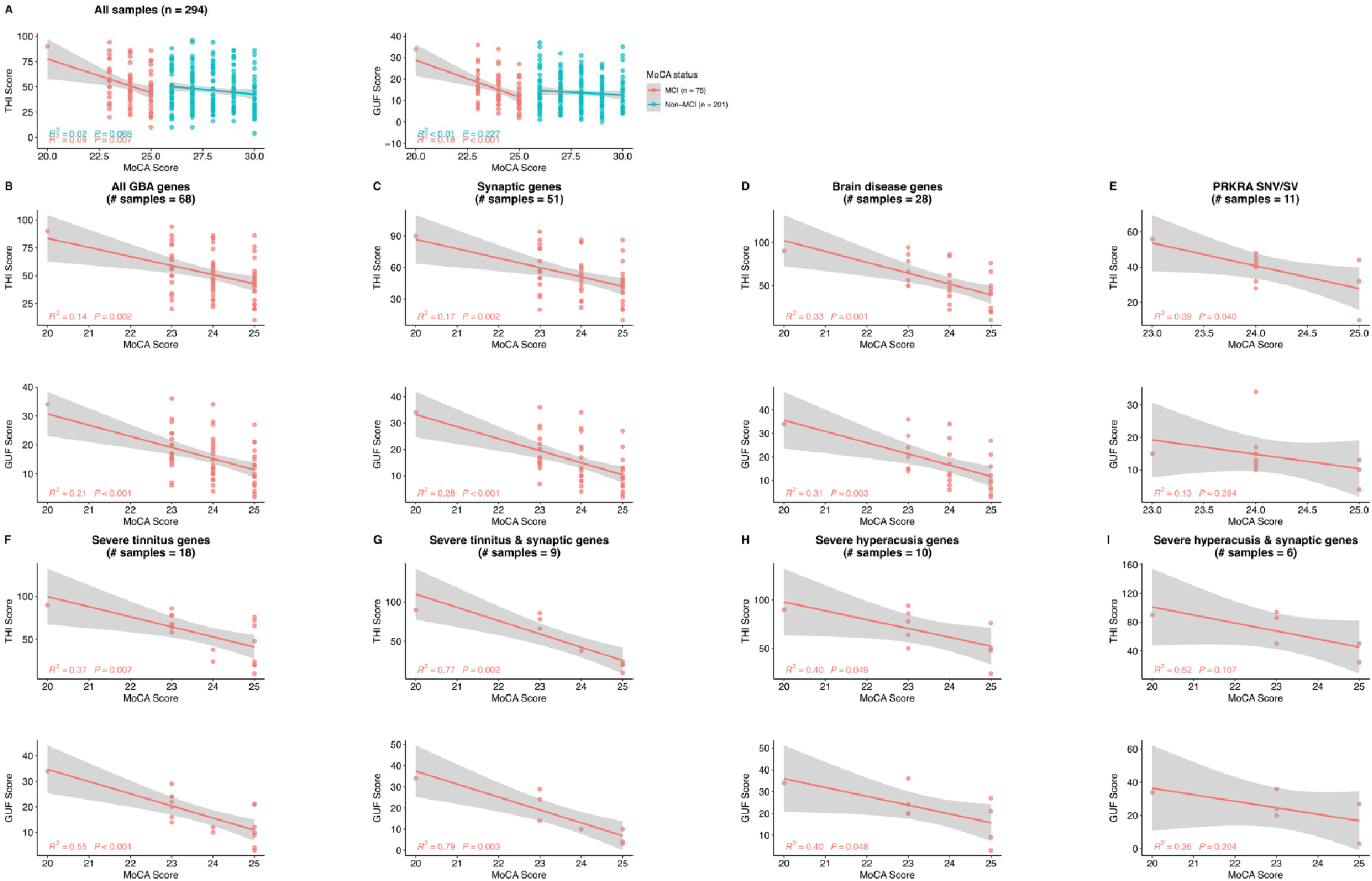
Correlation analyses between psychometric instruments Tinnitus Handicap inventory (THI), hyperacusis questionnaire (GÜF), and Montreal Cognitive Score (MoCA) score in the UNITI cohort. Scatterplot showing the correlation between MoCA score (x-axis) and THI/GÜF score (y-axis) of **(A)** 294 UNITI individuals (75 MCI and 201 non-MCI); **(B)** 68 MCI individuals carrying rare missense and loss-of-function variants in 52 burden genes; **(C)** 51 MCI individuals carrying rare missense and loss-of-function variants in 21 synaptic burden genes; **(D)** 28 MCI individuals carrying rare missense and loss-of-function variants in 12 burden genes associated with brain diseases; **(E)** 11 MCI individuals carrying rare missense and loss-of-function variants (including structural variants) in *PRKRA* gene, associated with MCI and mild-to-moderate tinnitus; **(F)** 20 MCI individuals carrying rare missense and loss-of-function variants in 3 synaptic genes (*GRAMD1A*, *ANKRD52*, *ENDOG*) and 8 non-synaptic genes (*NCKAP5*, *LGALS12*, *ERMARD*, *PSKH1*, *KCNK12*, *UPK2*, *CHDR4*, *PLEKHA3*) shared between MCI and severe/catastrophic tinnitus; **(G)** 9 MCI individuals carrying rare missense and loss-of-function variants in 3 synaptic genes (*GRAMD1A*, *ANKRD52*, *ENDOG*) shared between MCI and severe-to-catastrophic tinnitus; **(H)** 10 MCI individuals carrying rare missense and loss-of-function variants in 2 synaptic genes (*ANKRD52*, *SCN5A*) and 2 non-synaptic genes (*NCKAP5*, *RBM23*) shared between MCI and severe-to-very-severe hyperacusis; **(I)** 6 MCI individuals carrying rare missense and loss-of-function variants in 2 synaptic genes (*ANKRD52*, *SCN5A*) shared between MCI and severe-to-very-severe hyperacusis. Sample size for regression plots are indicated in each panel title. GBA, gene burden analysis; MCI, mild cognitive impairment.

We also examined hearing thresholds (in dB HL) at normal-to-high frequencies in relation to MoCA, THI, and GÜF scores (Supplementary Figs. 12-14). Similar to THI and GÜF measures, a moderate correlation was observed between HFHL at 4 and 8 kHz and MoCA score only in MCI patients with variants in BD-associated synaptic genes (*r^2^* = 0.19, *p* = 0.02; Supplementary Fig. 12D). Different from HFHL, no significant relationships were detected between MoCA and hearing thresholds at the conversational frequency range (500 Hz, 1 kHz, and 2 kHz) across any of the representative gene sets associated MCI, tinnitus, or hyperacusis. No significant association was found between HL and tinnitus/hyperacusis (Supplementary Figs. 13-14).

Notably, the *PRKRA* gene, which harbored two LoF variants and one SV in 11 MCI individuals with mild-to-moderate tinnitus, showed a strong association between MoCA and THI score (*r^2^* = 0.39, *p* = 0.04; Fig. 2E), but no correlation with hyperacusis (*r^2^* = 0.13, *p* > 0.05; Fig. 2E) or HFHL (*r^2^* < 0.01, *p* > 0.05; Supplementary Fig. 12E). In contrast, a remarkedly strong correlation between MoCA and THI/GÜF was observed in individuals carrying variants in genes shared between MCI and severe-to-catastrophic tinnitus, and this association was further strengthened for synaptic genes (Figs. 2F-G). Associations of THI vs. GÜF or PTA vs. HFHL scores were largely preserved across most gene sets (Supplementary Figs. 15-16). Strong overlaps in gene burden between tinnitus and hyperacusis in the same severity levels were also observed (Supplementary Fig. 17).

Given the established association of PHQ-9 for depression, cognitive impairment and hearing scores in the UNITI cohort^9^, we further examined these relationships in representative gene sets linked to these traits (Supplementary Figs. 18-19). Similarly to tinnitus or hyperacusis, a stronger correlation between PHQ-9 and MoCA scores was observed in BD-associated genes enriched in the MCI individuals (*r^2^* = 0.26, *p* = 0.01; Supplementary Fig. 18D) compared to non-BD genes (*r^2^* = 0.11, *p* = 0.03; Supplementary Fig. 18C). Its association with THI/GÜF score was substantially strong in synaptic genes associated with MCI and severe tinnitus (Supplementary Figs. 18F-G), but not PTA or HFHL scores (Supplementary Fig. 19).

We further assessed the distribution of MoCA and hearing scores in *APOE* ε4 carriers from the same gene sets but did not observe significant differences between individuals with and without *APOE* ε4 (Supplementary Figs. 20-21).

### Characterisation of *PRKRA* variants in the MCI subgroup

We identified two splice acceptor *PRKRA* variants located at the 5’ end of exon 7, resulting in PACT mutants with insertions at Lys-173 (K173) within the evolutionarily conserved dsRNA binding motif 2 (dsRBM2) (Fig. 3A). The variants were predicted to weaken the canonical acceptor site (score 0.73) 1 bp upstream and disrupt a donor site 95 bp upstream, without generating new cryptic donor or acceptor sites (Supplementary Tables 20-21). Additionally, we identified a ∼3 kB loss-of-function deletion in *PRKRA*, predicted to cause a frameshift after ILE-105 (I105) that locates in a non-functional domain between dsRBM2 and dsRBM3. Structural modelling of the mutant PACT protein confirmed premature translation termination, leading to the loss of N-terminal conserved domain 1/2, which likely disrupted the interaction between wild-type PACT and double-stranded-RNA-dependent protein kinase PKR (encoded by *EIF2AK2*) (Fig. 3B, Supplementary Fig. 22). The protein-kinase interaction between mutant PACT and PKR exhibited ∼380x tighter binding affinity and increased hydrogen/hydrophobic contacts (Supplementary Table 22).

**Figure 3.**
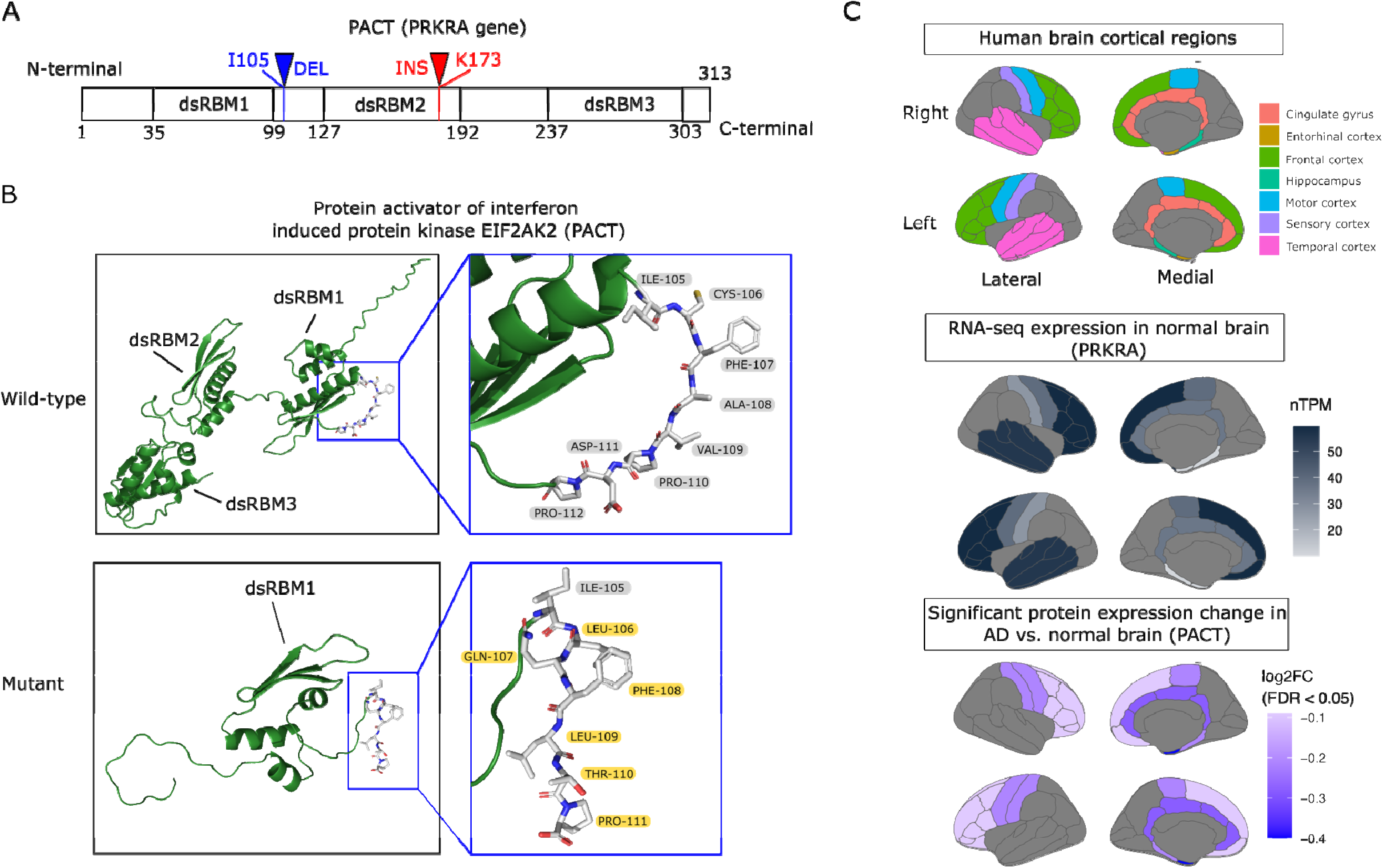
Protein models and protein expression of protein activator of interferon induced protein kinase EIF2AK2 (PACT) in human brain. **(A)** Locations of loss-of-function splice acceptor variants (red) and a 3,007-bp deletion (blue) along the PACT protein sequence; **(B)** Structural models of wild-type PACT and mutant PACT carrying the 3,007-bp deletion. Altered amino acids are marked in yellow; **(C)** RNA-seq expression of *PRKRA* across human cortical regions, measured in normalized transcript per million (nTPM), based on the Human Protein Atlas (upper and middle panels). Lower panel shows differential protein expression of PACT, measured in average log2-fold change (log_2_FC) in Alzheimer’s disease (AD) versus normal brain tissues (FDR < 0.05) ^31^.

### Expression of PACT (*PRKRA*) in the human brain

In human cortical brain regions, PACT mRNA was highly expressed in the frontal and temporal cortices. At the protein level, PACT expression was reduced in the entorhinal cortex, frontal cortex, and hippocampus in Alzheimer’s disease compared with normal brain tissues (Fig. 3C).

### Transcriptome profiling of *PRKRA* knockout cell lines

Approximately 7% of differentially expressed genes were synaptic genes associated with various dementia subtypes, including Alzheimer’s disease, frontotemporal dementia, vascular dementia, Lewy body dementia, and Parkinson’s disease dementia. The most significantly dysregulated genes, including *CLU*, *EEF1A2*, VIM, and RAB27A, have been linked to Alzheimer’s disease (Fig. 4A). Notably, we observed transcriptional modulation of established dementia risk genes, including *APP*, *PSEN1*, and *GBA1* in *PRKRA* knockout cells. In addition, severe tinnitus-associated genes such as *TSC2* were also down-regulated following *PRKRA* loss. Pathway enrichment analysis consistently identified dementia-related pathways among the top five over-represented KEGG pathways (Fig. 4B). Gene ontology biological processes were selectively enriched for the regulation of protein synthesis, unfolded protein response, stress granule assembly, and intrinsic cellular stress-sensing pathways (Fig. 4C).

**Figure 4.**
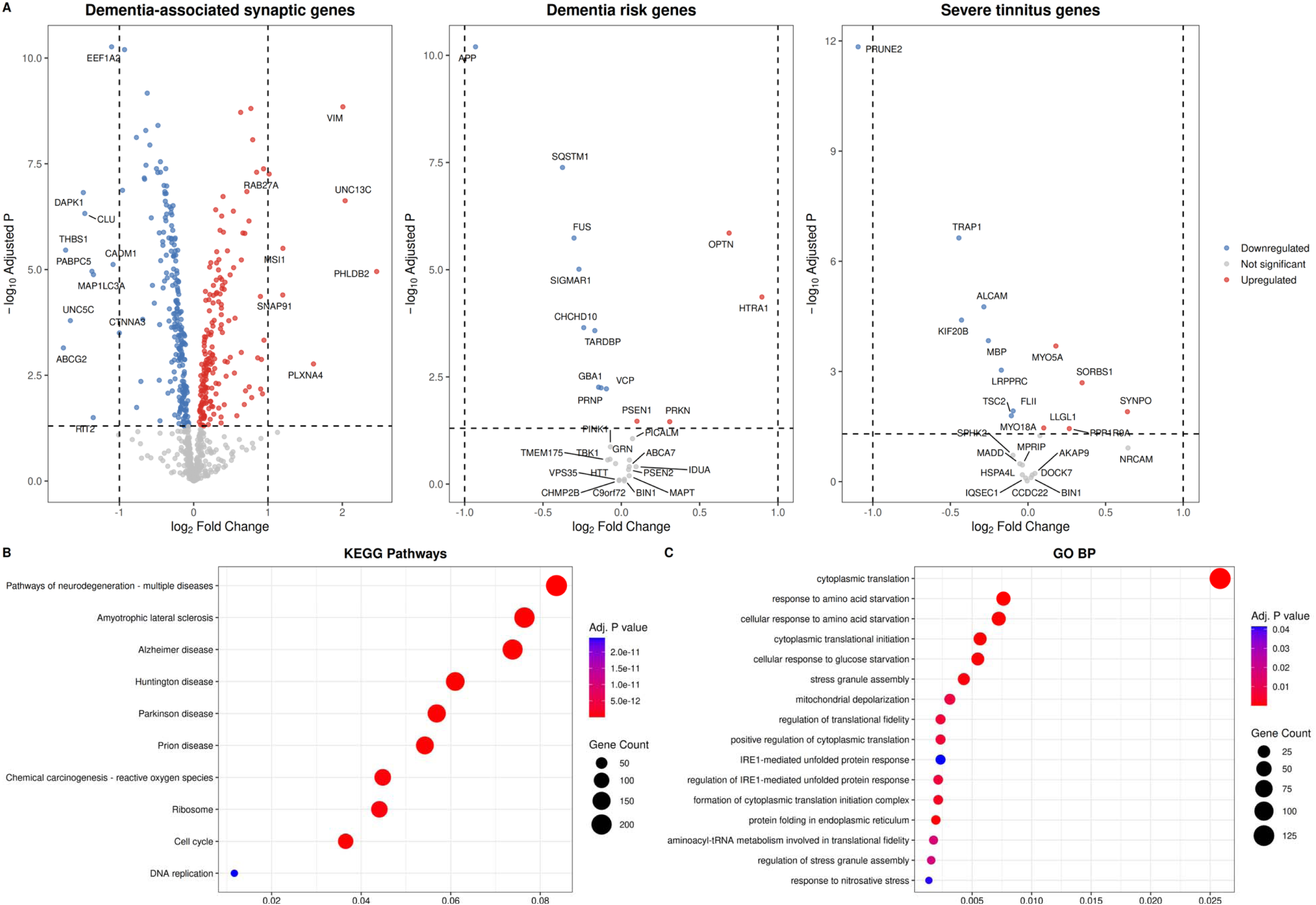
Differential gene expression and pathway enrichment analysis of *PRKRA* cellular knockout model. **(A)** Volcano plots showing differentially expressed genes (FDR-adjusted *p* < 0.05), including up-regulated (red) and down-regulated (blue) genes in dementia-associated synaptic genes^29^ with the most significant genes highlighted (left), dementia risk genes^66,67^ (middle), and severe tinnitus-related genes^13^ (right). The x-axis represents log2 fold change (*PRKRA* knockout vs. control), and y-axis represents -log10(FDR-adjusted *p*-value); **(B)** Dotplot showing the top 10 enriched Kyoto Encyclopedia of Genes and Genomes (KEGG) pathways and **(C)** Gene Ontology biological processes (GO-BP). The x-axis shows gene ratio, dot size represents the number of genes involved, and color intensity reflects FDR-adjusted *p*-values.

## Discussion

This study has found that about 25% of individuals with chronic tinnitus show MCI in their early fifties, and support that hearing testing and cognitive screening would be recommended in all patients with chronic tinnitus.

In the UNITI cohort, most patients with chronic tinnitus have a moderate to severe tinnitus, sound hypersensitivity, and HFHL at their age of fifties. MCI participants had higher ages and more severe HFHL compared to chronic tinnitus patients without MCI. Difference in sex and other hearing measurements were unnoticeable.

This study has identified 12 candidate genes that carry rare missense and LoF variants in MCI-associated tinnitus patients. These genes are involved in the synaptic proteome of human brain and have been implicated in neurodegeneration. The correlation between cognitive impairment (MoCA) and tinnitus distress (THI) scores was modest in groups of patients with or without MCI, but was notably stronger among the carriers of rare variants in MCI-associated synaptic genes, notably *PRKRA*, *EIF4G1*, and *ENDOG* genes involved in transcriptional and translational regulation and implemented in Parkinson’s disease dementia^51–53^. These genes have also been associated with hearing impairment phenotypes, including *PRKRA* associated with hearing loss, *ENDOG* associated with noise-induced hearing loss, or *ERBB2* with ultra rare structural variants found in individuals with severe tinnitus^54–56^. Of note, the 12 MCI candidate genes were not linked to *APOE* ε4 allele, which is the established genetic risk factor for late-onset Alzheimer’s disease.

Among top MCI candidate genes, *PRKRA* emerged as the most promising gene linked to tinnitus and MCI. We identified two loss-of-function short insertions and a structural deletion in *PRKRA* that link tinnitus with MCI. These associations were also independent of the *APOE* ε4 genotype. The *PRKRA* deletion was not observed in tinnitus individuals without MCI and the deletion carriers reported pre-existing tinnitus approximately eight years before MCI was observed during the UNITI recruitment. This supports that tinnitus may occur several years before MCI and that this deletion observed in individuals with tinnitus is likely to be a risk factor for early-onset dementia.

The *PRKRA* gene encodes PACT, a stress-responsive double-stranded RNA-binding protein that activates protein kinase R (PKR), which initiates integrated stress response (ISR) under cellular stress conditions. Our study suggests that *PRKRA* variants identified in MCI individuals may disrupt this regulatory mechanism. Structural modelling of a recurrent ∼3 kb deletion (observed in 14 MCI cases) predicts a truncated PACT protein with enhanced PKR binding, potentially resulting in aberrant PKR activation and sustained eIF2α phosphorylation. Prolonged activation of the PACT-PKR-eIF2α stress signaling pathway has been implicated in translational dysregulation, synaptic dysfunction, and neuronal death, suggesting a potential mechanism linking *PRKRA* variants to neurodegeneration^57,58^.

*PRKRA* mutations have been reported in unrelated European families with early-onset dystonia-parkinsonism^52,59^. Affected individuals present with muscular movement difficulties and, in some cases, later-onset mild parkinsonian features accompanied by cognitive deficits. On the other hand, *PRKRA* knockout animal models support a critical role of PACT (*PRKRA*) in ear development and auditory function^54^. Its loss leads to movement disorders with hearing abnormalities, including impaired hearing and abnormal ear morphology. In human, variants mapping to the DFNB27 locus (2q23-q31) overlap the *PRKRA* genomic region^60^, further implicating this locus in hearing-related phenotypes. Consistently, another mouse model carrying a *PRKRA* frameshift mutation shows progressive dystonia with cerebellar defects and impaired eIF2α phosphorylation via PACT-PKR-eIF2α stress response pathway^61^. In our study, deletion of PACT in human cell line models also suggests transcriptional dysregulation of multiple dementia-associated synaptic genes and risk genes, particularly APP and PSEN1, together with enrichment of neurodegenerative disorders and regulation of stress-related pathways.

In situ hybridization has found *Prkra* mRNA in the marginal layer of the stria vascularis in the mouse cochlea^54^. It is also highly expressed in the frontal and temporal cortices in human brain^47^. PACT protein expression is observed to be decreased in Alzheimer’s disease brain tissues, mostly in hippocampus, sensory, motor, and frontal cortices^31^. These regional and expression-specific patterns reinforce their potential involvement in vulnerability to neurodegeneration and hearing impairment. These findings highlight the dual role of PACT in hearing and cognitive function.

Collectively, these findings suggest that *PRKRA* variants in MCI-associated tinnitus patients may contribute to both tinnitus distress and cognitive vulnerability, potentially through shared dysregulation of the stress response pathways such as PKR-mediated ISR pathway implicated in Alzheimer’s disease and Parkinson’s disease^62–64^.

Our study also observed that UNITI patients may have tinnitus several years before developing MCI, which supports the hypothesis that identified *PRKRA* variants may exhibit vertical pleiotropy, where a single genetic alteration influences stress-response and synaptic vulnerability that underlie the auditory disorder and later contribute to cognitive impairment. *PRKRA*-related mechanisms may operate at a pre-degenerative or stress-adaptive stage, where translational control and synaptic homeostasis are disrupted, but not yet exhausted in mild tinnitus, possibly owing to compensatory pathways before neurodegeneration onset.

Our transcriptomic findings in the PRKRA knock-out cellular model show neurodegeneration as the top enriched pathway and support a causal molecular mechanism linking the dysfunction of auditory pathways to changes in gene expression involving frontal and temporal cortices overtime.

Further functional validation, including co-expression profiling, PKR-eIF2α signaling assays, and auditory phenotyping in model systems, are warranted to confirm the pathogenic relevance of *PRKRA* variants in the mechanism linking tinnitus to dementia manifestations. Replication in independent non-European cohorts is warranted to clarify whether these genetic mutations represent tinnitus-specific mechanisms or broader pathways shared with non-amyloidogenic forms of dementia.

Despite these limitations, our findings strengthen the proposed link between cognitive impairment and tinnitus, highlighting the involvement of ISR pathways mediated by PACT-PKR-eIF2α signaling cascade. These results support the rationale for further mechanistic studies and exploration of this pathway as a potential therapeutic target.

## Conclusions

The strong association of rare LoF structural variants in *PRKRA* with both tinnitus distress and cognitive impairment suggests a vertical pleiotropic effect, potentially mediated by genetic disruption in neuronal stress-response pathways. The *PRKRA* association is not observed with hyperacusis or HFHL, and it is independent of the *APOE* ε4 allele. Our results support a mechanistic link between chronic tinnitus and MCI, possibly through dysregulation of the PACT-PKR-eIF2α stress signaling cascade, which warrants further validations. The findings emphasize the central role of stress-related translational control in maintaining cognitive and auditory function and may open new avenues for therapeutic exploration targeting *PRKRA* as a potential genetic factor for dementia susceptibility.

## Data Availability

All data produced in the present study are available upon reasonable request to the authors.

## Contributors

MTP: conceptualisation, methodology, data curation, formal analysis, visualization, and writing, review and editing. PCG: methodology, formal analysis, review and editing. ABR, AEB, SS, PRB, BM, DK, RC, CRC, SG, MS, BL, PPC, WS: data collection, review and editing. JALE: conceptualisation, design, funding acquisition, project administration, supervision, writing, review and editing. All authors reviewed the final draft and approved the decision to submit for publication.

## Data availability

The data that support the findings of this study are openly available in ClinVar at https://www.ncbi.nlm.nih.gov/clinvar/, under the accession numbers SCV007579990 to SCV007579992. De-identified cohort-level VCF file are available upon request. Publicly available bulk RNA-sequencing FASTQ files from *PRKRA* knockout and control LNCaP cells were retrieved from the NCBI Sequence Read Archive (SRA) under accession number GSE253245 (https://www.ncbi.nlm.nih.gov/geo/query/acc.cgi?acc-GSE253245).

## Code availability

Source code of whole genome sequencing data preprocessing, and germline SNV, indel, structural variant calling is available at https://github.com/nf-core/sarek. RNA-sequencing data processing and differential gene expression pipeline is available at https://github.com/MDNLAtlas/bulk-RNAseq-analysis-workflow. Scripts for variant annotation, statistical analysis, and data visualisation used in this study is available at https://github.com/MDNLAtlas/tinnitus-cognitive-genetics.

## Consent statement

All study participants provided informed consent prior to undergoing study procedures.

## Acknowledgements

We acknowledge all members of the Unification of Treatments and Interventions for Tinnitus Patients (UNITI) Project that have contributed to generate this dataset. List of participants in the UNITI Consortium available on https://uniti.tinnitusresearch.net.

## Supplementary materials

Supplementary material is available online.

