## Supplements for "Loss of function and structural variants in the *PRKRA* synaptic gene link mild cognitive impairment and tinnitus"

**Supplementary Appendix**

This appendix formed part of the original submission.

[**Supplementary Table 6: Rare structural variants (SV) (AF_gnomAD-NFE_ < 0.05) overlapped with synaptic genes previously identified as enriched with LoF/missense SNVs in MCI vs external controls.**	18](#_Toc230093642)

### **Supplementary Tables**

#### **Supplementary Table 1: Participant characteristics in the study cohort UNITI.**

|  | **UNITI** | **MCI**  **(20 ≤ MoCA ≤ 25)** | **Non-MCI**  **(MoCA > 25)** | **MCI vs. non-MCI** |
| --- | --- | --- | --- | --- |
| ***n*** | 294 | 75 | 201 |  |
| MoCA score^†^ | 27.0 (25.0, 29.0) | 24.0 (23.0, 25.0) | 28.0 (27.0, 29.0) |  |
| Age, years, mean (SD) | 51.6 (12.1) | 54.4 (11.0) | 50.6 (12.3) | Two-sided Wilcoxon test, * |
| Sex, *N* (% female) | 116 (39.5) | 33 (44.0) | 75 (37.3) | Fisher’s Exact test, *ns* |
| HL group, *N* (%) |  |  |  | Fisher’s Exact test, *ns* |
| Unilateral | 42 (14.3) | 16 (21.3) | 26 (12.9) |  |
| Bilateral | 42 (14.3) | 13 (17.3) | 28 (13.9) |  |
| Normal hearing | 203 (69.0) | 44 (58.7) | 142 (70.7) |  |
| Other/not reported | 7 (2.4) | 2 (2.7) | 5 (2.5) |  |
| PTA, db HL | 15.8 (10.0, 25.4) | 19.2 (13.3, 28.3) | 15.8 (10.0, 24.2) | Two-sided Wilcoxon test, ** |
| High-frequency HL, *N* (%) |  |  |  | Fisher’s Exact test, *ns* |
| Unilateral | 50 (17.0) | 13 (17.3) | 32 (15.9) |  |
| Bilateral | 137 (46.6) | 37 (49.3) | 94 (46.8) |  |
| Normal hearing | 93 (31.6) | 21 (28.0) | 65 (32.3) |  |
| Other/not reported | 14 (4.8) | 4 (5.4) | 10 (5.0) |  |
| HFHL, db HL | 37.5 (22.5, 53.6) | 45.0 (27.5, 56.3) | 37.5 (20.9, 51.6) | Two-sided Wilcoxon test, * |
| *APOE* ε4, *N* (% carrier*) | 74 (25.2) | 14 (18.7) | 56 (27.9) | Fisher’s Exact test, *ns* |
| THI score^†^ | 46.0 (30.5, 62.0) | 48.0 (36.0, 63.0) | 44.0 (30.0, 62.0) | Two-sided Wilcoxon test, *ns* |
| GÜF score^†^ | 13.0 (8.0, 19.0) | 13.5 (9.0, 20.0) | 12.0 (8.0, 18.8) | Two-sided Wilcoxon test, *ns* |
| PHQ-9 score | 7.0 (4.0, 10.0) | 7.0 (5.0, 9.75) | 6.0 (3.0, 10.0) | Two-sided Wilcoxon test, *ns* |
| MoCA vs. THI, *r* | -0.17, * | -0.31, * | -0.13, *ns* |  |
| MoCA vs. GÜF, *r* | -0.15, * | -0.42, *** | -0.09, *ns* |  |
| MoCA vs. PTA, *r* | -0.21, ** | -0.12, *ns* | -0.17, * |  |
| MoCA vs. HFHL, *r* | -0.20, ** | -0.16, *ns* | -0.17, * |  |
| MoCA vs. PHQ-9, *r* | -0.09, *ns* | -0.26, *ns* | -0.05, *ns* |  |
| THI vs. GÜF, *r* | 0.57, **** | 0.66, **** | 0.54, **** |  |
| THI vs. PTA, *r* | 0.10, *ns* | 0.01, *ns* | 0.10, *ns* |  |
| THI vs. HFHL, *r* | 0.02, *ns* | 0.13, *ns* | -0.05, *ns* |  |
| THI vs. PHQ-9, *r* | 0.67, **** | 0.66, **** | 0.68, **** |  |
| GÜF vs. PTA, *r* | 0.13, *ns* | 0.05, *ns* | 0.15, *ns* |  |
| GÜF vs. HFHL, *r* | -0.02, *ns* | 0.02, *ns* | -0.04, *ns* |  |
| GÜF vs. PHQ-9, *r* | 0.48, **** | 0.52, **** | 0.48, **** |  |
| PTA vs. HFHL, *r* | 0.60, **** | 0.55, **** | 0.59, **** |  |
| PTA vs. PHQ-9, *r* | 0.04, *ns* | 0.05, *ns* | 0.04, *ns* |  |
| HFHL vs. PHQ-9, *r* | -0.06, *ns* | 0.15, *ns* | -0.14, *ns* |  |

* APOE ε4 carriers have either ε2/ε4, ε3/ε4, or ε4/ε4 genotype.

^†^ All scores are reported as baseline (without treatment).

*Note*: all values are reported as median (IQR), unless otherwise noted; *P* values of Pearson’s correlation were corrected for multiple testing using BH method for UNITI, MCI, and non-MCI subgroups. Significance values: *ns* *p* > 0.05, * *p* < 0.05, ** *p* < 0.01, *** *p* < 0.001, **** *p* < 0.0001

Abbreviations: MCI, mild cognitive impairment; MoCA, Montreal Cognitive Assessment; PTA, Pure Tone Audiometry; HFHL, High-Frequency Hearing Loss; THI, Tinnitus Handicap Inventory; IQR, Interquartile Range; SD, Standard Deviation; *ns*, non-significant.

#### **Supplementary Table 2: Rare high-confident loss-of-function variants with CADD ≥ 20 in synaptic gene burden in the MCI subgroup.**

| Gene | Position | rsID | Exon | Number of individuals | Amino acid change | Consequence | CADD | AF | | |
| --- | --- | --- | --- | --- | --- | --- | --- | --- | --- | --- |
|  |  |  |  |  |  |  |  | **MCI** | **gnomADg**  **(NFE)*** | **gnomADg (global)**^†^ |
| *PRKRA* | chr2:178441705:C>  CAGTTTCCATAAATGACTCTAGCC  TGCAAATTGTAGTATATTCTCTCTTA | rs779497940 | - | 2 | - | splice_acceptor_variant | 23.2 | 0.013 | 0.000527 | 0.000308 |
|  | 2:178441705:C>  CAGTTTCCATAAATGACTCTAGCC  TGCAAATTGTAGTATATTCTCTCT | rs779497940 | - | 2 | - | splice_acceptor_variant | 23.2 | 0.013 | 1.5e-05 | 7e-06 |
| *HK3* | 5:176889700:G>GC | rs776857802 | 7/19 | 2 | G/GX | frameshift_variant | 26.9 | 0.013 | 0.000124 | 9.80E-05 |
| *IGFN1* | 1:201213165:C>T | rs377387750 | 12/24 | 2 | Q/* | stop_gained | 36 | 0.013 | 0.000248 | 0.000391 |
| *PIK3C2G* | 12:18338490:CCAAA>C | rs747980002 | 9/33 | 1 | TK/X | frameshift_variant | 28.6 | 0.0067 | 0.000171 | 9.8e-05 |
|  | 12:18562893:G>A | rs187352063 | - | 3 | - | splice_donor_variant | 31 | 0.02 | 0.002911 | 0.001849 |
|  | 12:18594540:CAGTT>C | rs556768893 | 30/33 | 1 | TV/X | frameshift_variant | 22.5 | 0.0067 | 0.000341 | 0.000398 |
| *GRAMD1A* | 19:35014388:G>A | rs192263602 | - | 4 | - | splice_donor_variant | 36 | 0.027 | 0.002153 | 0.00127 |
| *ENDOG* | 9:128822601:CAGTA>C | rs864309587 (rs1353989257) | 3/3 | 3 | SK/X | frameshift_variant | 27 | 0.02 | 0.001859 | 0.002136 |

* Non-Finnish European subpopulation in gnomAD genome v4.1.

^†^ All populations in gnomAD genome v4.1.

Abbreviations: MCI, mild cognitive impairment; NFE, Non-Finnish European; CADD, Combined Annotation Dependent Depletion; rsID, Reference SNP-cluster Identification; AF, Allele Frequency.

#### **Supplementary Table 3: Common variants used as ancestry markers (n = 111) in gene burden regions enriched for rare LoF variants in the MCI cohort.**

| **Gene** | **Variant ID (gnomAD)** | **Variant consequence** | **AF.MCI** | **AF.nonMCI** | **AF.nfe** | **Max ∆AF *** | **AF.afr** | **AF.amr** | **AF.asj** | **AF.eas** | **AF.mid** | **AF.fin** | **AF.ami** | **AF.oth** | **AF.sas** |
| --- | --- | --- | --- | --- | --- | --- | --- | --- | --- | --- | --- | --- | --- | --- | --- |
| *IGFN1* | 1-201193283-G-A | 5_prime_UTR_variant | 0.44 | 0.405 | 0.352 | 0.312 | 0.121 | 0.24 | 0.271 | 0.04 | 0.338 | 0.327 | 0.404 | 0.285 | 0.134 |
| *IGFN1* | 1-201197153-C-T | intron_variant | 0.333 | 0.366 | 0.405 | 0.183 | 0.589 | 0.333 | 0.324 | 0.526 | 0.337 | 0.516 | 0.368 | 0.432 | 0.498 |
| *IGFN1* | 1-201197255-G-A | missense_variant | 0.487 | 0.505 | 0.558 | 0.279 | 0.783 | 0.617 | 0.587 | 0.837 | 0.558 | 0.606 | 0.427 | 0.618 | 0.748 |
| *IGFN1* | 1-201197307-T-C | synonymous_variant | 0.927 | 0.93 | 0.927 | 0.103 | 0.865 | 0.824 | 0.866 | 0.838 | 0.892 | 0.951 | 0.861 | 0.905 | 0.876 |
| *IGFN1* | 1-201199371-G-A | synonymous_variant | 0.16 | 0.144 | 0.132 | 0.129 | 0.147 | 0.253 | 0.261 | 0.262 | 0.211 | 0.09 | 0.062 | 0.163 | 0.24 |
| *IGFN1* | 1-201200426-T-G | intron_variant | 0.153 | 0.142 | 0.125 | 0.137 | 0.173 | 0.249 | 0.259 | 0.261 | 0.213 | 0.088 | 0.059 | 0.16 | 0.239 |
| *IGFN1* | 1-201200473-C-T | intron_variant | 0.433 | 0.378 | 0.341 | 0.34 | 0.055 | 0.192 | 0.218 | 0.001 | 0.308 | 0.326 | 0.375 | 0.262 | 0.113 |
| *IGFN1* | 1-201201879-T-G | intron_variant | 0.767 | 0.736 | 0.735 | 0.221 | 0.554 | 0.62 | 0.645 | 0.957 | 0.671 | 0.806 | 0.83 | 0.714 | 0.678 |
| *IGFN1* | 1-201205076-C-T | splice_region_variant | 0.327 | 0.281 | 0.236 | 0.195 | 0.041 | 0.196 | 0.194 | 0.234 | 0.257 | 0.219 | 0.372 | 0.203 | 0.129 |
| *IGFN1* | 1-201206099-C-A | missense_variant | 0.753 | 0.736 | 0.718 | 0.122 | 0.84 | 0.746 | 0.773 | 0.684 | 0.758 | 0.654 | 0.655 | 0.727 | 0.752 |
| *IGFN1* | 1-201206146-A-G | missense_variant | 0.527 | 0.46 | 0.471 | 0.273 | 0.199 | 0.419 | 0.37 | 0.554 | 0.462 | 0.531 | 0.657 | 0.44 | 0.352 |
| *IGFN1* | 1-201206587-G-T | missense_variant | 0.76 | 0.736 | 0.717 | 0.124 | 0.841 | 0.746 | 0.773 | 0.684 | 0.758 | 0.655 | 0.655 | 0.727 | 0.752 |
| *IGFN1* | 1-201206890-A-G | missense_variant | 0.753 | 0.736 | 0.719 | 0.122 | 0.842 | 0.747 | 0.774 | 0.685 | 0.758 | 0.656 | 0.656 | 0.728 | 0.752 |
| *IGFN1* | 1-201206980-G-A | missense_variant | 0.26 | 0.323 | 0.343 | 0.155 | 0.219 | 0.311 | 0.277 | 0.339 | 0.258 | 0.336 | 0.188 | 0.328 | 0.374 |
| *IGFN1* | 1-201207840-A-G | missense_variant | 0.267 | 0.323 | 0.343 | 0.155 | 0.202 | 0.31 | 0.276 | 0.339 | 0.257 | 0.335 | 0.188 | 0.327 | 0.374 |
| *IGFN1* | 1-201208118-T-C | synonymous_variant | 0.267 | 0.323 | 0.343 | 0.156 | 0.228 | 0.312 | 0.291 | 0.339 | 0.26 | 0.335 | 0.188 | 0.33 | 0.374 |
| *IGFN1* | 1-201208150-C-T | missense_variant | 0.267 | 0.321 | 0.344 | 0.156 | 0.228 | 0.313 | 0.291 | 0.339 | 0.26 | 0.339 | 0.188 | 0.331 | 0.374 |
| *IGFN1* | 1-201208410-G-T | missense_variant | 0.267 | 0.326 | 0.343 | 0.157 | 0.203 | 0.32 | 0.277 | 0.34 | 0.258 | 0.336 | 0.186 | 0.327 | 0.378 |
| *IGFN1* | 1-201209342-G-A | synonymous_variant | 0.253 | 0.323 | 0.343 | 0.155 | 0.201 | 0.313 | 0.277 | 0.34 | 0.259 | 0.335 | 0.188 | 0.327 | 0.378 |
| *IGFN1* | 1-201212089-A-G | missense_variant | 0.267 | 0.328 | 0.343 | 0.155 | 0.265 | 0.315 | 0.292 | 0.34 | 0.262 | 0.336 | 0.188 | 0.333 | 0.377 |
| *IGFN1* | 1-201212792-G-T | synonymous_variant | 0.26 | 0.326 | 0.343 | 0.155 | 0.206 | 0.311 | 0.278 | 0.34 | 0.259 | 0.33 | 0.188 | 0.329 | 0.376 |
| *IGFN1* | 1-201213651-A-G | intron_variant | 0.26 | 0.326 | 0.343 | 0.158 | 0.207 | 0.306 | 0.279 | 0.34 | 0.259 | 0.331 | 0.186 | 0.329 | 0.378 |
| *IGFN1* | 1-201214235-C-T | synonymous_variant | 0.267 | 0.326 | 0.343 | 0.155 | 0.265 | 0.309 | 0.291 | 0.34 | 0.261 | 0.33 | 0.188 | 0.333 | 0.376 |
| *IGFN1* | 1-201214274-T-C | synonymous_variant | 0.267 | 0.326 | 0.343 | 0.155 | 0.213 | 0.318 | 0.278 | 0.374 | 0.257 | 0.334 | 0.188 | 0.33 | 0.377 |
| *IGFN1* | 1-201214969-G-A | intron_variant | 0.227 | 0.251 | 0.278 | 0.13 | 0.148 | 0.233 | 0.213 | 0.281 | 0.228 | 0.335 | 0.345 | 0.265 | 0.244 |
| *IGFN1* | 1-201215147-C-T | synonymous_variant | 0.267 | 0.323 | 0.343 | 0.158 | 0.259 | 0.322 | 0.294 | 0.376 | 0.262 | 0.335 | 0.186 | 0.336 | 0.377 |
| *IGFN1* | 1-201218636-T-C | synonymous_variant | 0.92 | 0.878 | 0.885 | 0.114 | 0.955 | 0.837 | 0.785 | 0.999 | 0.859 | 0.938 | 0.859 | 0.886 | 0.876 |
| *IGFN1* | 1-201218700-T-C | intron_variant | 0.92 | 0.878 | 0.885 | 0.113 | 0.954 | 0.836 | 0.783 | 0.998 | 0.857 | 0.938 | 0.86 | 0.885 | 0.874 |
| *IGFN1* | 1-201221604-G-A | synonymous_variant | 0.22 | 0.254 | 0.281 | 0.134 | 0.147 | 0.249 | 0.214 | 0.235 | 0.22 | 0.338 | 0.352 | 0.264 | 0.259 |
| *IGFN1* | 1-201225991-C-T | missense_variant | 0.68 | 0.662 | 0.683 | 0.407 | 0.276 | 0.59 | 0.546 | 0.829 | 0.598 | 0.662 | 0.717 | 0.647 | 0.599 |
| *IGFN1* | 1-201226168-G-A | intron_variant | 0.1 | 0.104 | 0.123 | 0.115 | 0.238 | 0.153 | 0.167 | 0.132 | 0.154 | 0.167 | 0.038 | 0.142 | 0.236 |
| *IGFN1* | 1-201228343-T-C | intron_variant | 0.413 | 0.361 | 0.347 | 0.268 | 0.079 | 0.252 | 0.309 | 0.283 | 0.393 | 0.359 | 0.585 | 0.318 | 0.203 |
| *UPK2* | 11-118958232-A-G | stop_retained_variant | 0.113 | 0.142 | 0.169 | 0.201 | 0.369 | 0.117 | 0.061 | 0.105 | 0.147 | 0.232 | 0.144 | 0.18 | 0.304 |
| *PIK3C2G* | 12-18282464-GCCC-G | inframe_deletion | 0.393 | 0.435 | 0.412 | 0.196 | 0.216 | 0.381 | 0.371 | 0.259 | 0.323 | 0.434 | 0.503 | 0.376 | 0.341 |
| *PIK3C2G* | 12-18320947-T-G | intron_variant | 0.127 | 0.109 | 0.116 | 0.116 | 0.102 | 0.062 | 0.065 | 0 | 0.111 | 0.132 | 0.073 | 0.095 | 0.058 |
| *PIK3C2G* | 12-18320994-T-C | synonymous_variant | 0.12 | 0.075 | 0.073 | 0.333 | 0.406 | 0.248 | 0.114 | 0.213 | 0.105 | 0.146 | 0.081 | 0.118 | 0.081 |
| *PIK3C2G* | 12-18346702-C-T | synonymous_variant | 0.087 | 0.067 | 0.061 | 0.221 | 0.174 | 0.282 | 0.106 | 0.176 | 0.083 | 0.142 | 0.045 | 0.089 | 0.05 |
| *PIK3C2G* | 12-18381932-C-A | intron_variant | 0.113 | 0.08 | 0.077 | 0.222 | 0.236 | 0.299 | 0.122 | 0.18 | 0.098 | 0.147 | 0.081 | 0.112 | 0.073 |
| *PIK3C2G* | 12-18488430-TTC-T | intron_variant | 0.087 | 0.072 | 0.068 | 0.107 | 0.175 | 0.046 | 0.073 | 0.04 | 0.101 | 0.046 | 0.086 | 0.073 | 0.09 |
| *PIK3C2G* | 12-18496123-C-T | missense_variant | 0.253 | 0.236 | 0.228 | 0.245 | 0.473 | 0.304 | 0.246 | 0.326 | 0.277 | 0.193 | 0.132 | 0.263 | 0.372 |
| *PIK3C2G* | 12-18503240-G-C | intron_variant | 0.167 | 0.142 | 0.158 | 0.157 | 0.241 | 0.11 | 0.157 | 0.001 | 0.179 | 0.177 | 0.249 | 0.15 | 0.077 |
| *PIK3C2G* | 12-18503291-G-A | synonymous_variant | 0.173 | 0.142 | 0.166 | 0.167 | 0.333 | 0.115 | 0.161 | 0.001 | 0.192 | 0.177 | 0.247 | 0.162 | 0.078 |
| *PIK3C2G* | 12-18567123-T-C | intron_variant | 0.187 | 0.182 | 0.194 | 0.133 | 0.06 | 0.148 | 0.104 | 0.252 | 0.086 | 0.232 | 0.074 | 0.177 | 0.241 |
| *PIK3C2G* | 12-18594594-C-T | intron_variant | 0.22 | 0.216 | 0.246 | 0.301 | 0.547 | 0.233 | 0.2 | 0.221 | 0.196 | 0.284 | 0.146 | 0.262 | 0.256 |
| *PIK3C2G* | 12-18648055-CA-C | 3_prime_UTR_variant | 0.127 | 0.085 | 0.096 | 0.105 | 0.201 | 0.063 | 0.13 | 0.064 | 0.147 | 0.094 | 0.02 | 0.106 | 0.063 |
| *RBM23* | 14-22901846-G-C | missense_variant | 0.3 | 0.294 | 0.307 | 0.221 | 0.086 | 0.277 | 0.222 | 0.285 | 0.285 | 0.25 | 0.268 | 0.279 | 0.266 |
| *RBM23* | 14-22902046-G-GGGC | inframe_insertion | 0.607 | 0.622 | 0.566 | 0.451 | 0.115 | 0.584 | 0.542 | 0.763 | 0.565 | 0.655 | 0.715 | 0.551 | 0.566 |
| *RBM23* | 14-22902059-A-G | synonymous_variant | 0.053 | 0.052 | 0.109 | 0.109 | 0.009 | 0.103 | 0.116 | 0.044 | 0.09 | 0.098 | 0 | 0.089 | 0.068 |
| *RBM23* | 14-22905226-G-A | synonymous_variant | 0.6 | 0.577 | 0.575 | 0.461 | 0.113 | 0.619 | 0.551 | 0.793 | 0.585 | 0.566 | 0.718 | 0.57 | 0.707 |
| *RBM23* | 14-22905653-C-T | synonymous_variant | 0.613 | 0.582 | 0.573 | 0.46 | 0.113 | 0.619 | 0.551 | 0.793 | 0.585 | 0.567 | 0.716 | 0.569 | 0.707 |
| *TTC6* | 14-37737613-G-A | intron_variant | 0.687 | 0.697 | 0.698 | 0.26 | 0.438 | 0.736 | 0.657 | 0.898 | 0.642 | 0.633 | 0.759 | 0.687 | 0.709 |
| *TTC6* | 14-37737803-A-AT | intron_variant | 0.68 | 0.697 | 0.564 | 0.208 | 0.408 | 0.66 | 0.558 | 0.772 | 0.544 | 0.556 | 0.753 | 0.555 | 0.591 |
| *TTC6* | 14-37749138-A-G | missense_variant | 0.16 | 0.167 | 0.164 | 0.265 | 0.091 | 0.385 | 0.139 | 0.428 | 0.111 | 0.175 | 0.113 | 0.171 | 0.143 |
| *TTC6* | 14-37749667-T-G | intron_variant | 0.607 | 0.617 | 0.619 | 0.276 | 0.423 | 0.702 | 0.6 | 0.895 | 0.59 | 0.555 | 0.743 | 0.627 | 0.662 |
| *TTC6* | 14-37753086-G-C | intron_variant | 0.993 | 0.945 | 0.952 | 0.218 | 0.734 | 0.955 | 0.926 | 1 | 0.912 | 0.97 | 0.984 | 0.934 | 0.905 |
| *TTC6* | 14-37790671-G-A | intron_variant | 0.167 | 0.152 | 0.155 | 0.228 | 0.161 | 0.159 | 0.142 | 0.383 | 0.161 | 0.148 | 0.147 | 0.168 | 0.18 |
| *TTC6* | 14-37790829-T-C | synonymous_variant | 0.16 | 0.152 | 0.159 | 0.18 | 0.105 | 0.156 | 0.144 | 0.338 | 0.157 | 0.147 | 0.148 | 0.16 | 0.12 |
| *TTC6* | 14-37790884-C-T | intron_variant | 1 | 0.99 | 0.997 | 0.271 | 0.725 | 0.929 | 0.995 | 0.827 | 0.975 | 0.952 | 1 | 0.959 | 0.978 |
| *TTC6* | 14-37796869-C-T | synonymous_variant | 0.487 | 0.448 | 0.464 | 0.389 | 0.075 | 0.272 | 0.438 | 0.29 | 0.352 | 0.412 | 0.558 | 0.4 | 0.309 |
| *TTC6* | 14-37808794-C-T | missense_variant | 0.347 | 0.381 | 0.36 | 0.276 | 0.167 | 0.447 | 0.415 | 0.084 | 0.409 | 0.388 | 0.267 | 0.348 | 0.446 |
| *TTC6* | 14-37841401-G-A | intron_variant | 0.913 | 0.908 | 0.914 | 0.581 | 0.333 | 0.848 | 0.867 | 0.879 | 0.79 | 0.928 | 0.853 | 0.859 | 0.908 |
| *VEZF1* | 17-57979189-C-G | synonymous_variant | 0.113 | 0.097 | 0.103 | 0.103 | 0.092 | 0.051 | 0.126 | 0 | 0.121 | 0.056 | 0.141 | 0.095 | 0.068 |
| *GRAMD1A* | 19-34996221-G-A | missense_variant | 0.18 | 0.231 | 0.192 | 0.126 | 0.102 | 0.161 | 0.319 | 0.278 | 0.279 | 0.223 | 0.294 | 0.205 | 0.156 |
| *GRAMD1A* | 19-35009838-A-T | intron_variant | 0.053 | 0.052 | 0.054 | 0.114 | 0.058 | 0.034 | 0.056 | 0.169 | 0.045 | 0.016 | 0.094 | 0.062 | 0.084 |
| *GRAMD1A* | 19-35010015-T-C | intron_variant | 0.427 | 0.47 | 0.395 | 0.197 | 0.593 | 0.366 | 0.528 | 0.424 | 0.537 | 0.397 | 0.492 | 0.432 | 0.37 |
| *GRAMD1A* | 19-35013723-C-A | intron_variant | 0.393 | 0.475 | 0.409 | 0.174 | 0.234 | 0.345 | 0.542 | 0.409 | 0.513 | 0.403 | 0.495 | 0.408 | 0.293 |
| *GRAMD1A* | 19-35015825-G-A | splice_region_variant | 0.347 | 0.368 | 0.309 | 0.155 | 0.276 | 0.249 | 0.464 | 0.24 | 0.456 | 0.318 | 0.366 | 0.316 | 0.183 |
| *GRAMD1A* | 19-35019198-G-C | synonymous_variant | 0.373 | 0.415 | 0.353 | 0.177 | 0.53 | 0.335 | 0.491 | 0.242 | 0.493 | 0.383 | 0.394 | 0.379 | 0.289 |
| *GRAMD1A* | 19-35019600-A-G | intron_variant | 0.333 | 0.366 | 0.312 | 0.196 | 0.116 | 0.239 | 0.461 | 0.239 | 0.453 | 0.319 | 0.366 | 0.307 | 0.182 |
| *GRAMD1A* | 19-35022842-T-G | intron_variant | 0.153 | 0.182 | 0.143 | 0.129 | 0.22 | 0.112 | 0.272 | 0.18 | 0.196 | 0.179 | 0.14 | 0.173 | 0.196 |
| *CAPN12* | 19-38731075-CCCATGCCCCA-C | intron_variant | 0.187 | 0.184 | 0.185 | 0.182 | 0.087 | 0.131 | 0.219 | 0.003 | 0.335 | 0.166 | 0.348 | 0.171 | 0.167 |
| *CAPN12* | 19-38733773-A-G | synonymous_variant | 0.753 | 0.784 | 0.761 | 0.195 | 0.956 | 0.66 | 0.756 | 0.693 | 0.765 | 0.73 | 0.884 | 0.771 | 0.85 |
| *CAPN12* | 19-38737604-T-C | missense_variant | 0.18 | 0.189 | 0.182 | 0.179 | 0.083 | 0.124 | 0.216 | 0.003 | 0.322 | 0.16 | 0.354 | 0.167 | 0.161 |
| *CAPN12* | 19-38738228-G-GCA | intron_variant | 0.753 | 0.774 | 0.759 | 0.207 | 0.552 | 0.624 | 0.709 | 0.677 | 0.732 | 0.729 | 0.884 | 0.736 | 0.848 |
| *CAPN12* | 19-38742506-A-G | splice_region_variant | 0.727 | 0.769 | 0.758 | 0.212 | 0.546 | 0.621 | 0.704 | 0.676 | 0.739 | 0.728 | 0.883 | 0.735 | 0.848 |
| *PRKRA* | 2-178432094-G-A | 3_prime_UTR_variant | 0.273 | 0.219 | 0.272 | 0.116 | 0.156 | 0.281 | 0.264 | 0.293 | 0.23 | 0.205 | 0.321 | 0.251 | 0.198 |
| *PRKRA* | 2-178432257-G-A | splice_region_variant | 0.013 | 0.01 | 0.154 | 0.154 | 0.025 | 0.073 | 0.065 | 0.065 | 0.049 | 0.03 | 0 | 0.107 | 0.047 |
| *PRKRA* | 2-178436244-G-A | synonymous_variant | 0.26 | 0.216 | 0.272 | 0.116 | 0.156 | 0.285 | 0.265 | 0.293 | 0.23 | 0.206 | 0.316 | 0.252 | 0.2 |
| *PRKRA* | 2-178451054-C-T | 5_prime_UTR_variant | 0.16 | 0.154 | 0.265 | 0.137 | 0.128 | 0.31 | 0.262 | 0.33 | 0.23 | 0.222 | 0.281 | 0.25 | 0.202 |
| *CDHR4* | 3-49791893-G-A | intron_variant | 0.327 | 0.281 | 0.318 | 0.26 | 0.058 | 0.185 | 0.232 | 0.204 | 0.194 | 0.245 | 0.41 | 0.272 | 0.292 |
| *CDHR4* | 3-49795355-G-A | missense_variant | 0.093 | 0.114 | 0.125 | 0.124 | 0.019 | 0.058 | 0.197 | 0.001 | 0.155 | 0.224 | 0.056 | 0.124 | 0.159 |
| *HK3* | 5-176881869-C-CAG | intron_variant | 0.18 | 0.192 | 0.18 | 0.137 | 0.138 | 0.301 | 0.196 | 0.318 | 0.25 | 0.189 | 0.09 | 0.197 | 0.27 |
| *HK3* | 5-176887433-C-T | intron_variant | 0.353 | 0.366 | 0.339 | 0.222 | 0.116 | 0.508 | 0.344 | 0.363 | 0.268 | 0.346 | 0.283 | 0.341 | 0.398 |
| *HK3* | 5-176887458-G-A | synonymous_variant | 0.347 | 0.368 | 0.339 | 0.175 | 0.217 | 0.514 | 0.361 | 0.362 | 0.272 | 0.346 | 0.281 | 0.35 | 0.397 |
| *HK3* | 5-176887638-G-A | synonymous_variant | 0.353 | 0.366 | 0.338 | 0.222 | 0.117 | 0.508 | 0.341 | 0.362 | 0.268 | 0.346 | 0.283 | 0.34 | 0.397 |
| *HK3* | 5-176888298-C-T | intron_variant | 0.36 | 0.368 | 0.34 | 0.223 | 0.117 | 0.497 | 0.344 | 0.361 | 0.268 | 0.344 | 0.281 | 0.34 | 0.399 |
| *ERMARD* | 6-169751670-C-T | splice_region_variant | 0.12 | 0.119 | 0.112 | 0.182 | 0.293 | 0.204 | 0.131 | 0.293 | 0.133 | 0.102 | 0.053 | 0.146 | 0.218 |
| *ERMARD* | 6-169755235-A-T | intron_variant | 0.233 | 0.231 | 0.217 | 0.189 | 0.406 | 0.292 | 0.276 | 0.325 | 0.246 | 0.172 | 0.134 | 0.247 | 0.296 |
| *ERMARD* | 6-169758927-G-A | intron_variant | 0.4 | 0.418 | 0.402 | 0.576 | 0.861 | 0.709 | 0.48 | 0.978 | 0.516 | 0.386 | 0.366 | 0.51 | 0.607 |
| *ERMARD* | 6-169762419-A-C | intron_variant | 0.12 | 0.137 | 0.141 | 0.217 | 0.358 | 0.109 | 0.166 | 0.053 | 0.23 | 0.093 | 0.124 | 0.164 | 0.21 |
| *ERMARD* | 6-169762441-C-T | synonymous_variant | 0.08 | 0.087 | 0.088 | 0.188 | 0.245 | 0.198 | 0.098 | 0.276 | 0.087 | 0.084 | 0.046 | 0.119 | 0.179 |
| *ERMARD* | 6-169776311-G-A | intron_variant | 0.233 | 0.229 | 0.216 | 0.266 | 0.482 | 0.298 | 0.267 | 0.329 | 0.23 | 0.171 | 0.126 | 0.252 | 0.287 |
| *ERMARD* | 6-169776371-A-G | missense_variant | 0.227 | 0.224 | 0.216 | 0.205 | 0.421 | 0.293 | 0.267 | 0.328 | 0.227 | 0.171 | 0.126 | 0.246 | 0.286 |
| *ERMARD* | 6-169776551-C-T | synonymous_variant | 0.233 | 0.226 | 0.216 | 0.265 | 0.481 | 0.303 | 0.267 | 0.328 | 0.229 | 0.172 | 0.127 | 0.251 | 0.287 |
| *ERMARD* | 6-169776552-A-G | missense_variant | 0.233 | 0.226 | 0.216 | 0.265 | 0.481 | 0.303 | 0.267 | 0.328 | 0.229 | 0.172 | 0.126 | 0.251 | 0.287 |
| *ENTPD4* | 8-23434322-T-C | synonymous_variant | 0.133 | 0.169 | 0.179 | 0.12 | 0.059 | 0.178 | 0.161 | 0.06 | 0.194 | 0.157 | 0.085 | 0.168 | 0.098 |
| *ENTPD4* | 8-23436879-C-T | intron_variant | 0.127 | 0.169 | 0.178 | 0.119 | 0.063 | 0.179 | 0.16 | 0.059 | 0.191 | 0.157 | 0.084 | 0.168 | 0.097 |
| *ENTPD4* | 8-23436899-A-G | intron_variant | 0.127 | 0.172 | 0.179 | 0.119 | 0.11 | 0.187 | 0.162 | 0.06 | 0.202 | 0.157 | 0.084 | 0.172 | 0.098 |
| *ENTPD4* | 8-23437248-T-C | missense_variant | 0.047 | 0.06 | 0.059 | 0.218 | 0.154 | 0.225 | 0.042 | 0.276 | 0.038 | 0.103 | 0.072 | 0.083 | 0.102 |
| *ENTPD4* | 8-23443827-A-C | intron_variant | 0.273 | 0.279 | 0.274 | 0.172 | 0.289 | 0.243 | 0.257 | 0.446 | 0.223 | 0.216 | 0.331 | 0.285 | 0.349 |
| *ENTPD4* | 8-23444651-A-G | intron_variant | 0.353 | 0.336 | 0.313 | 0.312 | 0.625 | 0.297 | 0.365 | 0.447 | 0.331 | 0.288 | 0.354 | 0.353 | 0.37 |
| *RABEPK* | 9-125200637-G-A | synonymous_variant | 0.407 | 0.515 | 0.453 | 0.169 | 0.487 | 0.417 | 0.503 | 0.622 | 0.458 | 0.287 | 0.471 | 0.454 | 0.421 |
| *RABEPK* | 9-125200693-G-A | missense_variant | 0.1 | 0.149 | 0.14 | 0.135 | 0.027 | 0.078 | 0.19 | 0.005 | 0.11 | 0.121 | 0.202 | 0.119 | 0.122 |
| *RABEPK* | 9-125203121-A-C | intron_variant | 0.1 | 0.149 | 0.143 | 0.139 | 0.105 | 0.079 | 0.192 | 0.004 | 0.117 | 0.12 | 0.201 | 0.132 | 0.122 |
| *RABEPK* | 9-125207506-G-A | intron_variant | 0.413 | 0.512 | 0.463 | 0.197 | 0.57 | 0.444 | 0.508 | 0.661 | 0.485 | 0.292 | 0.469 | 0.479 | 0.45 |
| *RABEPK* | 9-125207551-C-T | intron_variant | 0.1 | 0.149 | 0.144 | 0.141 | 0.027 | 0.073 | 0.191 | 0.004 | 0.113 | 0.12 | 0.201 | 0.128 | 0.122 |
| *RABEPK* | 9-125213375-C-T | missense_variant | 0.1 | 0.149 | 0.145 | 0.141 | 0.125 | 0.081 | 0.192 | 0.004 | 0.118 | 0.12 | 0.201 | 0.135 | 0.122 |
| *RABEPK* | 9-125220496-CT-C | intron_variant | 0.593 | 0.493 | 0.538 | 0.17 | 0.45 | 0.701 | 0.497 | 0.46 | 0.541 | 0.708 | 0.529 | 0.535 | 0.599 |
| *RABEPK* | 9-125233767-A-G | synonymous_variant | 0.067 | 0.09 | 0.067 | 0.126 | 0.194 | 0.047 | 0.109 | 0.14 | 0.119 | 0.094 | 0.075 | 0.088 | 0.107 |
| *ENDOG* | 9-128818719-C-T | missense_variant | 0.687 | 0.779 | 0.748 | 0.413 | 0.335 | 0.764 | 0.768 | 0.76 | 0.68 | 0.836 | 0.794 | 0.703 | 0.616 |
| *ENDOG* | 9-128820891-T-C | intron_variant | 0.72 | 0.789 | 0.748 | 0.404 | 0.344 | 0.784 | 0.761 | 0.738 | 0.688 | 0.832 | 0.793 | 0.706 | 0.625 |

* Maximum of allele frequency difference between NFE vs. non-NFE populations in gnomAD

Abbreviations: MCI, mild cognitive impairment; AF, Allele frequency; nfe, Non-Finnish European; afr, African American; amr: Admixed American; asj; Ashkenazi Jewish; eas, East Asian; mid, Middle Eastern; fin, Finnish European; ami, Amish; asa, South Asian; oth, Remaining.

#### **Supplementary Table 4: Rare missense variants with CADD ≥ 20 in synaptic gene burden in the MCI subgroup.**

| Gene | Position | rsID | Exon | Number of individuals | Amino acid change | Consequence | CADD | ACMG | AF | | |
| --- | --- | --- | --- | --- | --- | --- | --- | --- | --- | --- | --- |
|  |  |  |  |  |  |  |  |  | **MCI** | **gnomADg (NFE)*** | **gnomADg (global)**^†^ |
| *EIF4G1* | chr3:184315833:G>C | - | 3/33 | 4 | A13P | missense_variant | 24.2 | Likely benign (BP4, BP6) | 0.027 | 0.000177 | 0.000268 |
| *ERBB3* | chr12:56101239:G>A | rs2271188 | 27/28 | 2 | R1127H | missense_variant | 23.9 | Benign (BP4, BP6, BS1, BS2) | 0.013 | 3.10E-05 | 0.000419 |
| *ATP12A* | chr13:24690378:C>A | rs141275215 | 6/23 | 1 | P196H | missense_variant | 22.5 | Likely benign (BP4) | 0.0067 | 0.001986 | 0.001529 |
|  | chr13:24698695:G>A | - | 12/23 | 8 | R517H | missense_variant | 22 | Benign (BP4, BP6, BS1, BS2) | 0.053 | 0.012222 | 0.017248 |
|  | chr13:24706425:C>A | rs61998252 | 15/23 | 6 | Q711K | missense_variant | 24.8 | Benign (BP4, BS1, BS2) | 0.04 | 0.008701 | 0.006335 |
| *EP400* | chr12:132053386:A>C | - | 43/53 | 2 | Q2506P | missense_variant | 22.2 | VUS | 0.013 | 1.60E-05 | 2.90E-05 |
| *PRX* | chr19:40397301:G>A | - | 7/7 | 2 | P351S | missense_variant | 24.9 | Benign (BP4, BP6, BA1) | 0.013 | 0.000217 | 0.020233 |
| *KIAA1328* | chr18:37067460:C>T | - | 7/10 | 2 | R383C | missense_variant | 25.8 | Benign (BP4, BA1) | 0.013 | 0.000341 | 0.032436 |
| *ANO2* | chr12:5744308:C>T | rs17788563 | 12/25 | 2 | M400I | missense_variant | 20.7 | Benign (BP4, BS1, BS2) | 0.013 | 0.000356 | 0.004313 |
| *ANKRD52* | chr12:56248815:C>T | rs201680602 | 16/28 | 3 | G550S | missense_variant | 26.3 | Likely benign (PP3, BP4, BS2) | 0.02 | 0.001177 | 0.000796 |
| *ATP6V1E2* | chr2:46512350:A>G | rs147855126 | 5/5 | 4 | L121P | missense_variant | 27.4 | Benign (PP3, BP4, BS2) | 0.027 | 0.002447 | 0.001983 |
| *SCN5A* | chr3:38633208:G>A | rs6791924 | 2/28 | 3 | R34C | missense_variant | 23.6 | Benign (PM1, BP4, BP6, BA1) | 0.02 | 0.001255 | 0.030206 |
| *REEP6* | chr19:1496349:G>A | rs150955785 | 4/5 | 2 | R138H | missense_variant | 20.2 | Benign (BP4, BP6, BS1, BS2) | 0.013 | 0.000496 | 0.000691 |
| *AGAP3* | chr7:151141913:T>G | - | 14/18 | 4 | F607C | missense_variant | 32 | VUS | 0.027 | 0.000221 | 0.004211 |
|  | chr7:151143876:C>T | - | 18/18 | 1 | P890L | missense_variant | 22.8 | Benign (BP4, BP6, BS2) | 0.0067 | 0.000284 | 0.003496 |
| *TBCC* | chr6:42745816:C>A | rs144361927 | 1/1 | 3 | E86D | missense_variant | 23.2 | Benign (BP4, BS2) | 0.02 | 0.001533 | 0.001186 |
| *TMED3* | chr15:79313844:G>A | rs3784543 | 2/3 | 2 | D86N | missense_variant | 27.3 | Benign (BP4, BA1) | 0.013 | 0.000557 | 0.006818 |
| *TNKS1BP1* | chr11:57320343:C>T | rs143756483 | 3/12 | 2 | R155H | missense_variant | 26.4 | Benign (BP4, BS2) | 0.013 | 0.00582 | 0.01708 |
|  | chr11:57320509:C>G | rs34448143 | 3/12 | 10 | A100P | missense_variant | 21.7 | Benign (BP4, BA1) | 0.067 | 0.00422 | 0.014676 |

* Non-Finnish European subpopulation in gnomAD genome v4.1.

^†^ All populations in gnomAD genome v4.1.

Abbreviations: MCI, mild cognitive impairment; NFE, Non-Finnish European; CADD, Combined Annotation Dependent Depletion; ACMG, American College of Medical Genetics and genomics; rsID, Reference SNP-cluster Identification; AF, Allele Frequency; VUS, Variant of Uncertain Significance.

#### **Supplementary Table 5: Common variants used as ancestry markers (n = 193) in gene burden regions enriched for rare missense variants in the MCI cohort.**

| **Gene** | **Variant ID (gnomAD)** | **Variant consequence** | **AF.MCI** | **AF.nonMCI** | **AF.nfe** | **Max ∆AF** | **AF.afr** | **AF.amr** | **AF.asj** | **AF.eas** | **AF.mid** | **AF.fin** | **AF.ami** | **AF.oth** | **AF.sas** |
| --- | --- | --- | --- | --- | --- | --- | --- | --- | --- | --- | --- | --- | --- | --- | --- |
| *C10orf67* | 10-23344637-C-A | synonymous_variant | 0.48 | 0.517 | 0.468 | 0.355 | 0.709 | 0.658 | 0.604 | 0.823 | 0.628 | 0.443 | 0.399 | 0.543 | 0.591 |
| *TNKS1BP1* | 11-57309990-T-C | synonymous_variant | 0.413 | 0.336 | 0.33 | 0.104 | 0.434 | 0.236 | 0.327 | 0.328 | 0.368 | 0.25 | 0.376 | 0.32 | 0.278 |
| *TNKS1BP1* | 11-57311247-C-T | synonymous_variant | 0.32 | 0.281 | 0.291 | 0.191 | 0.1 | 0.179 | 0.27 | 0.22 | 0.267 | 0.221 | 0.373 | 0.251 | 0.216 |
| *TNKS1BP1* | 11-57311402-C-G | 5_prime_UTR_variant | 0.067 | 0.1 | 0.099 | 0.25 | 0.349 | 0.104 | 0.091 | 0.014 | 0.1 | 0.047 | 0.111 | 0.11 | 0.046 |
| *TNKS1BP1* | 11-57313723-G-C | missense_variant | 0.473 | 0.525 | 0.533 | 0.402 | 0.13 | 0.649 | 0.55 | 0.666 | 0.486 | 0.594 | 0.492 | 0.533 | 0.623 |
| *LGALS12* | 11-63511149-C-G | intron_variant | 0.44 | 0.463 | 0.528 | 0.397 | 0.131 | 0.487 | 0.418 | 0.247 | 0.378 | 0.612 | 0.49 | 0.458 | 0.424 |
| *LGALS12* | 11-63515748-C-G | intron_variant | 0.2 | 0.226 | 0.184 | 0.312 | 0.496 | 0.146 | 0.149 | 0.199 | 0.196 | 0.21 | 0.168 | 0.217 | 0.3 |
| *EP400* | 12-131979805-C-T | intron_variant | 0.113 | 0.085 | 0.079 | 0.168 | 0.247 | 0.11 | 0.132 | 0.023 | 0.111 | 0.043 | 0.16 | 0.098 | 0.081 |
| *EP400* | 12-132013715-T-C | intron_variant | 0.147 | 0.109 | 0.093 | 0.679 | 0.771 | 0.122 | 0.15 | 0.121 | 0.189 | 0.181 | 0.16 | 0.168 | 0.204 |
| *EP400* | 12-132021011-A-G | intron_variant | 0.167 | 0.122 | 0.109 | 0.539 | 0.648 | 0.158 | 0.156 | 0.121 | 0.186 | 0.19 | 0.16 | 0.171 | 0.221 |
| *EP400* | 12-132023844-A-G | synonymous_variant | 0.153 | 0.119 | 0.11 | 0.604 | 0.713 | 0.159 | 0.159 | 0.12 | 0.19 | 0.19 | 0.16 | 0.178 | 0.211 |
| *EP400* | 12-132029945-C-T | intron_variant | 0.147 | 0.122 | 0.11 | 0.604 | 0.713 | 0.159 | 0.159 | 0.121 | 0.188 | 0.19 | 0.16 | 0.178 | 0.211 |
| *EP400* | 12-132045024-TTCTGCTGTCTGCCTG-T | intron_variant | 0.093 | 0.08 | 0.07 | 0.143 | 0.015 | 0.212 | 0.045 | 0.107 | 0.052 | 0.078 | 0.083 | 0.071 | 0.068 |
| *EP400* | 12-132054906-G-A | intron_variant | 0.113 | 0.082 | 0.079 | 0.167 | 0.246 | 0.107 | 0.132 | 0.024 | 0.105 | 0.042 | 0.16 | 0.097 | 0.073 |
| *EP400* | 12-132062548-A-ACAG | inframe_insertion | 0.273 | 0.289 | 0.307 | 0.178 | 0.129 | 0.332 | 0.268 | 0.199 | 0.269 | 0.273 | 0.334 | 0.273 | 0.244 |
| *EP400* | 12-132062548-A-G | synonymous_variant | 0.013 | 0.027 | 0.061 | 0.104 | 0.165 | 0.088 | 0.092 | 0.023 | 0.085 | 0.017 | 0.063 | 0.073 | 0.073 |
| *EP400* | 12-132067495-G-A | intron_variant | 0.153 | 0.124 | 0.113 | 0.422 | 0.021 | 0.189 | 0.17 | 0.535 | 0.086 | 0.11 | 0.115 | 0.134 | 0.078 |
| *EP400* | 12-132077442-C-T | synonymous_variant | 0.053 | 0.057 | 0.052 | 0.102 | 0.138 | 0.039 | 0.017 | 0.095 | 0.047 | 0.154 | 0.043 | 0.058 | 0.072 |
| *EP400* | 12-132077581-G-A | missense_variant | 0.113 | 0.174 | 0.144 | 0.138 | 0.134 | 0.077 | 0.088 | 0.007 | 0.118 | 0.133 | 0.144 | 0.119 | 0.045 |
| *EP400* | 12-132077730-G-A | 3_prime_UTR_variant | 0.06 | 0.07 | 0.058 | 0.215 | 0.273 | 0.062 | 0.032 | 0.104 | 0.088 | 0.155 | 0.1 | 0.079 | 0.075 |
| *EMP1* | 12-13213665-C-T | intron_variant | 0.14 | 0.174 | 0.206 | 0.175 | 0.031 | 0.079 | 0.156 | 0.141 | 0.151 | 0.177 | 0.184 | 0.175 | 0.168 |
| *ANO2* | 12-5565588-G-A | synonymous_variant | 0.473 | 0.5 | 0.466 | 0.227 | 0.239 | 0.333 | 0.494 | 0.301 | 0.391 | 0.397 | 0.489 | 0.439 | 0.413 |
| *ERBB3* | 12-56083910-A-T | splice_region_variant | 0.6 | 0.525 | 0.577 | 0.25 | 0.327 | 0.731 | 0.551 | 0.792 | 0.7 | 0.603 | 0.419 | 0.595 | 0.714 |
| *ERBB3* | 12-56100038-A-C | intron_variant | 0.373 | 0.455 | 0.408 | 0.224 | 0.183 | 0.372 | 0.409 | 0.342 | 0.292 | 0.39 | 0.505 | 0.378 | 0.294 |
| *ERBB3* | 12-56101207-G-A | synonymous_variant | 0.347 | 0.45 | 0.402 | 0.281 | 0.121 | 0.352 | 0.37 | 0.319 | 0.263 | 0.385 | 0.504 | 0.358 | 0.258 |
| *ERBB3* | 12-56101214-A-T | missense_variant | 0.127 | 0.102 | 0.115 | 0.114 | 0.084 | 0.062 | 0.108 | 0.001 | 0.087 | 0.117 | 0.141 | 0.1 | 0.079 |
| *ERBB3* | 12-56101522-C-T | splice_region_variant | 0.127 | 0.1 | 0.115 | 0.114 | 0.157 | 0.067 | 0.108 | 0.001 | 0.091 | 0.117 | 0.141 | 0.105 | 0.079 |
| *ANKRD52* | 12-56254127-G-C | synonymous_variant | 0.92 | 0.935 | 0.932 | 0.459 | 0.473 | 0.907 | 0.957 | 0.965 | 0.946 | 0.944 | 0.951 | 0.908 | 0.977 |
| *ANKRD52* | 12-56255817-A-G | synonymous_variant | 0.92 | 0.943 | 0.932 | 0.456 | 0.476 | 0.905 | 0.957 | 0.965 | 0.95 | 0.945 | 0.951 | 0.908 | 0.977 |
| *ANO2* | 12-5647658-C-T | intron_variant | 0.887 | 0.799 | 0.827 | 0.167 | 0.837 | 0.682 | 0.815 | 0.688 | 0.847 | 0.788 | 0.799 | 0.807 | 0.66 |
| *ANO2* | 12-5647814-G-A | intron_variant | 0.167 | 0.114 | 0.121 | 0.117 | 0.178 | 0.069 | 0.105 | 0.004 | 0.088 | 0.068 | 0.122 | 0.109 | 0.093 |
| *ANO2* | 12-5647863-C-T | intron_variant | 0.413 | 0.358 | 0.366 | 0.333 | 0.651 | 0.256 | 0.392 | 0.032 | 0.335 | 0.266 | 0.429 | 0.349 | 0.243 |
| *ANO2* | 12-5732567-A-C | missense_variant | 0.08 | 0.077 | 0.08 | 0.15 | 0.23 | 0.104 | 0.113 | 0.132 | 0.074 | 0.09 | 0.047 | 0.099 | 0.08 |
| *ANO2* | 12-5732675-G-C | intron_variant | 0.42 | 0.42 | 0.454 | 0.261 | 0.194 | 0.437 | 0.344 | 0.543 | 0.426 | 0.533 | 0.353 | 0.428 | 0.474 |
| *ANO2* | 12-5751013-T-C | intron_variant | 0.453 | 0.433 | 0.462 | 0.344 | 0.118 | 0.343 | 0.352 | 0.582 | 0.437 | 0.528 | 0.323 | 0.427 | 0.485 |
| *ANO2* | 12-5851882-G-A | intron_variant | 0.98 | 0.965 | 0.981 | 0.169 | 0.812 | 0.974 | 0.94 | 1 | 0.951 | 0.993 | 0.99 | 0.964 | 0.978 |
| *ANO2* | 12-5920988-G-A | intron_variant | 0.193 | 0.211 | 0.196 | 0.123 | 0.073 | 0.128 | 0.22 | 0.164 | 0.196 | 0.142 | 0.253 | 0.19 | 0.197 |
| *ANO2* | 12-5920999-G-A | intron_variant | 0.193 | 0.211 | 0.196 | 0.129 | 0.067 | 0.128 | 0.219 | 0.164 | 0.197 | 0.141 | 0.253 | 0.19 | 0.196 |
| *ANO2* | 12-5921135-G-A | missense_variant | 0.153 | 0.112 | 0.119 | 0.1 | 0.019 | 0.058 | 0.116 | 0.025 | 0.13 | 0.076 | 0.116 | 0.102 | 0.129 |
| *ANO2* | 12-5921136-T-C | synonymous_variant | 0.187 | 0.209 | 0.197 | 0.129 | 0.068 | 0.13 | 0.225 | 0.164 | 0.199 | 0.144 | 0.253 | 0.191 | 0.208 |
| *ANO2* | 12-5921239-A-G | missense_variant | 0.193 | 0.211 | 0.199 | 0.131 | 0.068 | 0.131 | 0.225 | 0.165 | 0.201 | 0.144 | 0.253 | 0.195 | 0.233 |
| *ANO2* | 12-5921271-A-G | synonymous_variant | 0.04 | 0.057 | 0.064 | 0.105 | 0.018 | 0.062 | 0.044 | 0.169 | 0.051 | 0.044 | 0.023 | 0.077 | 0.169 |
| *ANO2* | 12-5922606-C-T | intron_variant | 0.173 | 0.1 | 0.114 | 0.18 | 0.295 | 0.085 | 0.123 | 0.062 | 0.157 | 0.074 | 0.159 | 0.121 | 0.128 |
| *ANO2* | 12-5945229-A-G | 5_prime_UTR_variant | 0.94 | 0.958 | 0.964 | 0.307 | 0.657 | 0.92 | 0.951 | 0.843 | 0.908 | 0.942 | 0.927 | 0.927 | 0.878 |
| *ATP12A* | 13-24685442-C-T | intron_variant | 0.1 | 0.104 | 0.133 | 0.133 | 0.054 | 0.064 | 0.083 | 0 | 0.063 | 0.094 | 0.112 | 0.104 | 0.068 |
| *ATP12A* | 13-24690528-G-T | intron_variant | 0.193 | 0.182 | 0.223 | 0.279 | 0.171 | 0.44 | 0.183 | 0.502 | 0.186 | 0.211 | 0.176 | 0.241 | 0.315 |
| *ATP12A* | 13-24690577-G-A | intron_variant | 0.2 | 0.182 | 0.223 | 0.278 | 0.171 | 0.44 | 0.183 | 0.502 | 0.185 | 0.211 | 0.174 | 0.241 | 0.316 |
| *ATP12A* | 13-24690585-C-T | intron_variant | 0.2 | 0.182 | 0.223 | 0.279 | 0.171 | 0.439 | 0.183 | 0.502 | 0.185 | 0.211 | 0.174 | 0.241 | 0.316 |
| *ATP12A* | 13-24690965-A-G | splice_region_variant | 0.187 | 0.182 | 0.223 | 0.279 | 0.171 | 0.44 | 0.183 | 0.502 | 0.186 | 0.211 | 0.177 | 0.241 | 0.316 |
| *ATP12A* | 13-24691001-T-C | synonymous_variant | 0.2 | 0.182 | 0.223 | 0.279 | 0.171 | 0.44 | 0.183 | 0.503 | 0.186 | 0.211 | 0.177 | 0.241 | 0.316 |
| *ATP12A* | 13-24692748-C-T | intron_variant | 0.193 | 0.177 | 0.222 | 0.276 | 0.179 | 0.438 | 0.175 | 0.498 | 0.19 | 0.206 | 0.178 | 0.239 | 0.302 |
| *ATP12A* | 13-24692794-C-A | synonymous_variant | 0.093 | 0.075 | 0.09 | 0.416 | 0.114 | 0.376 | 0.093 | 0.507 | 0.128 | 0.107 | 0.068 | 0.136 | 0.222 |
| *ATP12A* | 13-24692950-C-T | intron_variant | 0.193 | 0.182 | 0.233 | 0.276 | 0.18 | 0.44 | 0.181 | 0.509 | 0.19 | 0.219 | 0.188 | 0.248 | 0.311 |
| *ATP12A* | 13-24694638-A-G | intron_variant | 0.193 | 0.182 | 0.232 | 0.278 | 0.194 | 0.441 | 0.179 | 0.51 | 0.209 | 0.223 | 0.187 | 0.249 | 0.31 |
| *ATP12A* | 13-24698732-C-T | synonymous_variant | 0.187 | 0.172 | 0.149 | 0.288 | 0.436 | 0.118 | 0.184 | 0.17 | 0.184 | 0.129 | 0.273 | 0.181 | 0.175 |
| *ATP12A* | 13-24709356-C-T | splice_region_variant | 0.12 | 0.127 | 0.116 | 0.111 | 0.054 | 0.07 | 0.139 | 0.129 | 0.128 | 0.11 | 0.227 | 0.126 | 0.142 |
| *ATP12A* | 13-24709519-G-C | intron_variant | 0.473 | 0.495 | 0.47 | 0.178 | 0.334 | 0.292 | 0.467 | 0.322 | 0.434 | 0.494 | 0.414 | 0.442 | 0.359 |
| *OR5AU1* | 14-21155131-T-C | missense_variant | 0.753 | 0.774 | 0.782 | 0.288 | 0.831 | 0.671 | 0.743 | 0.494 | 0.804 | 0.775 | 0.694 | 0.75 | 0.724 |
| *OR5AU1* | 14-21155330-C-T | synonymous_variant | 0.173 | 0.219 | 0.177 | 0.531 | 0.668 | 0.405 | 0.227 | 0.708 | 0.223 | 0.191 | 0.087 | 0.264 | 0.415 |
| *OR5AU1* | 14-21155489-G-A | synonymous_variant | 0.593 | 0.557 | 0.585 | 0.48 | 0.361 | 0.341 | 0.528 | 0.105 | 0.571 | 0.577 | 0.596 | 0.513 | 0.387 |
| *OR5AU1* | 14-21155677-G-A | missense_variant | 0.767 | 0.741 | 0.776 | 0.304 | 0.664 | 0.649 | 0.763 | 0.472 | 0.773 | 0.711 | 0.667 | 0.733 | 0.731 |
| *OR5AU1* | 14-21156038-T-C | 5_prime_UTR_variant | 0.72 | 0.729 | 0.746 | 0.342 | 0.404 | 0.606 | 0.773 | 0.43 | 0.743 | 0.636 | 0.716 | 0.693 | 0.745 |
| *KCNK13* | 14-90184689-G-A | missense_variant | 0.04 | 0.072 | 0.053 | 0.237 | 0.291 | 0.046 | 0.043 | 0.108 | 0.079 | 0.068 | 0.011 | 0.078 | 0.157 |
| *KCNK13* | 14-90185013-G-A | 3_prime_UTR_variant | 0.32 | 0.351 | 0.299 | 0.267 | 0.052 | 0.177 | 0.289 | 0.522 | 0.289 | 0.334 | 0.566 | 0.287 | 0.329 |
| *TMED3* | 15-79382959-T-C | intron_variant | 0.1 | 0.152 | 0.119 | 0.127 | 0.246 | 0.189 | 0.139 | 0.195 | 0.154 | 0.181 | 0.177 | 0.14 | 0.096 |
| *FSD2* | 15-82759440-C-T | missense_variant | 0.173 | 0.179 | 0.173 | 0.172 | 0.135 | 0.164 | 0.125 | 0.001 | 0.155 | 0.16 | 0.041 | 0.148 | 0.052 |
| *FSD2* | 15-82759441-G-A | synonymous_variant | 0.1 | 0.102 | 0.107 | 0.142 | 0.201 | 0.193 | 0.11 | 0.25 | 0.124 | 0.121 | 0.218 | 0.122 | 0.063 |
| *FSD2* | 15-82772260-C-A | intron_variant | 0.127 | 0.142 | 0.167 | 0.154 | 0.031 | 0.254 | 0.121 | 0.244 | 0.121 | 0.186 | 0.042 | 0.166 | 0.321 |
| *FSD2* | 15-82778879-T-G | missense_variant | 0.127 | 0.142 | 0.167 | 0.153 | 0.031 | 0.26 | 0.119 | 0.218 | 0.12 | 0.184 | 0.042 | 0.165 | 0.321 |
| *FSD2* | 15-82786671-G-A | intron_variant | 0.147 | 0.142 | 0.146 | 0.144 | 0.137 | 0.145 | 0.088 | 0.001 | 0.146 | 0.156 | 0.035 | 0.127 | 0.051 |
| *ZNF768* | 16-30525351-A-G | synonymous_variant | 0.627 | 0.547 | 0.601 | 0.468 | 0.915 | 0.394 | 0.594 | 0.134 | 0.611 | 0.445 | 0.601 | 0.601 | 0.82 |
| *ZNF768* | 16-30525597-C-G | missense_variant | 0.347 | 0.331 | 0.396 | 0.394 | 0.478 | 0.223 | 0.322 | 0.002 | 0.312 | 0.259 | 0.437 | 0.344 | 0.184 |
| *DBNDD1* | 16-90019379-G-C | 5_prime_UTR_variant | 0.04 | 0.06 | 0.075 | 0.586 | 0.086 | 0.226 | 0.06 | 0.661 | 0.062 | 0.256 | 0.003 | 0.115 | 0.136 |
| *KIAA1328* | 18-36885549-T-TA | splice_region_variant | 0.267 | 0.159 | 0.159 | 0.159 | 0.041 | 0.159 | 0.178 | 0.001 | 0.135 | 0.15 | 0.115 | 0.143 | 0.097 |
| *KIAA1328* | 18-37067213-T-C | synonymous_variant | 0.087 | 0.159 | 0.137 | 0.457 | 0.594 | 0.144 | 0.188 | 0.164 | 0.118 | 0.155 | 0.173 | 0.169 | 0.109 |
| *KIAA1328* | 18-37067360-T-A | synonymous_variant | 0.06 | 0.132 | 0.137 | 0.461 | 0.598 | 0.144 | 0.188 | 0.163 | 0.118 | 0.155 | 0.174 | 0.169 | 0.109 |
| *KIAA1328* | 18-37067360-T-G | synonymous_variant | 0.233 | 0.134 | 0.186 | 0.185 | 0.043 | 0.179 | 0.204 | 0.001 | 0.162 | 0.167 | 0.117 | 0.168 | 0.116 |
| *KIAA1328* | 18-37067562-CT-C | intron_variant | 0.333 | 0.291 | 0.371 | 0.13 | 0.501 | 0.367 | 0.403 | 0.3 | 0.364 | 0.368 | 0.277 | 0.379 | 0.334 |
| *KIAA1328* | 18-37067597-G-A | intron_variant | 0.36 | 0.316 | 0.323 | 0.32 | 0.643 | 0.342 | 0.391 | 0.166 | 0.303 | 0.327 | 0.289 | 0.337 | 0.226 |
| *KIAA1328* | 18-37074791-G-A | 3_prime_UTR_variant | 0.26 | 0.154 | 0.181 | 0.178 | 0.047 | 0.184 | 0.198 | 0.002 | 0.162 | 0.167 | 0.116 | 0.15 | 0.106 |
| *KIAA1328* | 18-37084130-A-G | missense_variant | 0.36 | 0.318 | 0.321 | 0.321 | 0.642 | 0.34 | 0.392 | 0.17 | 0.278 | 0.324 | 0.291 | 0.337 | 0.225 |
| *REEP6* | 19-1495278-C-T | intron_variant | 0.307 | 0.246 | 0.282 | 0.121 | 0.252 | 0.273 | 0.332 | 0.402 | 0.314 | 0.338 | 0.264 | 0.294 | 0.371 |
| *REEP6* | 19-1495635-G-A | intron_variant | 0.367 | 0.308 | 0.337 | 0.148 | 0.486 | 0.326 | 0.376 | 0.402 | 0.389 | 0.378 | 0.34 | 0.354 | 0.394 |
| *REEP6* | 19-1496264-A-G | intron_variant | 0.373 | 0.306 | 0.342 | 0.169 | 0.511 | 0.333 | 0.381 | 0.403 | 0.393 | 0.384 | 0.358 | 0.359 | 0.4 |
| *REEP6* | 19-1496717-TGC-T | intron_variant | 0.313 | 0.224 | 0.278 | 0.105 | 0.173 | 0.277 | 0.304 | 0.341 | 0.283 | 0.329 | 0.281 | 0.277 | 0.341 |
| *REEP6* | 19-1497122-G-T | intron_variant | 0.393 | 0.296 | 0.323 | 0.322 | 0.644 | 0.35 | 0.348 | 0.396 | 0.383 | 0.389 | 0.354 | 0.351 | 0.372 |
| *PRX* | 19-40395104-G-C | missense_variant | 0.113 | 0.177 | 0.191 | 0.167 | 0.037 | 0.078 | 0.099 | 0.024 | 0.107 | 0.143 | 0.189 | 0.157 | 0.168 |
| *PRX* | 19-40395589-T-C | missense_variant | 0.333 | 0.413 | 0.407 | 0.275 | 0.32 | 0.416 | 0.332 | 0.132 | 0.266 | 0.346 | 0.515 | 0.363 | 0.25 |
| *PRX* | 19-40395697-A-G | synonymous_variant | 0.533 | 0.545 | 0.534 | 0.348 | 0.801 | 0.518 | 0.461 | 0.186 | 0.45 | 0.449 | 0.652 | 0.515 | 0.471 |
| *PRX* | 19-40395707-A-G | missense_variant | 0.533 | 0.545 | 0.533 | 0.347 | 0.8 | 0.519 | 0.461 | 0.186 | 0.449 | 0.449 | 0.653 | 0.515 | 0.471 |
| *PRX* | 19-40398695-G-A | missense_variant | 0.127 | 0.182 | 0.192 | 0.168 | 0.037 | 0.08 | 0.099 | 0.024 | 0.113 | 0.143 | 0.189 | 0.159 | 0.188 |
| *PRX* | 19-40403914-C-T | 5_prime_UTR_variant | 0.187 | 0.229 | 0.209 | 0.157 | 0.167 | 0.145 | 0.232 | 0.108 | 0.141 | 0.2 | 0.325 | 0.183 | 0.052 |
| *NCKAP5* | 2-132731828-A-G | synonymous_variant | 0.167 | 0.164 | 0.159 | 0.123 | 0.036 | 0.125 | 0.144 | 0.171 | 0.122 | 0.166 | 0.2 | 0.137 | 0.071 |
| *NCKAP5* | 2-132732028-A-G | synonymous_variant | 0.32 | 0.281 | 0.279 | 0.198 | 0.081 | 0.153 | 0.249 | 0.084 | 0.258 | 0.222 | 0.391 | 0.246 | 0.275 |
| *NCKAP5* | 2-132781887-A-T | intron_variant | 0.54 | 0.532 | 0.49 | 0.249 | 0.512 | 0.482 | 0.525 | 0.734 | 0.517 | 0.401 | 0.739 | 0.506 | 0.47 |
| *NCKAP5* | 2-132781993-G-A | synonymous_variant | 0.54 | 0.532 | 0.491 | 0.247 | 0.512 | 0.486 | 0.527 | 0.735 | 0.517 | 0.401 | 0.737 | 0.507 | 0.466 |
| *NCKAP5* | 2-132783534-T-A | missense_variant | 0.26 | 0.241 | 0.24 | 0.17 | 0.07 | 0.182 | 0.264 | 0.279 | 0.254 | 0.139 | 0.293 | 0.231 | 0.242 |
| *NCKAP5* | 2-132783871-T-C | synonymous_variant | 0.593 | 0.567 | 0.525 | 0.38 | 0.822 | 0.672 | 0.549 | 0.905 | 0.58 | 0.491 | 0.735 | 0.574 | 0.584 |
| *NCKAP5* | 2-132783881-A-G | missense_variant | 0.573 | 0.552 | 0.502 | 0.404 | 0.807 | 0.661 | 0.542 | 0.906 | 0.55 | 0.447 | 0.733 | 0.555 | 0.576 |
| *NCKAP5* | 2-132784002-C-T | missense_variant | 0.307 | 0.326 | 0.282 | 0.177 | 0.455 | 0.316 | 0.3 | 0.459 | 0.295 | 0.324 | 0.458 | 0.303 | 0.24 |
| *NCKAP5* | 2-132784441-T-C | synonymous_variant | 0.58 | 0.565 | 0.514 | 0.388 | 0.808 | 0.68 | 0.552 | 0.902 | 0.575 | 0.449 | 0.736 | 0.567 | 0.58 |
| *NCKAP5* | 2-132784972-G-A | synonymous_variant | 0.287 | 0.299 | 0.257 | 0.2 | 0.436 | 0.308 | 0.272 | 0.457 | 0.274 | 0.269 | 0.443 | 0.281 | 0.224 |
| *NCKAP5* | 2-132785001-C-T | missense_variant | 0.26 | 0.241 | 0.239 | 0.161 | 0.078 | 0.179 | 0.266 | 0.279 | 0.253 | 0.138 | 0.293 | 0.231 | 0.242 |
| *NCKAP5* | 2-132796715-A-G | synonymous_variant | 0.613 | 0.57 | 0.534 | 0.384 | 0.709 | 0.681 | 0.592 | 0.918 | 0.616 | 0.5 | 0.741 | 0.581 | 0.598 |
| *NCKAP5* | 2-132860647-A-G | intron_variant | 0.233 | 0.234 | 0.226 | 0.233 | 0.365 | 0.342 | 0.284 | 0.458 | 0.27 | 0.148 | 0.174 | 0.26 | 0.311 |
| *NCKAP5* | 2-133123852-G-A | intron_variant | 0.04 | 0.075 | 0.073 | 0.102 | 0.029 | 0.071 | 0.04 | 0.176 | 0.019 | 0.045 | 0.024 | 0.072 | 0.116 |
| *NCKAP5* | 2-133303147-T-C | intron_variant | 0.073 | 0.107 | 0.119 | 0.157 | 0.276 | 0.1 | 0.096 | 0.123 | 0.148 | 0.085 | 0.16 | 0.126 | 0.138 |
| *PLEKHA3* | 2-178493938-A-G | synonymous_variant | 0.18 | 0.174 | 0.146 | 0.482 | 0.204 | 0.177 | 0.196 | 0.628 | 0.206 | 0.127 | 0.227 | 0.192 | 0.307 |
| *PLEKHA3* | 2-178499182-AT-A | intron_variant | 0.027 | 0.002 | 0.12 | 0.12 | 0.158 | 0.127 | 0.128 | 0.123 | 0.102 | 0.124 | 0 | 0.141 | 0.154 |
| *PLEKHA3* | 2-178502359-A-G | missense_variant | 0.167 | 0.182 | 0.139 | 0.481 | 0.524 | 0.195 | 0.225 | 0.62 | 0.25 | 0.169 | 0.245 | 0.213 | 0.277 |
| *ATP6V1E2* | 2-46512018-A-G | 3_prime_UTR_variant | 0.44 | 0.428 | 0.397 | 0.215 | 0.373 | 0.514 | 0.428 | 0.613 | 0.441 | 0.441 | 0.308 | 0.421 | 0.342 |
| *KCNK12* | 2-47521153-A-G | synonymous_variant | 0.427 | 0.353 | 0.377 | 0.412 | 0.789 | 0.397 | 0.369 | 0.605 | 0.414 | 0.458 | 0.502 | 0.434 | 0.392 |
| *SLC12A8* | 3-125083833-A-G | 3_prime_UTR_variant | 0.507 | 0.537 | 0.517 | 0.268 | 0.785 | 0.62 | 0.431 | 0.577 | 0.431 | 0.609 | 0.527 | 0.533 | 0.507 |
| *SLC12A8* | 3-125084037-A-G | synonymous_variant | 0.513 | 0.537 | 0.516 | 0.268 | 0.784 | 0.618 | 0.431 | 0.578 | 0.431 | 0.607 | 0.524 | 0.533 | 0.506 |
| *SLC12A8* | 3-125084044-C-T | missense_variant | 0.513 | 0.532 | 0.516 | 0.268 | 0.784 | 0.618 | 0.43 | 0.577 | 0.429 | 0.607 | 0.526 | 0.532 | 0.493 |
| *SLC12A8* | 3-125135792-A-T | intron_variant | 0.3 | 0.353 | 0.364 | 0.323 | 0.348 | 0.284 | 0.289 | 0.041 | 0.316 | 0.307 | 0.367 | 0.316 | 0.242 |
| *SLC12A8* | 3-125177824-G-A | missense_variant | 0.047 | 0.04 | 0.051 | 0.166 | 0.01 | 0.115 | 0.035 | 0.015 | 0.036 | 0.055 | 0.217 | 0.044 | 0.051 |
| *SLC12A8* | 3-125178017-C-G | intron_variant | 1 | 0.993 | 0.999 | 0.117 | 0.882 | 0.99 | 1 | 1 | 0.992 | 1 | 1 | 0.99 | 0.999 |
| *NR2C2* | 3-15024278-G-A | intron_variant | 0.087 | 0.092 | 0.124 | 0.103 | 0.021 | 0.064 | 0.09 | 0.021 | 0.126 | 0.081 | 0.215 | 0.1 | 0.031 |
| *DAZL* | 3-16592157-C-T | intron_variant | 0.087 | 0.104 | 0.111 | 0.315 | 0.118 | 0.274 | 0.089 | 0.426 | 0.079 | 0.132 | 0.071 | 0.136 | 0.193 |
| *DAZL* | 3-16594586-TA-T | splice_region_variant | 0.247 | 0.274 | 0.362 | 0.118 | 0.374 | 0.287 | 0.41 | 0.245 | 0.364 | 0.401 | 0.399 | 0.366 | 0.323 |
| *DAZL* | 3-16595278-T-C | intron_variant | 0.087 | 0.104 | 0.114 | 0.309 | 0.118 | 0.266 | 0.089 | 0.423 | 0.08 | 0.132 | 0.071 | 0.139 | 0.191 |
| *DAZL* | 3-16596960-A-G | intron_variant | 0.087 | 0.1 | 0.113 | 0.314 | 0.119 | 0.275 | 0.09 | 0.427 | 0.075 | 0.132 | 0.071 | 0.138 | 0.194 |
| *DAZL* | 3-16597456-T-G | intron_variant | 0.087 | 0.104 | 0.113 | 0.313 | 0.119 | 0.275 | 0.089 | 0.427 | 0.073 | 0.132 | 0.071 | 0.138 | 0.193 |
| *DAZL* | 3-16598247-G-T | intron_variant | 0.58 | 0.565 | 0.571 | 0.251 | 0.589 | 0.488 | 0.635 | 0.32 | 0.558 | 0.557 | 0.55 | 0.554 | 0.475 |
| *DAZL* | 3-16598568-T-C | missense_variant | 0.147 | 0.134 | 0.137 | 0.16 | 0.021 | 0.084 | 0.145 | 0.059 | 0.13 | 0.161 | 0.296 | 0.119 | 0.057 |
| *EIF4G1* | 3-184321878-A-G | missense_variant | 0.747 | 0.731 | 0.745 | 0.253 | 0.957 | 0.819 | 0.828 | 0.649 | 0.821 | 0.695 | 0.492 | 0.763 | 0.672 |
| *EIF4G1* | 3-184326645-C-T | intron_variant | 0.113 | 0.104 | 0.122 | 0.114 | 0.02 | 0.103 | 0.104 | 0.019 | 0.079 | 0.165 | 0.008 | 0.101 | 0.105 |
| *EIF4G1* | 3-184328011-A-G | intron_variant | 0.747 | 0.734 | 0.746 | 0.258 | 0.935 | 0.817 | 0.828 | 0.711 | 0.822 | 0.694 | 0.488 | 0.764 | 0.675 |
| *EIF4G1* | 3-184328567-C-T | intron_variant | 0.727 | 0.734 | 0.745 | 0.254 | 0.957 | 0.818 | 0.828 | 0.624 | 0.82 | 0.695 | 0.491 | 0.761 | 0.668 |
| *EIF4G1* | 3-184328682-C-T | synonymous_variant | 0.28 | 0.249 | 0.248 | 0.211 | 0.053 | 0.135 | 0.243 | 0.038 | 0.279 | 0.136 | 0.268 | 0.225 | 0.278 |
| *SCN5A* | 3-38550915-A-G | synonymous_variant | 0.32 | 0.348 | 0.331 | 0.325 | 0.657 | 0.376 | 0.346 | 0.529 | 0.438 | 0.443 | 0.361 | 0.38 | 0.334 |
| *SCN5A* | 3-38557178-A-G | intron_variant | 0.04 | 0.09 | 0.056 | 0.225 | 0.124 | 0.075 | 0.076 | 0.281 | 0.118 | 0.122 | 0.009 | 0.086 | 0.086 |
| *SCN5A* | 3-38576589-A-G | intron_variant | 0.047 | 0.065 | 0.066 | 0.14 | 0.053 | 0.033 | 0.048 | 0 | 0.066 | 0.064 | 0.206 | 0.058 | 0.039 |
| *SCN5A* | 3-38580976-T-C | synonymous_variant | 0.92 | 0.898 | 0.89 | 0.11 | 0.885 | 0.934 | 0.861 | 0.999 | 0.894 | 0.924 | 0.787 | 0.895 | 0.942 |
| *SCN5A* | 3-38603929-T-C | missense_variant | 0.293 | 0.221 | 0.234 | 0.14 | 0.294 | 0.222 | 0.196 | 0.095 | 0.235 | 0.199 | 0.339 | 0.233 | 0.241 |
| *SCN5A* | 3-38604932-C-T | intron_variant | 0.273 | 0.182 | 0.199 | 0.109 | 0.168 | 0.195 | 0.154 | 0.09 | 0.179 | 0.176 | 0.166 | 0.19 | 0.206 |
| *SCN5A* | 3-38633221-T-C | synonymous_variant | 0.827 | 0.789 | 0.799 | 0.128 | 0.898 | 0.717 | 0.751 | 0.672 | 0.704 | 0.702 | 0.818 | 0.782 | 0.746 |
| *BEND3* | 6-107070192-G-A | synonymous_variant | 0.227 | 0.209 | 0.184 | 0.255 | 0.124 | 0.392 | 0.311 | 0.44 | 0.299 | 0.24 | 0.326 | 0.224 | 0.277 |
| *BEND3* | 6-107099236-A-AT | intron_variant | 0.18 | 0.177 | 0.19 | 0.189 | 0.291 | 0.093 | 0.128 | 0.001 | 0.063 | 0.251 | 0.108 | 0.164 | 0.058 |
| *BEND3* | 6-107099312-A-C | intron_variant | 0.387 | 0.413 | 0.364 | 0.261 | 0.103 | 0.625 | 0.511 | 0.481 | 0.509 | 0.429 | 0.397 | 0.388 | 0.432 |
| *NOTCH4* | 6-32196022-T-C | synonymous_variant | 0.08 | 0.055 | 0.055 | 0.102 | 0.085 | 0.039 | 0.049 | 0.053 | 0.071 | 0.021 | 0.156 | 0.06 | 0.094 |
| *NOTCH4* | 6-32197092-G-C | intron_variant | 0.38 | 0.351 | 0.278 | 0.201 | 0.197 | 0.293 | 0.479 | 0.179 | 0.437 | 0.292 | 0.323 | 0.278 | 0.362 |
| *NOTCH4* | 6-32197097-C-A | intron_variant | 0.373 | 0.313 | 0.256 | 0.221 | 0.133 | 0.286 | 0.477 | 0.179 | 0.43 | 0.262 | 0.283 | 0.261 | 0.361 |
| *NOTCH4* | 6-32198607-C-G | intron_variant | 0.047 | 0.065 | 0.066 | 0.141 | 0.036 | 0.067 | 0.151 | 0.207 | 0.07 | 0.029 | 0.042 | 0.076 | 0.062 |
| *NOTCH4* | 6-32200994-G-T | synonymous_variant | 0.08 | 0.109 | 0.144 | 0.124 | 0.089 | 0.046 | 0.083 | 0.021 | 0.044 | 0.115 | 0.073 | 0.118 | 0.078 |
| *NOTCH4* | 6-32202656-C-T | intron_variant | 0.553 | 0.507 | 0.48 | 0.17 | 0.371 | 0.41 | 0.65 | 0.343 | 0.588 | 0.452 | 0.577 | 0.475 | 0.583 |
| *NOTCH4* | 6-32203906-T-C | intron_variant | 0.553 | 0.505 | 0.482 | 0.169 | 0.381 | 0.409 | 0.651 | 0.422 | 0.565 | 0.451 | 0.576 | 0.48 | 0.589 |
| *NOTCH4* | 6-32204288-T-G | synonymous_variant | 0.907 | 0.883 | 0.86 | 0.138 | 0.975 | 0.948 | 0.971 | 0.998 | 0.943 | 0.869 | 0.928 | 0.892 | 0.976 |
| *NOTCH4* | 6-32210996-T-C | intron_variant | 0.553 | 0.51 | 0.483 | 0.167 | 0.381 | 0.41 | 0.65 | 0.422 | 0.569 | 0.452 | 0.581 | 0.48 | 0.589 |
| *NOTCH4* | 6-32212464-C-T | intron_variant | 0.127 | 0.107 | 0.119 | 0.185 | 0.224 | 0.135 | 0.179 | 0.304 | 0.099 | 0.056 | 0.077 | 0.143 | 0.097 |
| *NOTCH4* | 6-32212654-A-T | intron_variant | 0.047 | 0.062 | 0.066 | 0.141 | 0.036 | 0.067 | 0.151 | 0.207 | 0.07 | 0.029 | 0.042 | 0.076 | 0.062 |
| *NOTCH4* | 6-32215398-G-A | intron_variant | 0.453 | 0.4 | 0.393 | 0.192 | 0.211 | 0.322 | 0.541 | 0.201 | 0.458 | 0.375 | 0.292 | 0.372 | 0.426 |
| *NOTCH4* | 6-32216928-G-A | intron_variant | 0.82 | 0.781 | 0.727 | 0.206 | 0.864 | 0.691 | 0.933 | 0.902 | 0.883 | 0.631 | 0.71 | 0.772 | 0.877 |
| *NOTCH4* | 6-32219828-A-G | intron_variant | 0.547 | 0.537 | 0.553 | 0.157 | 0.522 | 0.517 | 0.71 | 0.695 | 0.58 | 0.486 | 0.608 | 0.561 | 0.593 |
| *NOTCH4* | 6-32219860-G-T | intron_variant | 0.067 | 0.095 | 0.139 | 0.121 | 0.091 | 0.047 | 0.078 | 0.018 | 0.049 | 0.107 | 0.06 | 0.112 | 0.076 |
| *NOTCH4* | 6-32220322-C-T | intron_variant | 0.393 | 0.291 | 0.251 | 0.202 | 0.129 | 0.245 | 0.453 | 0.134 | 0.338 | 0.289 | 0.232 | 0.252 | 0.234 |
| *NOTCH4* | 6-32220520-G-C | synonymous_variant | 0.46 | 0.338 | 0.301 | 0.201 | 0.212 | 0.281 | 0.502 | 0.265 | 0.398 | 0.312 | 0.38 | 0.312 | 0.333 |
| *NOTCH4* | 6-32220606-T-C | missense_variant | 0.54 | 0.45 | 0.448 | 0.24 | 0.301 | 0.329 | 0.583 | 0.208 | 0.442 | 0.424 | 0.457 | 0.428 | 0.406 |
| *NOTCH4* | 6-32220826-C-T | synonymous_variant | 0.467 | 0.343 | 0.304 | 0.199 | 0.214 | 0.282 | 0.502 | 0.187 | 0.398 | 0.309 | 0.398 | 0.311 | 0.329 |
| *NOTCH4* | 6-32220863-T-C | missense_variant | 0.467 | 0.343 | 0.303 | 0.198 | 0.214 | 0.282 | 0.502 | 0.187 | 0.397 | 0.309 | 0.396 | 0.311 | 0.329 |
| *NOTCH4* | 6-32220865-T-C | synonymous_variant | 0.467 | 0.343 | 0.303 | 0.198 | 0.214 | 0.282 | 0.501 | 0.266 | 0.397 | 0.309 | 0.395 | 0.313 | 0.333 |
| *NOTCH4* | 6-32221255-T-C | synonymous_variant | 0.467 | 0.341 | 0.304 | 0.198 | 0.215 | 0.283 | 0.502 | 0.266 | 0.399 | 0.31 | 0.398 | 0.314 | 0.334 |
| *NOTCH4* | 6-32222440-T-C | intron_variant | 0.28 | 0.316 | 0.348 | 0.172 | 0.377 | 0.301 | 0.397 | 0.52 | 0.379 | 0.227 | 0.33 | 0.372 | 0.399 |
| *NOTCH4* | 6-32222613-T-G | missense_variant | 0.287 | 0.328 | 0.382 | 0.122 | 0.376 | 0.277 | 0.378 | 0.504 | 0.304 | 0.307 | 0.328 | 0.372 | 0.375 |
| *NOTCH4* | 6-32222629-A-G | synonymous_variant | 0.373 | 0.388 | 0.39 | 0.154 | 0.465 | 0.349 | 0.452 | 0.527 | 0.407 | 0.236 | 0.415 | 0.416 | 0.465 |
| *NOTCH4* | 6-32222707-G-A | synonymous_variant | 0.12 | 0.119 | 0.086 | 0.212 | 0.298 | 0.113 | 0.077 | 0.238 | 0.112 | 0.086 | 0.029 | 0.123 | 0.205 |
| *NOTCH4* | 6-32222843-T-C | intron_variant | 0.34 | 0.358 | 0.304 | 0.256 | 0.475 | 0.336 | 0.414 | 0.56 | 0.366 | 0.223 | 0.281 | 0.345 | 0.467 |
| *NOTCH4* | 6-32223804-T-A | intron_variant | 0.34 | 0.284 | 0.242 | 0.182 | 0.321 | 0.31 | 0.425 | 0.385 | 0.336 | 0.134 | 0.329 | 0.259 | 0.207 |
| *NOTCH4* | 6-32223843-C-T | intron_variant | 0.293 | 0.269 | 0.244 | 0.111 | 0.245 | 0.216 | 0.355 | 0.253 | 0.306 | 0.185 | 0.175 | 0.254 | 0.276 |
| *NOTCH4* | 6-32223881-T-TAGC | inframe_insertion | 0.113 | 0.04 | 0.075 | 0.11 | 0.082 | 0.071 | 0.185 | 0.111 | 0.162 | 0.041 | 0.095 | 0.086 | 0.085 |
| *NOTCH4* | 6-32223881-T-TAGCAGC | inframe_insertion | 0.027 | 0.042 | 0.123 | 0.116 | 0.074 | 0.045 | 0.043 | 0.007 | 0.028 | 0.078 | 0.068 | 0.098 | 0.063 |
| *NOTCH4* | 6-32223881-TAGC-T | inframe_deletion | 0.2 | 0.209 | 0.248 | 0.284 | 0.532 | 0.352 | 0.281 | 0.353 | 0.42 | 0.193 | 0.322 | 0.289 | 0.398 |
| *NOTCH4* | 6-32223881-TAGCAGCAGCAGC-T | inframe_deletion | 0.033 | 0.035 | 0.088 | 0.189 | 0.078 | 0.121 | 0.276 | 0.155 | 0.148 | 0.081 | 0.126 | 0.113 | 0.118 |
| *NOTCH4* | 6-32223953-G-A | 5_prime_UTR_variant | 0.387 | 0.351 | 0.307 | 0.262 | 0.568 | 0.405 | 0.433 | 0.398 | 0.472 | 0.232 | 0.46 | 0.339 | 0.418 |
| *IP6K3* | 6-33723019-C-T | missense_variant | 0.353 | 0.445 | 0.44 | 0.382 | 0.166 | 0.587 | 0.461 | 0.821 | 0.428 | 0.591 | 0.51 | 0.461 | 0.705 |
| *IP6K3* | 6-33723020-A-G | synonymous_variant | 0.373 | 0.445 | 0.442 | 0.39 | 0.19 | 0.596 | 0.462 | 0.832 | 0.448 | 0.591 | 0.511 | 0.471 | 0.737 |
| *IP6K3* | 6-33728038-G-A | intron_variant | 0.287 | 0.219 | 0.194 | 0.116 | 0.093 | 0.151 | 0.178 | 0.081 | 0.188 | 0.134 | 0.119 | 0.169 | 0.077 |
| *IP6K3* | 6-33728354-C-T | intron_variant | 0.3 | 0.219 | 0.195 | 0.16 | 0.035 | 0.148 | 0.198 | 0.081 | 0.182 | 0.134 | 0.122 | 0.166 | 0.076 |
| *IP6K3* | 6-33735312-C-T | synonymous_variant | 0.153 | 0.157 | 0.15 | 0.118 | 0.092 | 0.083 | 0.109 | 0.032 | 0.14 | 0.108 | 0.205 | 0.124 | 0.062 |
| *IP6K3* | 6-33735453-G-A | synonymous_variant | 0.3 | 0.316 | 0.358 | 0.305 | 0.664 | 0.229 | 0.315 | 0.075 | 0.306 | 0.264 | 0.368 | 0.344 | 0.177 |
| *AGAP3* | 7-151115406-G-C | synonymous_variant | 0.713 | 0.652 | 0.677 | 0.306 | 0.852 | 0.801 | 0.786 | 0.982 | 0.739 | 0.758 | 0.632 | 0.717 | 0.702 |
| *AGAP3* | 7-151115652-G-C | intron_variant | 0.367 | 0.326 | 0.306 | 0.255 | 0.561 | 0.274 | 0.29 | 0.391 | 0.322 | 0.276 | 0.223 | 0.326 | 0.29 |
| *AGAP3* | 7-151115700-A-G | intron_variant | 0.36 | 0.331 | 0.314 | 0.262 | 0.576 | 0.283 | 0.299 | 0.415 | 0.334 | 0.283 | 0.223 | 0.336 | 0.296 |
| *AGAP3* | 7-151116890-T-TG | intron_variant | 0.32 | 0.291 | 0.267 | 0.138 | 0.13 | 0.152 | 0.279 | 0.194 | 0.254 | 0.257 | 0.179 | 0.257 | 0.251 |
| *AGAP3* | 7-151128752-G-A | intron_variant | 0.247 | 0.192 | 0.2 | 0.222 | 0.279 | 0.422 | 0.241 | 0.374 | 0.229 | 0.227 | 0.086 | 0.23 | 0.226 |
| *FBXW2* | 9-120787749-G-A | intron_variant | 0.807 | 0.786 | 0.738 | 0.193 | 0.824 | 0.859 | 0.788 | 0.545 | 0.81 | 0.703 | 0.695 | 0.749 | 0.821 |
| *PLPP7* | 9-131289981-G-C | 5_prime_UTR_variant | 0.313 | 0.393 | 0.395 | 0.176 | 0.219 | 0.322 | 0.318 | 0.505 | 0.288 | 0.443 | 0.426 | 0.381 | 0.451 |
| *PLPP7* | 9-131290501-T-C | intron_variant | 0.52 | 0.522 | 0.526 | 0.374 | 0.901 | 0.574 | 0.494 | 0.799 | 0.548 | 0.557 | 0.529 | 0.574 | 0.613 |
| *PLPP7* | 9-131290512-A-G | intron_variant | 0.307 | 0.398 | 0.399 | 0.112 | 0.486 | 0.357 | 0.322 | 0.511 | 0.316 | 0.447 | 0.425 | 0.405 | 0.457 |
| *PLPP7* | 9-131307992-C-T | missense_variant | 0.807 | 0.731 | 0.744 | 0.293 | 0.452 | 0.6 | 0.75 | 0.702 | 0.773 | 0.775 | 0.679 | 0.716 | 0.701 |
| *PLPP7* | 9-131308116-T-C | synonymous_variant | 0.807 | 0.731 | 0.745 | 0.258 | 0.487 | 0.604 | 0.75 | 0.704 | 0.774 | 0.775 | 0.681 | 0.718 | 0.703 |
| *PLPP7* | 9-131346846-G-C | missense_variant | 0.213 | 0.187 | 0.181 | 0.102 | 0.079 | 0.161 | 0.107 | 0.249 | 0.155 | 0.183 | 0.252 | 0.159 | 0.142 |

* Maximum of allele frequency difference between NFE vs. non-NFE populations in gnomAD

Abbreviations: MCI, mild cognitive impairment; AF, Allele frequency; nfe, Non-Finnish European; afr, African American; amr: Admixed American; asj; Ashkenazi Jewish; eas, East Asian; mid, Middle Eastern; fin, Finnish European; ami, Amish; asa, South Asian; oth, Remaining.

#### **Supplementary Table 6: Rare structural variants (SV) (AF_gnomAD-NFE_ < 0.05) overlapped with synaptic genes previously identified as enriched with LoF/missense SNVs in MCI vs external controls.**

| **Gene** | **Variant position** | **Type** | **Length (bp)** | **No. of individuals** | **Consequence** | **gnomAD SV v4.1** | | | | | | | **% Overlap** |
| --- | --- | --- | --- | --- | --- | --- | --- | --- | --- | --- | --- | --- | --- |
|  |  |  |  |  |  | **Variant** | **NFE** | | | **Global** | | |  |
|  |  |  |  |  |  |  | **AF** | **OR (95% CI)** | **adjusted-P** | **AF** | **OR (95% CI)** | **adjusted-P** |  |
| *PIK3C2G* | chr12:18570336-18572160 | DEL | 1,824 | 1 | Intronic | 12:18569977-18572160 | 1.3E-02 | 0.53  (0.07 – 3.77) | *ns* | 3.5E-02 | 0.18  (0.03 – 1.31) | *ns* | 100% (83.6%) |
| *ATP6V1E2* | chr2:46524048-46524172 | DEL | 124 | 1 | Intronic | 2:46523959-46524172 | 8.49E-05 | 79.05  (9.18 – 680.73) | 2.78E-04 | 3.85E-03 | 1.74  (0.24 – 12.42) | *ns* | 100% (58.2%) |
| *TNKS1BP1* | chr11:57304632-57304828 | DEL | 196 | 1 | Intronic | 11:57304601-57304828 | 1.12E-02 | 0.60  (0.08 – 4.30) | *ns* | 7.17E-03 | 0.93  (0.13 – 6.65) | *ns* | 100% (86.3%) |
| *PRKRA* | chr2:178444501-178447508 | DEL | 3,007 | 14 | Loss of function | 2:178444501-178447508 | 2.00E-04 | 513.86  (224.34 – 1177.02) | 1.04E-48 | 3.41E-04 | 302.20  (159.74 – 571.74) | 2.12E-68 | 100% (100%) |

* Non-Finnish European subpopulation in gnomAD SV v4.1.

^†^ All populations in gnomAD SV v4.1.

^‡^ % Overlap of the SV found in the MCI subgroup (% Overlap of the SV found in gnomAD SVs).

Abbreviations: NFE, Non-Finnish European; AF, Allele Frequency; DEL, Deletion; SV, Structural variant; bp, base-pair.

#### **Supplementary Table 7: A list of gene showing protein expression significantly changed in Alzheimer’s Disease (AD) vs. controls in ≥ 5 studies of human brain tissue.^1^**

**Down-regulated proteins**

| *VGF* | *RPH3A* | *CORO1A* | *ACTN2* | *HOMER1* | *DNM1L* | *SYT12* | *STXBP1* |
| --- | --- | --- | --- | --- | --- | --- | --- |
| *AP3D1* | *OTUB1* | *GNAI1* | *SUCLA2* | *SH3GL1* | *AP2A1* | *ATP2A2* | *CAP2* |
| *FARSB* | *GRIA2* | *PPP2R1A* | *SEPTIN5* | *YWHAQ* | *DLG4* | *DNAJC6* | *PLXNA1* |
| *SUCLG1* | *SYNPO* | *DPP6* | *SNAP25* | *CA4* | *OLFM1* | *GLS* | *LRPPRC* |
| *MAP2* | *STX1A* | *GAS7* | *PPME1* | *DBNL* | *MAPRE3* | *HPCA* | *RTN4* |
| *PREP* | *CYFIP2* | *OXCT1* | *PGM2L1* | *IRGQ* | *AP2M1* | *AP3B2* | *BAIAP2* |
| *KIF5C* | *NDUFA10* | *PAK1* | *SGIP1* | *SH3GLB2* | *TLN2* | *ANKS1B* | *LRRC57* |
| *MAP2K4* | *NRN1* | *YWHAZ* | *AP2A2* | *DCLK1* | *DLAT* | *DPYSL4* | *EPB41L3* |
| *FARSA* | *HSPA4L* | *PDHX* | *SYNGAP1* | *SYNJ1* | *CRMP1* | *DBN1* | *SYT1* |
| *ACTN1* | *ATL1* | *BSN* | *GNAZ* | *KIAA0513* | *NECAP1* | *PHF24* | *RALA* |
| *EPN1* | *NDUFV1* | *NUMBL* | *PCLO* | *SIRPA* | *SLC2A3* | *DBT* | *TIMM44* |
| *FBXL16* | *SCAI* | *SLC4A10* | *CYRIA* | *HGS* | *SGTB* | *SLC25A27* | *SLIRP* |
| *PLCB1* | *VPS35* | *AP2B1* | *ATP1A3* | *CD47* | *CLTC* | *PRKCG* | *GSK3A* |
| *NNT* | *OGDH* | *ADAP1* | *OGDHL* | *ABR* | *AP1G1* | *ARHGEF2* | *CADPS* |
| *CASKIN1* | *CORO2B* | *DIRAS2* | *DMXL2* | *GDAP1* | *GRPEL1* | *IARS2* | *LINGO1* |
| *NFS1* | *OPA1* | *PPM1H* | *RTN1* | *SEPTIN11* | *SLC30A3* | *CRIP2* | *NCKAP1* |
| *NDUFA5* | *PDCD6IP* | *SV2B* | *TWF2* | *KLC2* | *CTNND1* | *MGLL* | *RABEP1* |
| *RGS7* | *VTA1* | *NECTIN1* | *PRKACA* | *RAB3B* | *PALM* | *CAMK2A* | *IPO7* |
| *NDUFS6* | *ATP2B2* | *CADM3* | *ATP2B1* | *NAPB* | *PACSIN1* | *PDHA1* | *SPTBN2* |
| *TRAP1* | *TUBB3* | *IDH3A* | *PDHB* | *MYO5A* | *ABI2* | *CAMK4* | *CEP170B* |
| *CHCHD6* | *DLG1* | *DLG3* | *DYNC1LI1* | *GNL1* | *NDUFB9* | *NECAB2* | *PCDH1* |
| *PHACTR1* | *PIP5K1C* | *PLXNA4* | *PRKCE* | *RTN3* | *SLC25A22* | *TAGLN3* | *WDR7* |
| *AUH* | *CRAT* | *KRAS* | *PPP2R5E* | *PSD3* | *ROCK2* | *UQCRB* | *VSNL1* |
| *ACTR3B* | *GRIA3* | *GSK3B* | *HMOX2* | *LRRC40* | *NCALD* | *PLCL2* | *ASAP1* |
| *CACNG3* | *MRPS35* | *NPTX2* | *NRBP1* | *OLFM3* | *OSBPL8* | *PCSK1* | *PPEF1* |
| *SHANK2* | *USP14* | *ATP6V1H* | *DNM1* | *COPS4* | *DUSP3* | *COX6B1* | *ATP6V1D* |
| *ATP1A2* | *ATP1B1* | *DMTN* | *GDA* | *GPHN* | *HSPD1* | *STX1B* | *IMMT* |
| *PHB* | *TPPP* | *TUBA4A* | *TUBB* | *CDC42* | *STAM* | *AP1B1* | *AP3M2* |
| *CADM2* | *CPLX2* | *DNM3* | *LONP1* | *NDUFA9* | *PDK3* | *PPP2CA* | *PPP5C* |
| *RASAL1* | *SLC8A2* | *SYNGR1* | *TOM1L2* | *WARS1* | *WASF1* | *ATP5PO* | *CPNE5* |
| *CYC1* | *DIRAS1* | *FXYD6* | *NTM* | *RHOB* | *SAR1A* | *YARS1* | *NECAB1* |
| *GFRA2* | *SYT7* | *TBC1D24* | *CPLX1* | *GRM2* | *ABLIM2* | *AGK* | *DYNLL2* |
| *EHD3* | *FAM162A* | *GRIN1* | *LGALSL* | *MARK1* | *PRKRA* | *SARS2* | *TTC7B* |
| *GRK2* | *MMUT* | *RIMS1* | *SNX30* | *ADAM11* | *AGPAT1* | *CDH10* | *CXADR* |
| *EIF4G1* | *GRM3* | *NF1* | *P62158* | *PPP2R5B* | *SRGAP2* | *SST* | *STK24* |
| *EEF1B2* | *BASP1* | *COX5A* | *COX5B* | *LDHA* | *FKBP1A* | *HPCAL4* | *PACS1* |
| *CYRIB* | *RPLP0* | *ACTN4* | *CDK5* | *DLD* | *DSTN* | *GNB1* | *HSPE1* |
| *IDH3B* | *NAPG* | *NDUFS1* | *NDUFS3* | *PPP3CB* | *PURA* | *SEPTIN7* | *SLC25A3* |
| *SNAP91* | *SYN1* | *VAMP2* | *CKB* | *CLTA* | *CLTB* | *EEF1G* | *FABP3* |
| *PDXP* | *SLC25A11* | *AP1M1* | *TPT1* | *LY6H* | *NDUFS4* | *AFG3L2* | *AGAP2* |
| *ARPC1A* | *ATP2B3* | *DPP10* | *ELOB* | *EPHA4* | *HPRT1* | *LETM1* | *NCAN* |
| *NDUFB10* | *NDUFB7* | *NDUFC2* | *NDUFS2* | *NIT1* | *NT5DC3* | *PPP1CB* | *RELCH* |
| *RPS16* | *RPS4X* | *SLC17A7* | *SLC7A5* | *TBC1D10B* | *TTYH1* | *UQCRQ* | *WASF3* |
| *ACTR2* | *AIFM1* | *APEH* | *CALR* | *CAMKV* | *CTNNA2* | *DCTN1* | *DPYSL5* |
| *GNAI2* | *LANCL2* | *PDE2A* | *PPM1E* | *PTK2B* | *RAP2B* | *SDHA* | *SLC25A4* |
| *SLC25A5* | *SYNGR3* | *ECPAS* | *MT-ATP8* | *BPHL* | *DLGAP1* | *FXYD7* | *ZADH2* |
| *AIMP1* | *AVL9* | *C2CD2L* | *CDC42BPB* | *CRACDL* | *DLG2* | *EFR3B* | *GPR158* |
| *GRIN2B* | *GSTK1* | *IARS1* | *OAT* | *PICK1* | *PPP1CA* | *PPP2R5D* | *RAB3GAP2* |
| *RHEB* | *SDR39U1* | *SHMT2* | *SLC25A46* | *STRAP* | *SUB1* | *TMX4* | *ACSS1* |
| *CD200* | *IPO5* | *SCFD1* | *NRAS* | *ADGRB2* | *AKAP5* | *ASTN1* | *CALB1* |
| *CAMKK2* | *CIRBP* | *COA6* | *DGKZ* | *ELAVL2* | *EVL* | *EXOC7* | *FGF12* |
| *G3BP2* | *GJB6* | *GNG4* | *ISCU* | *KIAA1549L* | *MICOS10* | *MLIP* | *MOB4* |
| *NGEF* | *NUMB* | *PAK3* | *PCP4* | *PRUNE2* | *R3HDM2* | *RAB3GAP1* | *RAPGEF4* |
| *RASAL2* | *RDH13* | *REEP1* | *REEP5* | *SHISA7* | *SLC27A4* | *SLC39A10* | *STRN4* |
| *STXBP5L* | *BCAP29* | *FDX2* | *GGA3* | *MFF* | *MT-ND4* | *PRMT1* | *SYNE1* |
| *TPP2* | *UQCR10* |  |  |  |  |  |  |

**Up-regulated proteins**

| *GFAP* | *APP* | *HSPB1* | *CD44* | *CLU* | *CAPG* | *PADI2* | *PLCD1* |
| --- | --- | --- | --- | --- | --- | --- | --- |
| *ANXA5* | *GJA1* | *C4A* | *APOE* | *ANXA1* | *AHNAK* | *PRDX6* | *BAG3* |
| *MAOB* | *PBXIP1* | *GPNMB* | *PLEC* | *GNG12* | *S100A6* | *C4B; C4B_2* | *GMPR* |
| *ADIRF* | *APCS* | *HTRA1* | *ANXA2* | *PRDX1* | *CSRP1* | *DBI* | *FABP7* |
| *CAPS* | *HSPB8* | *MAPT* | *VIM* | *CSTB* | *EZR* | *MDK* | *SMOC1* |
| *AQP4* | *FABP5* | *ACAN* | *MTAP* | *PGAM2* | *AK1* | *GSTP1* | *PSAT1* |
| *C3* | *FHL1* | *MSN* | *ICAM1* | *PGD* | *SPR* | *UGP2* | *HP* |
| *CHGA* | *AMPD2* | *ITGB1* | *ANO6* | *ENO1* | *PAFAH1B3* | *CP* | *SERPINA1* |
| *LTA4H* | *ADD3* | *CAPN2* | *CAPNS1* | *GYG1* | *SELENBP1* | *SYNM* | *PNPO* |
| *PGM2* | *SCIN* | *TPP1* | *FLNA* | *PLSCR4* | *NPC2* | *PLIN3* | *S100A11* |
| *SQSTM1* | *GAPDH* | *CLIC1* | *FLT1* | *COTL1* | *FGA* | *FGB* | *TNC* |
| *MAPK1* | *NEFM* | *PKM* | *QDPR* | *SEPTIN2* | *TKT* | *FTL* | *LHPP* |
| *S100A1* | *LCP1* | *ALAD* | *BBOX1* | *H1-2* | *H4C1-C16* | *LGALS1* | *HSPB6* |
| *PLCD3* | *MACROD1* | *CD109* | *HEBP2* | *FGG* | *C1QC* | *OLFML3* | *PHGDH* |
| *CTSD* | *DKK3* | *HMGB1* | *ALDH1L1* | *DDAH2* | *LGALS3* | *H1-0* | *NTN1* |
| *SDC4* | *CA2* | *HEPACAM* | *NEFH* | *NEFL* | *SRI* | *MAP4* | *AGRN* |
| *A2M* | *DPYSL3* | *ESD* | *GNA13* | *GRHPR* | *MACROH2A1* | *PAICS* | *PGLS* |
| *TJP2* | *ATP8A1* | *CLIC4* | *G6PD* | *NDRG4* | *PSME1* | *PTRHD1* | *NUCKS1* |
| *QPRT* | *S100B* | *CD99* | *GYG2* | *H2AX* | *H3-3A; H3-3B* | *ITIH4* | *LAMP2* |
| *P08107* | *SPP1* | *TPD52L1* | *SERPING1* | *CNDP2* | *BLVRB* | *AKR1B1* | *RNH1* |
| *SCRN1* | *COL25A1* | *APRT* | *ARRB1* | *ENTPD2* | *FBXO2* | *TPPP3* | *SERPINA3* |
| *ALDH9A1* | *GSN* | *PYGB* | *RUVBL2* | *S100A9* | *SH3BGRL* | *NPEPPS* | *PRDX5* |
| *SOD1* | *TMPO* | *ASAH1* | *HLA-DRA* | *HPX* | *ATG3* | *ERLIN2* | *ADH5* |
| *ANXA4* | *BDH2* | *PGM3* | *PPT1* | *RDX* | *SLC9A3R1* | *TAGLN2* | *CRYL1* |
| *SORBS1* | *STOM* | *TMEM30A* | *CTHRC1* | *SAMHD1* | *GLRX* | *IGKC* | *ITGAV* |
| *NAMPT* | *NUDT5* | *PLXNB1* | *RNPEP* | *RPS6KA2* | *DST* | *RAB27B* | *RANGAP1* |
| *TPD52L2* | *IGHG1* | *ANP32B* | *GC* | *GPD1* | *H2AZ1* | *IGFBP5* | *ISYNA1* |
| *ITGA7* | *P05067a* | *PEA15* | *CBR1* | *CPNE6* | *HNRNPM* | *GLUD1* | *ABHD14B* |
| *SLIT2* | *PTN* | *AKR7A2* | *NAGK* | *TXNDC17* | *PLP1* | *IMPA1* | *LDHB* |
| *LMNA* | *MTHFD1* | *PEBP1* | *SRSF1* | *INA* | *LANCL1* | *GSPT1* | *TKFC* |
| *FGF1* | *FN3KRP* | *LRP1* | *RPS25* | *SPOCK2* | *DDX3X* | *SOD2* | *FAH* |
| *H1-4* | *H2AZ2* | *HSPA2* | *IDH1* | *LLGL1* | *MAP2K2* | *MLC1* | *SNRPD2* |
| *TCEAL3* | *HBA1-A2* | *HINT1* | *HSD17B4* | *KCTD12* | *PPP1R1B* | *SLC1A3* | *TLN1* |
| *SPOCK3* | *ETF1* | *S100A10* | *SFRP1* | *KRT9* | *DNPEP* | *EDF1* | *FGF2* |
| *GPX1* | *MPST* | *ISOC1* | *LAMP1* | *UBA6* | *ALB* | *ARL6IP4* | *ASPH* |
| *CDKN1B* | *CLDN10* | *CZIB* | *DEK* | *GSTZ1* | *H1-3* | *HDGF* | *HMGA1* |
| *HTATSF1* | *ITGB8* | *LCMT1* | *NT5C* | *NTN3* | *P0CG05* | *PLXDC2* | *PTGR2* |
| *RBKS* | *RPS6KA5* | *SLC39A12* | *UBE2I* | *CAVIN2* | *CFB* | *NEK7* | *PLIN4* |
| *SLC4A1* | *SYT2* |  |  |  |  |  |  |

**Either up- or down-regulated***

| *VCAN* | *IDH2* | *UCHL1* | *RAP1GDS1* | *HAPLN2* | *RAB3A* | *ORM1* | *YWHAB* |
| --- | --- | --- | --- | --- | --- | --- | --- |
| *IDH3G* | *UBA1* | *HSP90AA1* | *THY1* | *GAP43* | *WDR1* | *ATP1B2* | *MAP2K1* |
| *PDIA3* | *PPIA* | *ARHGDIA* | *OLA1* | *FASN* | *HP1BP3* | *PFN2* | *GABRA1* |
| *MTX2* | *RAC1* | *SEPTIN3* | *TBCB* | *MECP2* | *TANC2* | *HDHD2* | *VPS26A* |
| *AHCYL1* | *DDX1* | *DPYSL2* | *NDRG2* | *ACAT2* | *IGHA1* | *MTCH2* | *RIDA* |
| *CRKL* | *GPM6B* | *ORM2* | *RFTN1* | *RPLP1* | *PGAM1* | *HSP90AB1* | *PTPA* |
| *UBE2M* | *ATP5PF* | *ABAT* | *RPS3* | *TPI1* | *BCL2L13* | *ACYP2* | *EEF1A2* |
| *EIF4H* | *PLPBP* | *PSMA2* | *VPS29* | *KPNA3* | *SBDS* | *SNRPD3* | *TAX1BP3* |
| *RPS12* | *SMS* | *FSCN1* | *GPI* | *NCDN* | *CNRIP1* | *AASDHPPT* | *AKR1A1* |
| *ARL3* | *GNB2* | *S100A13* | *ANK2* | *CHCHD3* | *COPS6* | *SRSF7* | *AARS1* |
| *MAPRE2* | *PAFAH1B1* | *PRNP* | *PSMA1* | *RPL5* | *STUM* | *TARS1* | *OSBPL1A* |
| *BCAP31* | *NRGN* | *ACADM* | *FKBP2* | *SAMM50* | *SUCLG2* | *ARMT1* | *ATP5IF1* |
| *ATP5MK* | *DMXL1* | *RPS15* | *SNRPA* |  |  |  |  |

* The direction of protein expression changes varies across studies, with some reporting upregulation and others downregulation.

*Note*: Proteins are ranked from left to right and top to bottom according to the number of bulk tissue studies. Genes enriched for the mild cognitive decline (MCI) subgroup are highlighted in blue

#### **Supplementary Table 8: Gene ontology (GO) enrichment analysis for Molecular Function (MF) terms in the MCI gene burden set.**

| **Pathway ID** | **Description** | **Gene Ratio** | **Background Ratio** | **Odd Ratio** | ***P* value** | **Adjusted *P* value** | **Gene** |
| --- | --- | --- | --- | --- | --- | --- | --- |
| GO:0015079 | potassium ion transmembrane transporter activity | 4/49 | 158/18522 | 10.33 | 8.01E-04 | 0.04 | SLC12A8/ATP12A/KCNK13/KCNK12 |
| GO:0022841 | potassium ion leak channel activity | 2/49 | 16/18522 | 49.22 | 8.03E-04 | 0.04 | KCNK13/KCNK12 |
| GO:0022840 | leak channel activity | 2/49 | 19/18522 | 41.44 | 1.14E-03 | 0.04 | KCNK13/KCNK12 |

#### **Supplementary Table 9: Genes enriched for high-confidence loss-of-function (LoF) and missense variants (CADD ≥ 20) associated with very-mild-to-mild tinnitus (0 ≤ THI ≤ 36, n = 103).**

**Genes enriched with LoF variants**

| Gene | Number of variants | Number of individuals | gnomADg (NFE)* | | | gnomADg (global)^†^ | | |
| --- | --- | --- | --- | --- | --- | --- | --- | --- |
|  |  |  | **OR (95% CI)**^‡^ | **Adjusted-*P*** | **EF** | **OR (95% CI)** | **Adjusted-*P*** | **EF** |
| *PRKRA* | 3 | 6 | 13.64 (6 - 31.01) | 9.41E-08 | 0.93 | 14.06 (6.24 - 31.66) | 3.77E-08 | 0.93 |
| *CFAP157* | 1 | 2 | 158.29 (28.83 - 869.07) | 1.18E-06 | 0.99 | 351.35 (64 - 1928.95) | 3.21E-09 | 1 |
| *ANKDD1B* | 2 | 2 | 17.57 (6.39 - 48.33) | 5.91E-06 | 0.94 | 0.4 (0.15 - 1.07) | *ns* | -1.5 |
| *HK3* | 1 | 2 | 79.12 (16.7 - 374.89) | 7.72E-06 | 0.99 | 100.32 (22.66 - 444.24) | 2.71E-07 | 0.99 |
| *H4C2* | 2 | 3 | 25.6 (7.86 - 83.35) | 1.55E-05 | 0.96 | 33.9 (10.6 - 108.42) | 6.07E-07 | 0.97 |
| *SERPINA9* | 1 | 4 | 15.2 (5.52 - 41.84) | 2.93E-05 | 0.93 | 1.11 (0.41 - 2.99) | *ns* | 0.1 |
| *TRMT9B* | 1 | 2 | 20.98 (6.47 - 68.08) | 8.46E-05 | 0.95 | 12.74 (4.03 - 40.24) | 3.08E-03 | 0.92 |
| *TTC6* | 1 | 2 | 23.42 (5.53 - 99.15) | 3.89E-03 | 0.96 | 32.64 (7.85 - 135.62) | 3.43E-04 | 0.97 |
| *CAPN12* | 1 | 2 | 21.82 (5.17 - 92.04) | 5.72E-03 | 0.95 | 36 (8.64 - 150.09) | 1.84E-04 | 0.97 |
| *IQCE* | 2 | 4 | 8.27 (3.05 - 22.41) | 7.04E-03 | 0.88 | 13.43 (4.97 - 36.29) | 6.42E-05 | 0.93 |
| *ERMARD* | 2 | 3 | 11.01 (3.47 - 34.95) | 1.00E-02 | 0.91 | 18.11 (5.73 - 57.22) | 1.69E-04 | 0.94 |
| *CFAP99* | 1 | 4 | 7.12 (2.62 - 19.37) | 2.54E-02 | 0.86 | 6.67 (2.47 - 18.03) | 3.88E-02 | 0.85 |

**Genes enriched with missense variants**

| Gene | Number of variants | Number of individuals | gnomADg (NFE)* | | | gnomADg (global)^†^ | | |
| --- | --- | --- | --- | --- | --- | --- | --- | --- |
|  |  |  | **OR (95% CI)**^‡^ | **Adjusted-*P*** | **EF** | **OR (95% CI)** | **Adjusted-*P*** | **EF** |
| *EIF4G1* | 1 | 6 | 169.06 (61.92 - 461.57) | 4.82E-20 | 0.99 | 112.05 (46.77 - 268.43) | 1.22E-22 | 0.99 |
| *BEND3* | 1 | 5 | 1602.51 (186.38 - 13778.44) | 6.34E-08 | 1 | 3553.66 (413.31 - 30554.2) | 3.36E-10 | 1 |
| *LRP12* | 1 | 3 | 56.11 (16.32 - 192.96) | 5.85E-07 | 0.98 | 70.56 (21.36 - 233.06) | 1.03E-08 | 0.99 |
| *EP400* | 1 | 4 | 1249.8 (139.08 - 11231.03) | 6.92E-07 | 1 | 685.11 (170.17 - 2758.28) | 1.41E-16 | 1 |
| *ATP6V1E2* | 1 | 6 | 12.23 (5.35 - 27.97) | 1.04E-05 | 0.92 | 15.1 (6.65 - 34.29) | 3.14E-07 | 0.93 |
| *FBXW2* | 1 | 3 | 927.98 (96.12 - 8958.95) | 1.24E-05 | 1 | 227.99 (61.28 - 848.27) | 1.96E-12 | 1 |
| *VIL1* | 1 | 3 | 36.68 (11.01 - 122.14) | 1.56E-05 | 0.97 | 42.32 (13.09 - 136.79) | 1.39E-06 | 0.98 |
| *C17orf99* | 1 | 5 | 14.99 (6.05 - 37.14) | 1.77E-05 | 0.93 | 2 (0.82 - 4.86) | *ns* | 0.5 |
| *FAM53A* | 1 | 2 | 90.43 (18.67 - 437.95) | 7.75E-05 | 0.99 | 0.81 (0.2 - 3.27) | *ns* | -0.23 |
| *ADH1B* | 1 | 7 | 8.44 (3.93 - 18.1) | 1.54E-04 | 0.88 | 10.89 (5.09 - 23.26) | 2.53E-06 | 0.91 |
| *FREM1* | 3 | 9 | 4.98 (2.8 - 8.86) | 1.63E-04 | 0.8 | 4.64 (2.62 - 8.23) | 5.48E-04 | 0.78 |
| *CABP4* | 1 | 3 | 20.7 (6.38 - 67.08) | 1.57E-03 | 0.95 | 17.16 (5.41 - 54.38) | 4.85E-03 | 0.94 |
| *UBR1* | 1 | 3 | 20.29 (6.26 - 65.71) | 1.84E-03 | 0.95 | 1.78 (0.57 - 5.56) | *ns* | 0.44 |
| *SMPD3* | 1 | 2 | 37.23 (8.55 - 162.18) | 5.16E-03 | 0.97 | 36.02 (8.64 - 150.18) | 3.06E-03 | 0.97 |
| *ATXN2L* | 2 | 11 | 4.33 (2.37 - 7.91) | 6.83E-03 | 0.77 | 5.38 (2.95 - 9.81) | 1.47E-04 | 0.81 |
| *CDSN* | 2 | 9 | 4.64 (2.46 - 8.72) | 6.95E-03 | 0.78 | 7.35 (3.91 - 13.81) | 2.01E-06 | 0.86 |
| *THAP5* | 1 | 2 | 35.16 (8.11 - 152.51) | 7.04E-03 | 0.97 | 26.5 (6.42 - 109.49) | 2.11E-02 | 0.96 |
| *SLC16A3* | 1 | 3 | 17.02 (5.29 - 54.83) | 7.22E-03 | 0.94 | 26.8 (8.39 - 85.57) | 1.01E-04 | 0.96 |
| *ATF6B* | 3 | 12 | 3.96 (2.23 - 7.03) | 9.67E-03 | 0.75 | 6.43 (3.62 - 11.41) | 7.39E-07 | 0.84 |
| *STARD9* | 1 | 3 | 16.16 (5.02 - 51.97) | 1.08E-02 | 0.94 | 0.45 (0.14 - 1.41) | *ns* | -1.21 |
| *MTCH2* | 1 | 3 | 19.53 (5.57 - 68.48) | 1.22E-02 | 0.95 | 15.74 (4.84 - 51.25) | 1.67E-02 | 0.94 |
| *HCAR1* | 1 | 3 | 10.82 (3.96 - 29.58) | 1.24E-02 | 0.91 | 13.05 (4.81 - 35.42) | 1.64E-03 | 0.92 |
| *ABL2* | 2 | 12 | 3.71 (2.13 - 6.47) | 1.33E-02 | 0.73 | 5.59 (3.21 - 9.73) | 4.24E-06 | 0.82 |
| *TMEM214* | 1 | 3 | 15.63 (4.86 - 50.22) | 1.39E-02 | 0.94 | 26.8 (8.39 - 85.57) | 1.01E-04 | 0.96 |
| *CCDC183* | 2 | 4 | 5.2 (2.57 - 10.53) | 1.65E-02 | 0.81 | 7.43 (3.68 - 15.01) | 8.19E-05 | 0.87 |
| *ZNF302* | 1 | 3 | 15.13 (4.71 - 48.59) | 1.77E-02 | 0.93 | 2.22 (0.71 - 6.94) | *ns* | 0.55 |
| *OR3A2* | 1 | 3 | 14.67 (4.57 - 47.05) | 2.24E-02 | 0.93 | 24.62 (7.72 - 78.48) | 2.17E-04 | 0.96 |
| *TRANK1* | 1 | 2 | 27.51 (6.44 - 117.44) | 2.70E-02 | 0.96 | 14.17 (3.47 - 57.84) | *ns* | 0.93 |
| *VEPH1* | 2 | 5 | 7.58 (3.1 - 18.5) | 3.06E-02 | 0.87 | 8.7 (3.58 - 21.13) | 6.26E-03 | 0.89 |
| *IFITM10* | 1 | 8 | 4.98 (2.44 - 10.16) | 3.49E-02 | 0.8 | 7.2 (3.54 - 14.65) | 1.80E-04 | 0.86 |
| *LAMB3* | 2 | 15 | 3.09 (1.87 - 5.1) | 3.78E-02 | 0.68 | 2.69 (1.63 - 4.43) | *ns* | 0.63 |

* Non-Finnish European subpopulation in gnomAD genome v4.1.

^†^ All populations in gnomAD genome v4.1.

^‡^ Odds ratio with 95% confidence interval.

Abbreviations: CADD, Combined Annotation Dependent Depletion; NFE, Non-Finnish European; OR, Odds Ratio, CI, Confidence Interval; EF, Etiological Fraction; *ns*, non-significant.

Note: Genes are ordered by adjusted *P*-values (Bonferroni correction) of gnomADg (NFE).

#### **Supplementary Table 10: Genes enriched for high-confidence loss-of-function (LoF) and missense variants (CADD ≥ 20) associated with moderate tinnitus (38 ≤ THI ≤ 56, n = 101).**

**Genes enriched with LoF variants**

| Gene | Number of variants | Number of individuals | gnomADg (NFE)* | | | gnomADg (global)^†^ | | |
| --- | --- | --- | --- | --- | --- | --- | --- | --- |
|  |  |  | **OR (95% CI)**^‡^ | **Adjusted-*P*** | **EF** | **OR (95% CI)** | **Adjusted-*P*** | **EF** |
| *IGFN1* | 1 | 4 | 81.48 (27 - 245.89) | 1.26E-12 | 0.99 | 51.64 (18.55 - 143.77) | 9.44E-12 | 0.98 |
| *AIRE* | 1 | 3 | 29.48 (8.97 - 96.9) | 5.45E-06 | 0.97 | 25.71 (8.06 - 82) | 8.93E-06 | 0.96 |
| *CHRNA3* | 1 | 2 | 80.7 (17.03 - 382.41) | 6.88E-06 | 0.99 | 119.4 (26.55 - 536.95) | 9.80E-08 | 0.99 |
| *PRKRA* | 3 | 5 | 11.58 (4.73 - 28.35) | 1.82E-05 | 0.91 | 11.93 (4.91 - 28.97) | 9.50E-06 | 0.92 |
| *RSPH1* | 1 | 2 | 58.7 (12.93 - 266.52) | 2.87E-05 | 0.98 | 79.59 (18.35 - 345.29) | 1.09E-06 | 0.99 |
| *PIK3C2G* | 2 | 5 | 7.69 (3.15 - 18.77) | 1.62E-03 | 0.87 | 11.13 (4.58 - 27.08) | 2.32E-05 | 0.91 |
| *ENTPD4* | 1 | 2 | 23.9 (5.65 - 101.18) | 3.53E-03 | 0.96 | 34.93 (8.39 - 145.39) | 2.26E-04 | 0.97 |

**Genes enriched with missense variants**

| Gene | Number of variants | Number of individuals | gnomADg (NFE)* | | | gnomADg (global)^†^ | | |
| --- | --- | --- | --- | --- | --- | --- | --- | --- |
|  |  |  | **OR (95% CI)**^‡^ | **Adjusted-*P*** | **EF** | **OR (95% CI)** | **Adjusted-*P*** | **EF** |
| *EIF4G1* | 1 | 10 | 293.51 (123.21 - 699.2) | 3.99E-34 | 1 | 194.53 (95.37 - 396.76) | 4.76E-44 | 0.99 |
| *BEND3* | 1 | 12 | 4068.69 (526.41 - 31447.49) | 5.87E-12 | 1 | 9022.55 (1167.35 - 69735.92) | 9.17E-15 | 1 |
| *ADM* | 1 | 8 | 15.09 (7.33 - 31.07) | 6.43E-10 | 0.93 | 1.86 (0.92 - 3.77) | *ns* | 0.46 |
| *DLX4* | 1 | 5 | 27.64 (10.96 - 69.69) | 7.13E-09 | 0.96 | 26.36 (10.68 - 65.08) | 4.59E-09 | 0.96 |
| *EIF3F* | 1 | 3 | 69.49 (19.81 - 243.68) | 1.24E-07 | 0.99 | 107.9 (31.81 - 365.99) | 2.08E-10 | 0.99 |
| *RNF113B* | 1 | 3 | 60.82 (17.58 - 210.37) | 3.10E-07 | 0.98 | 0.89 (0.28 - 2.78) | *ns* | -0.12 |
| *FBXW2* | 1 | 4 | 1268.55 (141.15 - 11400.46) | 6.39E-07 | 1 | 311.66 (95.19 - 1020.4) | 8.33E-18 | 1 |
| *GDF7* | 1 | 3 | 44.21 (13.13 - 148.89) | 3.43E-06 | 0.98 | 1.13 (0.36 - 3.54) | *ns* | 0.12 |
| *WWC2* | 1 | 4 | 23.28 (8.36 - 64.81) | 6.04E-06 | 0.96 | 38.58 (13.98 - 106.5) | 6.35E-09 | 0.97 |
| *DCAF5* | 1 | 4 | 22.09 (7.95 - 61.4) | 1.05E-05 | 0.95 | 45.92 (16.56 - 127.37) | 6.96E-10 | 0.98 |
| *EP400* | 1 | 3 | 951.48 (98.55 - 9186.67) | 1.09E-05 | 1 | 521.58 (115.98 - 2345.55) | 1.23E-12 | 1 |
| *MTCH2* | 1 | 4 | 26.7 (8.71 - 81.82) | 3.23E-05 | 0.96 | 21.52 (7.64 - 60.65) | 2.29E-05 | 0.95 |
| *GAS2L2* | 1 | 2 | 64.55 (14.05 - 296.46) | 3.01E-04 | 0.98 | 1.56 (0.39 - 6.3) | *ns* | 0.36 |
| *AGAP3* | 2 | 7 | 7.87 (3.69 - 16.77) | 3.19E-04 | 0.87 | 9.24 (4.35 - 19.59) | 2.45E-05 | 0.89 |
| *SLC12A8* | 1 | 4 | 15.68 (5.7 - 43.19) | 3.58E-04 | 0.94 | 17.73 (6.51 - 48.28) | 6.59E-05 | 0.94 |
| *OVCH1* | 1 | 3 | 24.32 (7.46 - 79.27) | 4.27E-04 | 0.96 | 0.34 (0.11 - 1.05) | *ns* | -1.98 |
| *SCPEP1* | 1 | 2 | 645.73 (58.32 - 7150.17) | 4.76E-04 | 1 | 716.21 (100.39 - 5109.59) | 1.95E-07 | 1 |
| *DIP2A* | 2 | 5 | 11.15 (4.55 - 27.34) | 4.89E-04 | 0.91 | 0.49 (0.2 - 1.19) | *ns* | -1.04 |
| *CYP4V2* | 1 | 3 | 21.61 (6.66 - 70.12) | 1.10E-03 | 0.95 | 5.01 (1.6 - 15.74) | *ns* | 0.8 |
| *KLKB1* | 1 | 3 | 21.61 (6.66 - 70.12) | 1.10E-03 | 0.95 | 5.01 (1.6 - 15.74) | *ns* | 0.8 |
| *ZFP2* | 1 | 3 | 20.69 (6.39 - 67.04) | 1.56E-03 | 0.95 | 0.34 (0.11 - 1.07) | *ns* | -1.91 |
| *PRX* | 1 | 2 | 46.1 (10.41 - 204.16) | 1.61E-03 | 0.98 | 0.48 (0.12 - 1.95) | *ns* | -1.07 |
| *COL8A2* | 1 | 3 | 19.38 (6 - 62.67) | 2.62E-03 | 0.95 | 1.62 (0.52 - 5.06) | *ns* | 0.38 |
| *MAP3K20* | 1 | 2 | 40.34 (9.21 - 176.59) | 3.29E-03 | 0.98 | 0.67 (0.17 - 2.72) | *ns* | -0.48 |
| *VPS11* | 1 | 3 | 17.05 (5.3 - 54.91) | 7.11E-03 | 0.94 | 18.42 (5.81 - 58.44) | 2.69E-03 | 0.95 |
| *OR4N2* | 1 | 2 | 33.98 (7.86 - 146.85) | 8.36E-03 | 0.97 | 0.41 (0.1 - 1.66) | *ns* | -1.43 |
| *OR5AU1* | 1 | 2 | 32.28 (7.5 - 139.02) | 1.11E-02 | 0.97 | 0.32 (0.08 - 1.29) | *ns* | -2.11 |
| *ATF6* | 1 | 2 | 32.24 (7.49 - 138.85) | 1.12E-02 | 0.97 | 64.97 (15.18 - 278.13) | 6.59E-05 | 0.98 |
| *CEP55* | 1 | 3 | 15.69 (4.88 - 50.39) | 1.35E-02 | 0.94 | 0.49 (0.16 - 1.54) | *ns* | -1.03 |
| *ITGAX* | 1 | 3 | 15.44 (4.81 - 49.57) | 1.52E-02 | 0.94 | 10.42 (3.31 - 32.86) | *ns* | 0.9 |
| *IGSF9B* | 1 | 4 | 10.41 (3.81 - 28.44) | 1.77E-02 | 0.9 | 1.28 (0.47 - 3.44) | *ns* | 0.22 |
| *NDUFAF6* | 1 | 5 | 7.89 (3.21 - 19.37) | 2.33E-02 | 0.87 | 9.42 (3.86 - 23.01) | 3.07E-03 | 0.89 |
| *EVL* | 1 | 2 | 28.07 (6.57 - 119.84) | 2.40E-02 | 0.96 | 0.29 (0.07 - 1.16) | *ns* | -2.47 |
| *NDUFV3* | 1 | 7 | 5.7 (2.67 - 12.19) | 2.58E-02 | 0.82 | 7.77 (3.64 - 16.58) | 4.09E-04 | 0.87 |
| *PPP1R7* | 1 | 2 | 26.9 (6.32 - 114.58) | 3.03E-02 | 0.96 | 46.2 (10.98 - 194.35) | 6.07E-04 | 0.98 |

* Non-Finnish European subpopulation in gnomAD genome v4.1.

^†^ All populations in gnomAD genome v4.1.

^‡^ Odds ratio with 95% confidence interval.

Abbreviations: CADD, Combined Annotation Dependent Depletion; NFE, Non-Finnish European; OR, Odds Ratio, CI, Confidence Interval; EF, Etiological Fraction; *ns*, non-significant.

Note: Genes are ordered by adjusted *P*-values (Bonferroni correction) of gnomADg (NFE).

#### **Supplementary Table 11: Genes enriched for high-confidence loss-of-function (LoF) and missense variants (CADD ≥ 20) associated with severe tinnitus (58 ≤ THI ≤ 76, n = 58).**

**Genes enriched with LoF variants**

| Gene | Number of variants | Number of individuals | gnomADg (NFE)* | | | gnomADg (global)^†^ | | |
| --- | --- | --- | --- | --- | --- | --- | --- | --- |
|  |  |  | **OR (95% CI)**^‡^ | **Adjusted-*P*** | **EF** | **OR (95% CI)** | **Adjusted-*P*** | **EF** |
| *ALDH3A1* | 1 | 2 | 125.83 (26.89 - 588.8) | 1.12E-07 | 0.99 | 61.29 (14.65 - 256.4) | 2.36E-06 | 0.98 |
| *CD36* | 1 | 2 | 70.76 (16.08 - 311.28) | 2.38E-06 | 0.99 | 1.17 (0.29 - 4.73) | *ns* | 0.14 |
| *CDHR4* | 1 | 4 | 16.52 (6.01 - 45.41) | 7.40E-06 | 0.94 | 11.2 (4.12 - 30.51) | 3.08E-04 | 0.91 |
| *HOXD12* | 1 | 2 | 45.29 (10.6 - 193.45) | 3.60E-05 | 0.98 | 96.65 (22.67 - 412.01) | 8.76E-08 | 0.99 |
| *HSD17B13* | 1 | 2 | 36.52 (8.64 - 154.42) | 1.36E-04 | 0.97 | 28.88 (7.02 - 118.73) | 4.25E-04 | 0.97 |
| *ENDOG* | 1 | 2 | 14.26 (4.47 - 45.5) | 9.77E-04 | 0.93 | 12.4 (3.92 - 39.24) | 2.50E-03 | 0.92 |
| *UPK2* | 1 | 2 | 20.96 (5.05 - 86.98) | 3.80E-03 | 0.95 | 35.39 (8.58 - 146) | 1.11E-04 | 0.97 |
| *FKBP7* | 1 | 3 | 11.25 (3.54 - 35.81) | 5.65E-03 | 0.91 | 19.09 (6.02 - 60.6) | 7.59E-05 | 0.95 |
| *PXMP2* | 2 | 3 | 11 (3.48 - 34.74) | 5.94E-03 | 0.91 | 11.8 (3.76 - 37.07) | 3.21E-03 | 0.92 |

**Genes enriched with missense variants**

| Gene | Number of variants | Number of individuals | gnomADg (NFE)* | | | gnomADg (global)^†^ | | |
| --- | --- | --- | --- | --- | --- | --- | --- | --- |
|  |  |  | **OR (95% CI)**^‡^ | **Adjusted-*P*** | **EF** | **OR (95% CI)** | **Adjusted-*P*** | **EF** |
| *EIF4G1* | 1 | 6 | 307.38 (111.71 - 845.79) | 3.36E-25 | 1 | 203.72 (84.28 - 492.45) | 8.86E-29 | 1 |
| *OR2AG1* | 2 | 3 | 74.48 (31.5 - 176.14) | 2.35E-19 | 0.99 | 0.68 (0.3 - 1.53) | *ns* | -0.47 |
| *BEND3* | 1 | 9 | 5418.59 (680.54 - 43143.94) | 1.12E-12 | 1 | 12016.01 (1509.14 - 95673.22) | 1.72E-15 | 1 |
| *NCKAP5* | 1 | 2 | 125.8 (26.88 - 588.67) | 2.02E-06 | 0.99 | 8.68 (2.13 - 35.28) | *ns* | 0.88 |
| *DNHD1* | 1 | 2 | 102.99 (22.57 - 469.89) | 5.31E-06 | 0.99 | 2.48 (0.61 - 10.07) | *ns* | 0.6 |
| *SLC26A6* | 1 | 4 | 20.19 (7.32 - 55.67) | 1.55E-05 | 0.95 | 14.67 (5.38 - 39.99) | 3.75E-04 | 0.93 |
| *AGAP3* | 1 | 2 | 79.39 (17.84 - 353.31) | 2.28E-05 | 0.99 | 61.66 (14.73 - 258.19) | 4.13E-05 | 0.98 |
| *EP400* | 1 | 2 | 1107.28 (99.7 - 12297.9) | 2.82E-05 | 1 | 606.98 (110.08 - 3346.99) | 4.61E-10 | 1 |
| *FBXW2* | 1 | 2 | 1101.63 (99.19 - 12235.16) | 2.89E-05 | 1 | 270.65 (57.84 - 1266.49) | 2.77E-09 | 1 |
| *LGALS12* | 1 | 2 | 75.5 (17.07 - 333.93) | 2.93E-05 | 0.99 | 1.29 (0.32 - 5.24) | *ns* | 0.23 |
| *PON3* | 1 | 2 | 62.88 (14.42 - 274.14) | 8.67E-05 | 0.98 | 4.58 (1.13 - 18.56) | *ns* | 0.78 |
| *ITGA6* | 1 | 2 | 56.62 (13.08 - 245.04) | 1.64E-04 | 0.98 | 0.69 (0.17 - 2.79) | *ns* | -0.45 |
| *ANK3* | 1 | 3 | 24.81 (7.7 - 80) | 1.86E-04 | 0.96 | 36.17 (11.31 - 115.67) | 3.54E-06 | 0.97 |
| *SLIT3* | 1 | 2 | 53.94 (12.5 - 232.71) | 2.20E-04 | 0.98 | 0.54 (0.13 - 2.2) | *ns* | -0.84 |
| *WWC3* | 1 | 4 | 7.11 (3.45 - 14.67) | 2.59E-04 | 0.86 | 12.37 (6 - 25.47) | 2.19E-08 | 0.92 |
| *MASP1* | 1 | 4 | 13.45 (4.91 - 36.88) | 1.08E-03 | 0.93 | 20.68 (7.57 - 56.5) | 8.55E-06 | 0.95 |
| *PSKH1* | 1 | 2 | 36.52 (8.64 - 154.42) | 2.45E-03 | 0.97 | 26.16 (6.37 - 107.39) | 1.45E-02 | 0.96 |
| *DOCK9* | 1 | 3 | 18.02 (5.63 - 57.71) | 2.76E-03 | 0.94 | 19.79 (6.23 - 62.82) | 1.00E-03 | 0.95 |
| *PLEKHA3* | 1 | 2 | 30.6 (7.29 - 128.48) | 7.26E-03 | 0.97 | 15.79 (3.87 - 64.48) | *ns* | 0.94 |
| *BRMS1L* | 1 | 2 | 27.62 (6.6 - 115.53) | 1.35E-02 | 0.96 | 39.28 (9.5 - 162.4) | 9.81E-04 | 0.97 |
| *PBX3* | 1 | 5 | 6.56 (2.87 - 14.99) | 1.98E-02 | 0.85 | 8.85 (3.88 - 20.18) | 5.34E-04 | 0.89 |
| *CYP4V2* | 1 | 2 | 25.15 (6.03 - 104.91) | 2.36E-02 | 0.96 | 5.84 (1.44 - 23.69) | *ns* | 0.83 |
| *KLKB1* | 1 | 2 | 25.15 (6.03 - 104.91) | 2.36E-02 | 0.96 | 5.84 (1.44 - 23.69) | *ns* | 0.83 |
| *HAGH* | 1 | 3 | 13.36 (4.19 - 42.61) | 2.88E-02 | 0.93 | 13.91 (4.39 - 44.04) | 1.86E-02 | 0.93 |
| *OR7G1* | 1 | 8 | 5 (2.43 - 10.29) | 2.97E-02 | 0.8 | 7.2 (3.51 - 14.8) | 1.88E-04 | 0.86 |
| *P2RY13* | 1 | 5 | 7.32 (2.97 - 18.03) | 3.70E-02 | 0.86 | 10.74 (4.37 - 26.42) | 5.66E-04 | 0.91 |
| *HKDC1* | 4 | 18 | 2.63 (1.7 - 4.08) | 3.76E-02 | 0.62 | 3.63 (2.34 - 5.63) | 1.94E-05 | 0.72 |
| *C8orf48* | 1 | 2 | 23.1 (5.55 - 96.14) | 3.89E-02 | 0.96 | 39.27 (9.5 - 162.36) | 9.83E-04 | 0.97 |
| *RAP1GDS1* | 1 | 4 | 9.08 (3.32 - 24.81) | 4.14E-02 | 0.89 | 9.17 (3.37 - 24.94) | 3.50E-02 | 0.89 |
| *SLIT2* | 1 | 5 | 7.19 (2.92 - 17.71) | 4.38E-02 | 0.86 | 10.96 (4.46 - 26.94) | 4.51E-04 | 0.91 |
| *CFAP45* | 1 | 7 | 5.33 (2.47 - 11.48) | 4.76E-02 | 0.81 | 1.38 (0.64 - 2.96) | *ns* | 0.27 |

* Non-Finnish European subpopulation in gnomAD genome v4.1.

^†^ All populations in gnomAD genome v4.1.

^‡^ Odds ratio with 95% confidence interval.

Abbreviations: CADD, Combined Annotation Dependent Depletion; NFE, Non-Finnish European; OR, Odds Ratio, CI, Confidence Interval; EF, Etiological Fraction; *ns*, non-significant.

Note: Genes are ordered by adjusted *P*-values (Bonferroni correction) of gnomADg (NFE).

#### **Supplementary Table 12: Genes enriched for high-confidence loss-of-function (LoF) and missense variants (CADD ≥ 20) associated with catastrophic tinnitus (78 ≤ THI ≤ 100, n = 32).**

**Genes enriched with LoF variants**

| Gene | Number of variants | Number of individuals | gnomADg (NFE)* | | | gnomADg (global)^†^ | | |
| --- | --- | --- | --- | --- | --- | --- | --- | --- |
|  |  |  | **OR (95% CI)**^‡^ | **Adjusted-*P*** | **EF** | **OR (95% CI)** | **Adjusted-*P*** | **EF** |
| *GRAMD1A* | 1 | 4 | 30.9 (11.08 - 86.19) | 4.04E-09 | 0.97 | 52.42 (18.85 - 145.74) | 2.35E-12 | 0.98 |
| *MYO15B* | 1 | 3 | 22.46 (6.96 - 72.42) | 1.39E-05 | 0.96 | 24.84 (7.75 - 79.62) | 4.74E-06 | 0.96 |
| *ERMARD* | 1 | 2 | 24.19 (5.82 - 100.47) | 8.45E-04 | 0.96 | 40.17 (9.71 - 166.19) | 2.51E-05 | 0.98 |
| *TOMM20L* | 1 | 2 | 23.64 (5.69 - 98.16) | 9.71E-04 | 0.96 | 26.55 (6.44 - 109.37) | 4.13E-04 | 0.96 |
| *ESF1* | 1 | 3 | 12.5 (3.9 - 40.1) | 1.58E-03 | 0.92 | 20.61 (6.43 - 65.99) | 2.55E-05 | 0.95 |
| *MYBPC2* | 1 | 2 | 15.5 (3.75 - 64.02) | 1.11E-02 | 0.94 | 20.04 (4.87 - 82.41) | 2.37E-03 | 0.95 |

**Genes enriched with missense variants**

| Gene | Number of variants | Number of individuals | gnomADg (NFE)* | | | gnomADg (global)^†^ | | |
| --- | --- | --- | --- | --- | --- | --- | --- | --- |
|  |  |  | **OR (95% CI)**^‡^ | **Adjusted-*P*** | **EF** | **OR (95% CI)** | **Adjusted-*P*** | **EF** |
| *BEND3* | 1 | 5 | 5459.41 (628.23 - 47443.34) | 8.85E-12 | 1 | 12106.53 (1393.13 - 105207.33) | 2.24E-14 | 1 |
| *USP34* | 1 | 2 | 54.78 (12.93 - 232.03) | 7.84E-05 | 0.98 | 65 (15.6 - 270.86) | 1.42E-05 | 0.98 |
| *CEP250* | 1 | 2 | 53.74 (12.69 - 227.65) | 9.06E-05 | 0.98 | 68.88 (16.51 - 287.43) | 9.15E-06 | 0.99 |
| *GALNS* | 1 | 3 | 22.31 (6.92 - 71.94) | 2.88E-04 | 0.96 | 41.16 (12.79 - 132.44) | 6.52E-07 | 0.98 |
| *EVC2* | 1 | 2 | 43.27 (10.29 - 181.96) | 3.91E-04 | 0.98 | 5.74 (1.4 - 23.5) | *ns* | 0.83 |
| *MTCH2* | 1 | 2 | 42.63 (9.49 - 191.51) | 1.41E-03 | 0.98 | 34.36 (8.13 - 145.18) | 2.16E-03 | 0.97 |
| *ZFHX4* | 2 | 8 | 3.84 (2.24 - 6.59) | 1.46E-03 | 0.74 | 3.11 (1.82 - 5.34) | 5.18E-02 | 0.68 |
| *ZBTB47* | 1 | 2 | 35.27 (8.43 - 147.53) | 1.52E-03 | 0.97 | 49.16 (11.85 - 203.9) | 1.15E-04 | 0.98 |
| *ARHGEF12* | 1 | 3 | 17.3 (5.38 - 55.64) | 2.48E-03 | 0.94 | 19.05 (5.95 - 61) | 9.91E-04 | 0.95 |
| *GPRC5C* | 1 | 2 | 32.52 (7.79 - 135.8) | 2.58E-03 | 0.97 | 33.48 (8.11 - 138.23) | 1.74E-03 | 0.97 |
| *MAML2* | 2 | 9 | 4.72 (2.47 - 9.01) | 3.68E-03 | 0.79 | 6.97 (3.65 - 13.3) | 5.63E-06 | 0.86 |
| *ACOT9* | 1 | 5 | 5.71 (2.72 - 12.01) | 6.16E-03 | 0.82 | 6.02 (2.87 - 12.65) | 3.01E-03 | 0.83 |
| *ANKRD52* | 1 | 2 | 27.38 (6.58 - 113.94) | 7.72E-03 | 0.96 | 40.51 (9.79 - 167.62) | 4.64E-04 | 0.98 |
| *FITM2* | 1 | 6 | 6.96 (3 - 16.17) | 9.27E-03 | 0.86 | 10.56 (4.55 - 24.51) | 5.93E-05 | 0.91 |
| *OR2B11* | 1 | 2 | 25.05 (6.03 - 104.1) | 1.34E-02 | 0.96 | 3.43 (0.84 - 14.04) | *ns* | 0.71 |
| *TJP2* | 1 | 2 | 24.19 (5.82 - 100.46) | 1.66E-02 | 0.96 | 2.47 (0.6 - 10.11) | *ns* | 0.6 |
| *KCNK12* | 1 | 2 | 23.11 (5.57 - 95.92) | 2.19E-02 | 0.96 | 22.52 (5.47 - 92.69) | 2.29E-02 | 0.96 |
| *USP24* | 1 | 3 | 12.03 (3.75 - 38.58) | 4.12E-02 | 0.92 | 15.62 (4.88 - 49.96) | 5.18E-03 | 0.94 |

* Non-Finnish European subpopulation in gnomAD genome v4.1.

^†^ All populations in gnomAD genome v4.1.

^‡^ Odds ratio with 95% confidence interval.

Abbreviations: CADD, Combined Annotation Dependent Depletion; NFE, Non-Finnish European; OR, Odds Ratio, CI, Confidence Interval; EF, Etiological Fraction; *ns*, non-significant.

Note: Genes are ordered by adjusted *P*-values (Bonferroni correction) of gnomADg (NFE).

#### **Supplementary Table 13: Genes enriched for high-confidence loss-of-function (LoF) and missense variants (CADD ≥ 20) associated with mild hyperacusis (0 ≤ GÜF ≤ 10, n = 104).**

**Genes enriched with LoF variants**

| Gene | Number of variants | Number of individuals | gnomADg (NFE)* | | | gnomADg (global)^†^ | | |
| --- | --- | --- | --- | --- | --- | --- | --- | --- |
|  |  |  | **OR (95% CI)**^‡^ | **Adjusted-*P*** | **EF** | **OR (95% CI)** | **Adjusted-*P*** | **EF** |
| *HK3* | 1 | 3 | 118.11 (31.11 - 448.36) | 5.40E-10 | 0.99 | 149.75 (42.71 - 525.02) | 1.15E-12 | 0.99 |
| *RSPH1* | 1 | 3 | 85.9 (23.79 - 310.17) | 2.42E-09 | 0.99 | 116.48 (34.05 - 398.46) | 7.80E-12 | 0.99 |
| *PRKRA* | 3 | 6 | 13.51 (5.94 - 30.71) | 1.17E-07 | 0.93 | 13.92 (6.18 - 31.35) | 4.73E-08 | 0.93 |
| *KIR3DL3* | 1 | 2 | 89.21 (18.42 - 432.01) | 5.48E-06 | 0.99 | 197.23 (40.73 - 955.1) | 1.18E-08 | 0.99 |
| *ANKDD1B* | 2 | 2 | 17.4 (6.33 - 47.86) | 7.07E-06 | 0.94 | 0.4 (0.15 - 1.06) | *ns* | -1.52 |
| *IRAK2* | 1 | 2 | 41.77 (9.49 - 183.81) | 1.81E-04 | 0.98 | 60.45 (14.16 - 258.07) | 6.95E-06 | 0.98 |
| *IGFN1* | 1 | 2 | 39.16 (8.95 - 171.39) | 2.56E-04 | 0.97 | 24.82 (6.02 - 102.37) | 2.04E-03 | 0.96 |
| *ALMS1* | 1 | 2 | 38.95 (8.9 - 170.47) | 2.65E-04 | 0.97 | 44.57 (10.6 - 187.46) | 5.05E-05 | 0.98 |
| *C11orf16* | 1 | 2 | 29.85 (6.95 - 128.12) | 1.12E-03 | 0.97 | 0.28 (0.07 - 1.14) | *ns* | -2.52 |
| *GRAMD1A* | 1 | 4 | 9.09 (3.33 - 24.79) | 3.71E-03 | 0.89 | 15.42 (5.67 - 41.92) | 1.90E-05 | 0.94 |
| *SERPINA9* | 1 | 3 | 11.23 (3.52 - 35.83) | 9.92E-03 | 0.91 | 0.82 (0.26 - 2.57) | *ns* | -0.22 |
| *AIRE* | 1 | 2 | 18.98 (4.53 - 79.63) | 1.31E-02 | 0.95 | 16.55 (4.05 - 67.74) | 2.16E-02 | 0.94 |

**Genes enriched with missense variants**

| Gene | Number of variants | Number of individuals | gnomADg (NFE)* | | | gnomADg (global)^†^ | | |
| --- | --- | --- | --- | --- | --- | --- | --- | --- |
|  |  |  | **OR (95% CI)**^‡^ | **Adjusted-*P*** | **EF** | **OR (95% CI)** | **Adjusted-*P*** | **EF** |
| *EIF4G1* | 1 | 4 | 110.5 (34.9 - 349.89) | 4.48E-12 | 0.99 | 73.23 (25.87 - 207.34) | 2.23E-12 | 0.99 |
| *BEND3* | 1 | 5 | 1586.72 (184.55 - 13642.1) | 6.88E-08 | 1 | 3518.65 (409.26 - 30251.85) | 3.67E-10 | 1 |
| *PRX* | 1 | 3 | 67.46 (19.24 - 236.53) | 1.70E-07 | 0.99 | 0.71 (0.23 - 2.22) | *ns* | -0.41 |
| *CEP250* | 1 | 4 | 32.67 (11.55 - 92.37) | 1.77E-07 | 0.97 | 41.87 (15.12 - 115.94) | 2.41E-09 | 0.98 |
| *RNF113B* | 1 | 3 | 59.04 (17.07 - 204.17) | 4.26E-07 | 0.98 | 0.86 (0.28 - 2.7) | *ns* | -0.16 |
| *EP400* | 1 | 3 | 923.63 (95.67 - 8916.68) | 1.30E-05 | 1 | 506.31 (112.61 - 2276.48) | 1.70E-12 | 1 |
| *B4GALT7* | 1 | 4 | 20.74 (7.47 - 57.56) | 2.12E-05 | 0.95 | 32.28 (11.74 - 88.77) | 6.08E-08 | 0.97 |
| *CEP55* | 1 | 4 | 20.4 (7.35 - 56.61) | 2.51E-05 | 0.95 | 0.64 (0.24 - 1.72) | *ns* | -0.56 |
| *MTCH2* | 1 | 4 | 25.91 (8.46 - 79.39) | 4.42E-05 | 0.96 | 20.89 (7.41 - 58.85) | 3.23E-05 | 0.95 |
| *ADM* | 1 | 6 | 10.87 (4.76 - 24.8) | 5.31E-05 | 0.91 | 1.34 (0.59 - 3.02) | *ns* | 0.25 |
| *FAM53A* | 1 | 2 | 89.55 (18.49 - 433.67) | 8.47E-05 | 0.99 | 0.8 (0.2 - 3.24) | *ns* | -0.24 |
| *HELZ2* | 1 | 3 | 28.63 (8.71 - 94.09) | 1.19E-04 | 0.97 | 32.77 (10.21 - 105.14) | 1.62E-05 | 0.97 |
| *FREM1* | 3 | 9 | 4.93 (2.77 - 8.77) | 2.01E-04 | 0.8 | 4.59 (2.59 - 8.15) | 6.70E-04 | 0.78 |
| *SFRP5* | 1 | 2 | 69.6 (14.95 - 324.11) | 2.34E-04 | 0.99 | 99.34 (22.44 - 439.87) | 5.00E-06 | 0.99 |
| *SCPEP1* | 1 | 2 | 626.92 (56.62 - 6941.1) | 5.50E-04 | 1 | 695.35 (97.48 - 4960.06) | 2.41E-07 | 1 |
| *DCDC1* | 1 | 3 | 23.03 (7.07 - 74.96) | 6.88E-04 | 0.96 | 0.4 (0.13 - 1.25) | *ns* | -1.5 |
| *IQGAP3* | 1 | 2 | 48.21 (10.81 - 214.99) | 1.36E-03 | 0.98 | 0.42 (0.1 - 1.7) | *ns* | -1.37 |
| *SNIP1* | 1 | 2 | 48.2 (10.81 - 214.95) | 1.36E-03 | 0.98 | 73.16 (16.93 - 316.09) | 3.25E-05 | 0.99 |
| *MAP3K20* | 1 | 2 | 39.16 (8.95 - 171.42) | 4.06E-03 | 0.97 | 0.65 (0.16 - 2.64) | *ns* | -0.53 |
| *ATP12A* | 3 | 14 | 3.42 (2.08 - 5.63) | 5.00E-03 | 0.71 | 3.11 (1.89 - 5.12) | 2.76E-02 | 0.68 |
| *SMPD3* | 1 | 2 | 36.87 (8.46 - 160.59) | 5.62E-03 | 0.97 | 35.67 (8.56 - 148.71) | 3.34E-03 | 0.97 |
| *LRP12* | 1 | 2 | 36.86 (8.46 - 160.58) | 5.62E-03 | 0.97 | 46.36 (11.01 - 195.24) | 6.16E-04 | 0.98 |
| *SH3GLB2* | 1 | 4 | 11.59 (4.23 - 31.74) | 6.74E-03 | 0.91 | 18.96 (6.96 - 51.66) | 3.20E-05 | 0.95 |
| *ANOS1* | 1 | 3 | 7.34 (3.22 - 16.73) | 7.57E-03 | 0.86 | 11.21 (4.94 - 25.47) | 2.79E-05 | 0.91 |
| *SCN9A* | 2 | 4 | 5.44 (2.69 - 11.02) | 9.13E-03 | 0.82 | 1.33 (0.66 - 2.67) | *ns* | 0.25 |
| *VPS11* | 1 | 3 | 16.55 (5.14 - 53.29) | 9.17E-03 | 0.94 | 17.88 (5.64 - 56.72) | 3.52E-03 | 0.94 |
| *RNF125* | 1 | 4 | 10.9 (3.98 - 29.8) | 1.19E-02 | 0.91 | 1.83 (0.68 - 4.92) | *ns* | 0.45 |
| *STARD9* | 1 | 3 | 16 (4.98 - 51.46) | 1.19E-02 | 0.94 | 0.45 (0.14 - 1.4) | *ns* | -1.23 |
| *DLX4* | 1 | 3 | 15.94 (4.95 - 51.3) | 1.25E-02 | 0.94 | 15.2 (4.8 - 48.12) | 1.34E-02 | 0.93 |
| *TDRD12* | 1 | 7 | 6.03 (2.82 - 12.89) | 1.33E-02 | 0.83 | 6.06 (2.84 - 12.92) | 1.11E-02 | 0.83 |
| *OR5AU1* | 1 | 2 | 31.34 (7.28 - 134.95) | 1.36E-02 | 0.97 | 0.31 (0.08 - 1.26) | *ns* | -2.21 |
| *AIMP2* | 1 | 3 | 15.48 (4.82 - 49.73) | 1.53E-02 | 0.94 | 21.39 (6.73 - 68.02) | 7.66E-04 | 0.95 |
| *ABL2* | 2 | 12 | 3.67 (2.11 - 6.4) | 1.60E-02 | 0.73 | 5.53 (3.18 - 9.63) | 5.36E-06 | 0.82 |
| *CCDC183* | 2 | 4 | 5.15 (2.54 - 10.42) | 1.91E-02 | 0.81 | 7.35 (3.64 - 14.86) | 9.76E-05 | 0.86 |
| *AGAP3* | 2 | 6 | 6.53 (2.89 - 14.75) | 2.27E-02 | 0.85 | 7.67 (3.41 - 17.24) | 3.04E-03 | 0.87 |
| *EVL* | 1 | 2 | 27.25 (6.38 - 116.33) | 2.92E-02 | 0.96 | 0.28 (0.07 - 1.13) | *ns* | -2.57 |
| *TRANK1* | 1 | 2 | 27.24 (6.38 - 116.3) | 2.93E-02 | 0.96 | 14.03 (3.44 - 57.27) | *ns* | 0.93 |
| *NTSR2* | 1 | 2 | 26.12 (6.13 - 111.23) | 3.69E-02 | 0.96 | 0.4 (0.1 - 1.6) | *ns* | -1.52 |
| *LIG1* | 2 | 9 | 4.44 (2.28 - 8.63) | 4.05E-02 | 0.77 | 6.1 (3.14 - 11.84) | 3.31E-04 | 0.84 |

* Non-Finnish European subpopulation in gnomAD genome v4.1.

^†^ All populations in gnomAD genome v4.1.

^‡^ Odds ratio with 95% confidence interval.

Abbreviations: CADD, Combined Annotation Dependent Depletion; NFE, Non-Finnish European; OR, Odds Ratio, CI, Confidence Interval; EF, Etiological Fraction; *ns*, non-significant.

Note: Genes are ordered by adjusted *P*-values (Bonferroni correction) of gnomADg (NFE).

#### **Supplementary Table 14: Genes enriched for high-confidence loss-of-function (LoF) and missense variants (CADD ≥ 20) associated with moderate hyperacusis (11 ≤ GÜF ≤ 17, n = 91).**

**Genes enriched with LoF variants**

| Gene | Number of variants | Number of individuals | gnomADg (NFE)* | | | gnomADg (global)^†^ | | |
| --- | --- | --- | --- | --- | --- | --- | --- | --- |
|  |  |  | **OR (95% CI)**^‡^ | **Adjusted-*P*** | **EF** | **OR (95% CI)** | **Adjusted-*P*** | **EF** |
| *PRKRA* | 2 | 5 | 51.34 (20 - 131.79) | 5.29E-14 | 0.98 | 88.45 (34.91 - 224.13) | 6.77E-19 | 0.99 |
| *TTC6* | 1 | 3 | 40.04 (12.04 - 133.18) | 3.52E-07 | 0.98 | 55.79 (17.15 - 181.48) | 4.68E-09 | 0.98 |
| *SAA1* | 1 | 2 | 79.71 (17.1 - 371.53) | 4.91E-06 | 0.99 | 1.18 (0.29 - 4.75) | *ns* | 0.15 |
| *ALDH3A1* | 1 | 2 | 79.69 (17.1 - 371.41) | 4.92E-06 | 0.99 | 38.82 (9.32 - 161.69) | 9.97E-05 | 0.97 |
| *ENDOG* | 1 | 3 | 12.07 (4.41 - 33.04) | 2.49E-04 | 0.92 | 10.5 (3.87 - 28.45) | 7.62E-04 | 0.9 |
| *PIK3C2G* | 2 | 5 | 8.55 (3.5 - 20.87) | 4.95E-04 | 0.88 | 12.37 (5.08 - 30.11) | 5.89E-06 | 0.92 |
| *CALHM2* | 1 | 2 | 27.59 (6.5 - 117.12) | 1.37E-03 | 0.96 | 20.95 (5.11 - 85.94) | 4.78E-03 | 0.95 |
| *CAPN12* | 1 | 2 | 24.73 (5.86 - 104.4) | 2.53E-03 | 0.96 | 40.8 (9.78 - 170.25) | 7.18E-05 | 0.98 |

**Genes enriched with missense variants**

| Gene | Number of variants | Number of individuals | gnomADg (NFE)* | | | gnomADg (global)^†^ | | |
| --- | --- | --- | --- | --- | --- | --- | --- | --- |
|  |  |  | **OR (95% CI)**^‡^ | **Adjusted-*P*** | **EF** | **OR (95% CI)** | **Adjusted-*P*** | **EF** |
| *EIF4G1* | 1 | 12 | 397.79 (173.13 - 913.98) | 1.21E-41 | 1 | 263.64 (135.14 - 514.32) | 1.46E-56 | 1 |
| *BEND3* | 1 | 10 | 3745.41 (476.84 - 29418.84) | 1.71E-11 | 1 | 8305.64 (1057.43 - 65237.34) | 3.13E-14 | 1 |
| *FBXW2* | 1 | 5 | 1773.81 (206.18 - 15260.48) | 3.21E-08 | 1 | 435.8 (144.61 - 1313.31) | 1.18E-23 | 1 |
| *SLC12A8* | 1 | 5 | 21.93 (8.79 - 54.73) | 1.22E-07 | 0.95 | 24.79 (10.06 - 61.1) | 1.02E-08 | 0.96 |
| *EP400* | 1 | 4 | 1418.31 (157.74 - 12752.77) | 3.15E-07 | 1 | 777.48 (192.94 - 3133.06) | 2.66E-17 | 1 |
| *ZNF302* | 1 | 4 | 23.01 (8.29 - 63.9) | 5.90E-06 | 0.96 | 3.37 (1.25 - 9.1) | *ns* | 0.7 |
| *LRRC69* | 1 | 3 | 37.3 (11.26 - 123.56) | 1.07E-05 | 0.97 | 40.64 (12.62 - 130.84) | 1.78E-06 | 0.98 |
| *IFI35* | 1 | 9 | 6.24 (3.17 - 12.25) | 3.66E-04 | 0.84 | 8.01 (4.09 - 15.71) | 4.60E-06 | 0.88 |
| *CYP4V2* | 1 | 3 | 24.02 (7.4 - 78.02) | 4.10E-04 | 0.96 | 5.57 (1.77 - 17.52) | *ns* | 0.82 |
| *KLKB1* | 1 | 3 | 24.02 (7.4 - 78.02) | 4.10E-04 | 0.96 | 5.57 (1.77 - 17.52) | *ns* | 0.82 |
| *BPIFB4* | 1 | 4 | 14.61 (5.32 - 40.12) | 6.62E-04 | 0.93 | 15.14 (5.57 - 41.16) | 3.37E-04 | 0.93 |
| *AGAP3* | 1 | 2 | 50.28 (11.35 - 222.84) | 8.40E-04 | 0.98 | 39.05 (9.37 - 162.81) | 1.63E-03 | 0.97 |
| *CDC25B* | 1 | 2 | 47.82 (10.86 - 210.63) | 1.07E-03 | 0.98 | 14.2 (3.48 - 57.93) | *ns* | 0.93 |
| *ZNF280A* | 1 | 3 | 20.8 (6.44 - 67.22) | 1.33E-03 | 0.95 | 19.82 (6.25 - 62.92) | 1.34E-03 | 0.95 |
| *PON3* | 1 | 2 | 39.82 (9.17 - 172.89) | 2.92E-03 | 0.97 | 2.9 (0.72 - 11.7) | *ns* | 0.65 |
| *DLX4* | 1 | 3 | 18.25 (5.66 - 58.82) | 3.85E-03 | 0.95 | 17.41 (5.49 - 55.17) | 4.07E-03 | 0.94 |
| *ATF6* | 1 | 2 | 35.82 (8.31 - 154.4) | 5.29E-03 | 0.97 | 72.19 (16.85 - 309.26) | 2.74E-05 | 0.99 |
| *PCSK4* | 1 | 8 | 5.73 (2.81 - 11.7) | 5.54E-03 | 0.83 | 9.92 (4.86 - 20.24) | 9.37E-07 | 0.9 |
| *SLC12A6* | 1 | 2 | 34.15 (7.95 - 146.7) | 6.93E-03 | 0.97 | 49.71 (11.82 - 208.98) | 3.27E-04 | 0.98 |
| *NCOA6* | 1 | 4 | 11.14 (4.07 - 30.46) | 8.83E-03 | 0.91 | 14.89 (5.48 - 40.47) | 4.02E-04 | 0.93 |
| *GDF7* | 1 | 2 | 32.58 (7.61 - 139.59) | 9.03E-03 | 0.97 | 0.83 (0.21 - 3.36) | *ns* | -0.2 |
| *PKHD1L1* | 8 | 32 | 2.07 (1.51 - 2.84) | 1.96E-02 | 0.52 | 1.9 (1.39 - 2.6) | *ns* | 0.47 |
| *PPM1G* | 2 | 2 | 10.08 (3.71 - 27.36) | 1.96E-02 | 0.9 | 16.21 (5.99 - 43.86) | 1.38E-04 | 0.94 |
| *CFAP53* | 1 | 4 | 7.77 (3.16 - 19.07) | 2.57E-02 | 0.87 | 0.78 (0.32 - 1.9) | *ns* | -0.28 |
| *PTAFR* | 1 | 5 | 7.76 (3.16 - 19.04) | 2.61E-02 | 0.87 | 10.12 (4.14 - 24.74) | 1.32E-03 | 0.9 |
| *LAT* | 1 | 2 | 25.61 (6.06 - 108.33) | 3.50E-02 | 0.96 | 29.47 (7.13 - 121.79) | 9.93E-03 | 0.97 |
| *KCNK13* | 1 | 2 | 25.61 (6.06 - 108.32) | 3.50E-02 | 0.96 | 40.82 (9.78 - 170.3) | 1.21E-03 | 0.98 |
| *FERMT2* | 1 | 5 | 7.38 (3.01 - 18.12) | 4.25E-02 | 0.86 | 12.86 (5.25 - 31.49) | 7.68E-05 | 0.92 |

* Non-Finnish European subpopulation in gnomAD genome v4.1.

^†^ All populations in gnomAD genome v4.1.

^‡^ Odds ratio with 95% confidence interval.

Abbreviations: CADD, Combined Annotation Dependent Depletion; NFE, Non-Finnish European; OR, Odds Ratio, CI, Confidence Interval; EF, Etiological Fraction; *ns*, non-significant.

Note: Genes are ordered by adjusted *P*-values (Bonferroni correction) of gnomADg (NFE).

#### **Supplementary Table 15: Genes enriched for high-confidence loss-of-function (LoF) and missense variants (CADD ≥ 20) associated with severe hyperacusis (18 ≤ GÜF ≤ 25, n = 54).**

**Genes enriched with LoF variants**

| Gene | Number of variants | Number of individuals | gnomADg (NFE)* | | | gnomADg (global)^†^ | | |
| --- | --- | --- | --- | --- | --- | --- | --- | --- |
|  |  |  | **OR (95% CI)**^‡^ | **Adjusted-*P*** | **EF** | **OR (95% CI)** | **Adjusted-*P*** | **EF** |
| *LILRB3* | 2 | 2 | 119.61 (34.38 - 416.17) | 7.59E-12 | 0.99 | 173.02 (51.56 - 580.57) | 1.00E-14 | 0.99 |
| *TRMT9B* | 1 | 2 | 40.57 (12.41 - 132.6) | 1.23E-07 | 0.98 | 24.63 (7.74 - 78.39) | 8.14E-06 | 0.96 |
| *CD36* | 1 | 2 | 76.1 (17.28 - 335.06) | 1.41E-06 | 0.99 | 1.26 (0.31 - 5.09) | *ns* | 0.2 |
| *RBM23* | 1 | 2 | 64.04 (14.73 - 278.37) | 4.02E-06 | 0.98 | 81.85 (19.39 - 345.46) | 2.82E-07 | 0.99 |
| *PRKRA* | 1 | 2 | 35.78 (8.49 - 150.84) | 1.52E-04 | 0.97 | 61.26 (14.66 - 255.93) | 2.35E-06 | 0.98 |
| *DOCK6* | 1 | 2 | 23.42 (5.63 - 97.38) | 2.01E-03 | 0.96 | 0.41 (0.1 - 1.65) | *ns* | -1.46 |
| *KCTD19* | 1 | 2 | 21.74 (5.24 - 90.24) | 3.10E-03 | 0.95 | 24.34 (5.94 - 99.84) | 1.29E-03 | 0.96 |
| *PXMP2* | 2 | 3 | 11.83 (3.74 - 37.37) | 3.57E-03 | 0.92 | 12.69 (4.04 - 39.88) | 1.89E-03 | 0.92 |
| *TOMM20L* | 1 | 2 | 13.83 (3.36 - 56.9) | 3.79E-02 | 0.93 | 15.53 (3.8 - 63.39) | 1.85E-02 | 0.94 |

**Genes enriched with missense variants**

| Gene | Number of variants | Number of individuals | gnomADg (NFE)* | | | gnomADg (global)^†^ | | |
| --- | --- | --- | --- | --- | --- | --- | --- | --- |
|  |  |  | **OR (95% CI)**^‡^ | **Adjusted-*P*** | **EF** | **OR (95% CI)** | **Adjusted-*P*** | **EF** |
| *AGAP3* | 1 | 4 | 174.05 (56.35 - 537.58) | 6.93E-16 | 0.99 | 135.19 (47.51 - 384.64) | 8.36E-17 | 0.99 |
| *BEND3* | 1 | 10 | 6573.57 (833.49 - 51844.54) | 1.65E-13 | 1 | 14577.24 (1848.32 - 114967.12) | 2.07E-16 | 1 |
| *EIF4G1* | 1 | 3 | 161.01 (44.28 - 585.44) | 2.74E-11 | 0.99 | 106.71 (32.4 - 351.49) | 3.65E-11 | 0.99 |
| *OR2AG1* | 2 | 2 | 52.93 (18.89 - 148.36) | 1.00E-10 | 0.98 | 0.48 (0.18 - 1.3) | *ns* | -1.07 |
| *GLB1L3* | 1 | 3 | 61.48 (18.48 - 204.55) | 4.28E-08 | 0.98 | 34.38 (10.76 - 109.87) | 5.43E-06 | 0.97 |
| *NCKAP5* | 1 | 2 | 135.29 (28.89 - 633.62) | 1.06E-06 | 0.99 | 9.33 (2.29 - 37.98) | *ns* | 0.89 |
| *TRPC7* | 1 | 4 | 22.14 (8.02 - 61.12) | 5.18E-06 | 0.95 | 32.38 (11.8 - 88.91) | 3.38E-08 | 0.97 |
| *LGALS12* | 1 | 2 | 81.2 (18.34 - 359.44) | 1.57E-05 | 0.99 | 1.39 (0.34 - 5.64) | *ns* | 0.28 |
| *DLX4* | 1 | 3 | 31.11 (9.59 - 100.92) | 2.34E-05 | 0.97 | 29.67 (9.3 - 94.67) | 2.32E-05 | 0.97 |
| *THAP5* | 1 | 2 | 67.66 (15.51 - 295.25) | 4.68E-05 | 0.99 | 51.01 (12.27 - 212.01) | 1.44E-04 | 0.98 |
| *SLIT3* | 1 | 2 | 58.01 (13.43 - 250.49) | 1.21E-04 | 0.98 | 0.59 (0.14 - 2.37) | *ns* | -0.71 |
| *CLCN2* | 1 | 3 | 24.9 (7.73 - 80.24) | 1.64E-04 | 0.96 | 21.08 (6.63 - 66.98) | 5.41E-04 | 0.95 |
| *KRT12* | 1 | 4 | 15.29 (5.57 - 42.01) | 2.78E-04 | 0.93 | 21.49 (7.86 - 58.78) | 5.20E-06 | 0.95 |
| *OR2B11* | 1 | 3 | 22.19 (6.9 - 71.32) | 4.46E-04 | 0.95 | 3.04 (0.96 - 9.59) | *ns* | 0.67 |
| *TESK2* | 1 | 3 | 21.93 (6.82 - 70.48) | 4.93E-04 | 0.95 | 20.75 (6.53 - 65.94) | 6.21E-04 | 0.95 |
| *BEST2* | 1 | 7 | 7.53 (3.48 - 16.27) | 6.37E-04 | 0.87 | 11.03 (5.11 - 23.79) | 2.14E-06 | 0.91 |
| *PARP4* | 1 | 2 | 39.28 (9.28 - 166.2) | 1.39E-03 | 0.97 | 50.98 (12.27 - 211.9) | 1.44E-04 | 0.98 |
| *MUC17* | 1 | 3 | 17.06 (5.33 - 54.58) | 3.98E-03 | 0.94 | 11.94 (3.77 - 37.81) | *ns* | 0.92 |
| *BRMS1L* | 1 | 2 | 29.7 (7.09 - 124.36) | 7.87E-03 | 0.97 | 42.24 (10.21 - 174.81) | 5.43E-04 | 0.98 |
| *GLS2* | 1 | 2 | 28.98 (6.93 - 121.27) | 9.13E-03 | 0.97 | 50 (12.04 - 207.71) | 1.66E-04 | 0.98 |
| *PTPRU* | 1 | 2 | 27.06 (6.48 - 112.96) | 1.39E-02 | 0.96 | 0.57 (0.14 - 2.32) | *ns* | -0.75 |
| *C17orf113* | 1 | 6 | 6.7 (2.93 - 15.32) | 1.51E-02 | 0.85 | 10.31 (4.51 - 23.55) | 7.13E-05 | 0.9 |
| *NYAP2* | 3 | 5 | 4.57 (2.35 - 8.88) | 1.73E-02 | 0.78 | 4.35 (2.24 - 8.45) | 3.18E-02 | 0.77 |
| *DNAJC18* | 1 | 2 | 25.88 (6.21 - 107.94) | 1.81E-02 | 0.96 | 27.24 (6.63 - 111.89) | 1.03E-02 | 0.96 |
| *SLC28A1* | 1 | 8 | 5.19 (2.52 - 10.69) | 1.84E-02 | 0.81 | 1.61 (0.78 - 3.31) | *ns* | 0.38 |
| *EVC2* | 1 | 2 | 25.31 (6.07 - 105.48) | 2.07E-02 | 0.96 | 3.36 (0.83 - 13.62) | *ns* | 0.7 |
| *SERINC5* | 1 | 2 | 24.85 (5.97 - 103.48) | 2.31E-02 | 0.96 | 0.53 (0.13 - 2.14) | *ns* | -0.9 |
| *VPS13B* | 1 | 2 | 22.52 (5.42 - 93.57) | 4.13E-02 | 0.96 | 28.13 (6.85 - 115.6) | 8.39E-03 | 0.96 |
| *SDF2* | 1 | 2 | 22.13 (5.33 - 91.91) | 4.57E-02 | 0.95 | 24.12 (5.88 - 98.91) | 2.23E-02 | 0.96 |

* Non-Finnish European subpopulation in gnomAD genome v4.1.

^†^ All populations in gnomAD genome v4.1.

^‡^ Odds ratio with 95% confidence interval.

Abbreviations: CADD, Combined Annotation Dependent Depletion; NFE, Non-Finnish European; OR, Odds Ratio, CI, Confidence Interval; EF, Etiological Fraction; *ns*, non-significant.

Note: Genes are ordered by adjusted *P*-values (Bonferroni correction) of gnomADg (NFE).

#### **Supplementary Table 16: Genes enriched for high-confidence loss-of-function (LoF) and missense variants (CADD ≥ 20) associated with very severe hyperacusis (26 ≤ GÜF ≤ 45, n = 29).**

**Genes enriched with LoF variants**

| Gene | Number of variants | Number of individuals | gnomADg (NFE)* | | | gnomADg (global)^†^ | | |
| --- | --- | --- | --- | --- | --- | --- | --- | --- |
|  |  |  | **OR (95% CI)**^‡^ | **Adjusted-*P*** | **EF** | **OR (95% CI)** | **Adjusted-*P*** | **EF** |
| *IGFN1* | 2 | 3 | 69.93 (21.48 - 227.62) | 1.03E-10 | 0.99 | 79.2 (24.73 - 253.6) | 1.06E-11 | 0.99 |
| *MYO15B* | 2 | 4 | 31.76 (11.56 - 87.24) | 1.17E-09 | 0.97 | 34.65 (12.7 - 94.55) | 2.63E-10 | 0.97 |
| *SLC22A25* | 1 | 2 | 100.24 (23.08 - 435.33) | 4.58E-08 | 0.99 | 4.33 (1.06 - 17.76) | *ns* | 0.77 |
| *GRAMD1A* | 1 | 2 | 16.55 (4 - 68.5) | 6.34E-03 | 0.94 | 28.08 (6.8 - 115.94) | 2.37E-04 | 0.96 |
| *USP54* | 1 | 2 | 14.83 (3.59 - 61.31) | 1.16E-02 | 0.93 | 14.95 (3.63 - 61.52) | 1.05E-02 | 0.93 |
| *CCDC102B* | 1 | 2 | 11.94 (2.89 - 49.27) | 3.58E-02 | 0.92 | 21.95 (5.32 - 90.46) | 1.13E-03 | 0.95 |
| *PPIC* | 1 | 3 | 7.28 (2.27 - 23.36) | 4.96E-02 | 0.86 | 11.05 (3.45 - 35.4) | 3.11E-03 | 0.91 |

**Genes enriched with missense variants**

| Gene | Number of variants | Number of individuals | gnomADg (NFE)* | | | gnomADg (global)^†^ | | |
| --- | --- | --- | --- | --- | --- | --- | --- | --- |
|  |  |  | **OR (95% CI)**^‡^ | **Adjusted-*P*** | **EF** | **OR (95% CI)** | **Adjusted-*P*** | **EF** |
| *BEND3* | 1 | 4 | 4771.93 (524.78 - 43392.33) | 6.86E-11 | 1 | 10582 (1163.73 - 96224.08) | 2.40E-13 | 1 |
| *DNHD1* | 1 | 2 | 209.67 (45.43 - 967.58) | 9.23E-09 | 1 | 5.06 (1.23 - 20.76) | *ns* | 0.8 |
| *TEAD3* | 1 | 2 | 88.66 (20.55 - 382.51) | 2.30E-06 | 0.99 | 138.33 (32.55 - 587.86) | 3.04E-08 | 0.99 |
| *PBK* | 1 | 2 | 67.74 (15.89 - 288.78) | 1.52E-05 | 0.99 | 2.07 (0.51 - 8.51) | *ns* | 0.52 |
| *USP34* | 1 | 2 | 60.65 (14.28 - 257.48) | 3.31E-05 | 0.98 | 71.97 (17.23 - 300.58) | 5.72E-06 | 0.99 |
| *ABCC12* | 1 | 2 | 49.04 (11.63 - 206.82) | 1.45E-04 | 0.98 | 1.36 (0.33 - 5.59) | *ns* | 0.27 |
| *SCARA5* | 1 | 2 | 49.03 (11.63 - 206.79) | 1.45E-04 | 0.98 | 1.41 (0.34 - 5.79) | *ns* | 0.29 |
| *MTCH2* | 1 | 2 | 47.19 (10.48 - 212.5) | 6.48E-04 | 0.98 | 38.04 (8.98 - 161.1) | 9.77E-04 | 0.97 |
| *ASB18* | 1 | 2 | 37.78 (9.02 - 158.28) | 8.49E-04 | 0.97 | 1.97 (0.48 - 8.1) | *ns* | 0.49 |
| *GPRC5C* | 1 | 2 | 36.01 (8.6 - 150.7) | 1.17E-03 | 0.97 | 37.07 (8.96 - 153.4) | 7.78E-04 | 0.97 |
| *ANKRD52* | 1 | 2 | 30.31 (7.27 - 126.44) | 3.59E-03 | 0.97 | 44.85 (10.82 - 186.01) | 2.01E-04 | 0.98 |
| *PYGM* | 1 | 2 | 29.91 (7.17 - 124.74) | 3.90E-03 | 0.97 | 44.45 (10.72 - 184.3) | 2.15E-04 | 0.98 |
| *SCN5A* | 1 | 2 | 28.43 (6.82 - 118.47) | 5.39E-03 | 0.96 | 1.15 (0.28 - 4.7) | *ns* | 0.13 |
| *ADGRL2* | 4 | 12 | 3.87 (2.16 - 6.93) | 6.51E-03 | 0.74 | 4.84 (2.71 - 8.67) | 1.32E-04 | 0.79 |
| *RPTN* | 1 | 2 | 26.47 (6.36 - 110.19) | 8.44E-03 | 0.96 | 47.36 (11.41 - 196.51) | 1.35E-04 | 0.98 |
| *GJB2* | 1 | 2 | 24.5 (5.89 - 101.85) | 1.36E-02 | 0.96 | 10.28 (2.5 - 42.25) | *ns* | 0.9 |
| *PTPRB* | 1 | 2 | 24.24 (5.83 - 100.77) | 1.45E-02 | 0.96 | 30.07 (7.28 - 124.19) | 3.23E-03 | 0.97 |
| *COPG2* | 1 | 2 | 23.26 (5.6 - 96.63) | 1.87E-02 | 0.96 | 37.04 (8.95 - 153.3) | 7.82E-04 | 0.97 |
| *MLXIPL* | 1 | 2 | 23.03 (5.54 - 95.65) | 1.99E-02 | 0.96 | 32.99 (7.98 - 136.36) | 1.73E-03 | 0.97 |
| *SHMT1* | 1 | 2 | 22.57 (5.44 - 93.75) | 2.24E-02 | 0.96 | 40.59 (9.8 - 168.12) | 4.11E-04 | 0.98 |
| *A3GALT2* | 3 | 4 | 5.86 (2.59 - 13.25) | 2.77E-02 | 0.83 | 10.17 (4.5 - 23) | 3.17E-05 | 0.9 |
| *OR52N2* | 2 | 3 | 12 (3.79 - 38) | 2.98E-02 | 0.92 | 1.23 (0.39 - 3.87) | *ns* | 0.19 |

* Non-Finnish European subpopulation in gnomAD genome v4.1.

^†^ All populations in gnomAD genome v4.1.

^‡^ Odds ratio with 95% confidence interval.

Abbreviations: CADD, Combined Annotation Dependent Depletion; NFE, Non-Finnish European; OR, Odds Ratio, CI, Confidence Interval; EF, Etiological Fraction; *ns*, non-significant.

Note: Genes are ordered by adjusted *P*-values (Bonferroni correction) of gnomADg (NFE).

#### **Supplementary Table 17: Genes enriched for high-confidence loss-of-function (LoF) and missense variants (CADD ≥ 20) associated with mild depressive symptoms (5 ≤ PHQ-9 ≤ 9, n = 104).**

**Genes enriched with LoF variants**

| Gene | Number of variants | Number of individuals | gnomADg (NFE)* | | | gnomADg (global)^†^ | | |
| --- | --- | --- | --- | --- | --- | --- | --- | --- |
|  |  |  | **OR (95% CI)**^‡^ | **Adjusted-*P*** | **EF** | **OR (95% CI)** | **Adjusted-*P*** | **EF** |
| *PRKRA* | 3 | 6 | 13.51 (5.94 - 30.71) | 1.12E-07 | 0.93 | 13.92 (6.18 - 31.35) | 4.53E-08 | 0.93 |
| *IGFN1* | 2 | 4 | 25.57 (9.19 - 71.18) | 1.19E-07 | 0.96 | 28.96 (10.6 - 79.11) | 1.14E-08 | 0.97 |
| *HK3* | 1 | 2 | 78.36 (16.54 - 371.23) | 8.51E-06 | 0.99 | 99.35 (22.44 - 439.9) | 3.02E-07 | 0.99 |
| *ALDH3A1* | 1 | 2 | 69.63 (14.95 - 324.25) | 1.40E-05 | 0.99 | 33.92 (8.15 - 141.15) | 2.79E-04 | 0.97 |
| *H4C2* | 2 | 3 | 25.35 (7.78 - 82.53) | 1.74E-05 | 0.96 | 33.57 (10.5 - 107.37) | 6.92E-07 | 0.97 |
| *TRMT9B* | 1 | 2 | 20.78 (6.41 - 67.41) | 9.46E-05 | 0.95 | 12.61 (3.99 - 39.84) | 3.42E-03 | 0.92 |
| *ALMS1* | 1 | 2 | 38.95 (8.9 - 170.47) | 2.53E-04 | 0.97 | 44.57 (10.6 - 187.46) | 4.83E-05 | 0.98 |
| *RBM23* | 1 | 2 | 32.95 (7.63 - 142.38) | 6.23E-04 | 0.97 | 42.12 (10.04 - 176.68) | 6.94E-05 | 0.98 |
| *AIRE* | 2 | 3 | 9.66 (3.05 - 30.6) | 2.52E-02 | 0.9 | 12.68 (4.03 - 39.9) | 3.05E-03 | 0.92 |

**Genes enriched with missense variants**

| Gene | Number of variants | Number of individuals | gnomADg (NFE)* | | | gnomADg (global)^†^ | | |
| --- | --- | --- | --- | --- | --- | --- | --- | --- |
|  |  |  | **OR (95% CI)**^‡^ | **Adjusted-*P*** | **EF** | **OR (95% CI)** | **Adjusted-*P*** | **EF** |
| *EIF4G1* | 1 | 8 | 225.41 (89.72 - 566.34) | 3.49E-27 | 1 | 149.4 (68.71 - 324.83) | 4.94E-33 | 0.99 |
| *AGAP3* | 1 | 5 | 111.46 (39.78 - 312.33) | 1.11E-15 | 0.99 | 86.57 (33.82 - 221.59) | 4.92E-17 | 0.99 |
| *BEND3* | 1 | 15 | 5006.81 (658.1 - 38092) | 6.87E-13 | 1 | 11102.88 (1459.37 - 84470.35) | 8.35E-16 | 1 |
| *SLC12A8* | 1 | 6 | 23.06 (9.96 - 53.42) | 8.80E-10 | 0.96 | 26.07 (11.41 - 59.57) | 3.76E-11 | 0.96 |
| *EP400* | 1 | 5 | 1554.56 (180.81 - 13365.54) | 7.79E-08 | 1 | 852.17 (227.19 - 3196.39) | 5.27E-20 | 1 |
| *MTCH2* | 1 | 5 | 32.55 (11.61 - 91.21) | 1.26E-07 | 0.97 | 26.24 (10.26 - 67.07) | 3.23E-08 | 0.96 |
| *FBXW2* | 1 | 4 | 1231.24 (137.02 - 11063.72) | 7.67E-07 | 1 | 302.49 (92.41 - 990.15) | 1.33E-17 | 1 |
| *GDF7* | 1 | 3 | 42.91 (12.74 - 144.5) | 4.65E-06 | 0.98 | 1.1 (0.35 - 3.44) | *ns* | 0.09 |
| *WWC2* | 1 | 4 | 22.59 (8.12 - 62.89) | 8.60E-06 | 0.96 | 37.45 (13.57 - 103.34) | 9.57E-09 | 0.97 |
| *ADM* | 1 | 6 | 10.87 (4.76 - 24.8) | 5.29E-05 | 0.91 | 1.34 (0.59 - 3.02) | *ns* | 0.25 |
| *PSKH1* | 1 | 3 | 30.47 (9.24 - 100.45) | 7.17E-05 | 0.97 | 21.82 (6.86 - 69.42) | 6.42E-04 | 0.95 |
| *RNF220* | 1 | 3 | 30.45 (9.24 - 100.41) | 7.19E-05 | 0.97 | 20.53 (6.46 - 65.26) | 1.09E-03 | 0.95 |
| *NCKAP5* | 1 | 2 | 69.62 (14.95 - 324.18) | 2.32E-04 | 0.99 | 4.8 (1.19 - 19.42) | *ns* | 0.79 |
| *EMP1* | 1 | 5 | 10.71 (4.35 - 26.39) | 9.22E-04 | 0.91 | 15.05 (6.14 - 36.89) | 1.11E-05 | 0.93 |
| *SNIP1* | 1 | 2 | 48.2 (10.81 - 214.95) | 1.36E-03 | 0.98 | 73.16 (16.93 - 316.09) | 3.24E-05 | 0.99 |
| *CFAP53* | 1 | 5 | 8.17 (3.59 - 18.58) | 1.99E-03 | 0.88 | 0.82 (0.36 - 1.85) | *ns* | -0.22 |
| *PRX* | 1 | 2 | 44.76 (10.11 - 198.18) | 1.99E-03 | 0.98 | 0.47 (0.12 - 1.89) | *ns* | -1.13 |
| *EIF3F* | 1 | 2 | 44.75 (10.11 - 198.15) | 1.99E-03 | 0.98 | 69.49 (16.14 - 299.2) | 4.49E-05 | 0.99 |
| *CDC25B* | 1 | 2 | 41.78 (9.49 - 183.87) | 2.86E-03 | 0.98 | 12.41 (3.05 - 50.57) | *ns* | 0.92 |
| *FMO2* | 2 | 8 | 5.79 (2.86 - 11.73) | 3.94E-03 | 0.83 | 4.09 (2.03 - 8.24) | *ns* | 0.76 |
| *C2CD3* | 6 | 32 | 2.32 (1.64 - 3.29) | 6.99E-03 | 0.57 | 0.97 (0.68 - 1.37) | *ns* | -0.03 |
| *DLX4* | 1 | 3 | 15.94 (4.95 - 51.3) | 1.25E-02 | 0.94 | 15.2 (4.8 - 48.12) | 1.33E-02 | 0.93 |
| *ANP32D* | 1 | 4 | 10.8 (3.95 - 29.55) | 1.28E-02 | 0.91 | 17.76 (6.52 - 48.37) | 6.54E-05 | 0.94 |
| *TDRD12* | 1 | 7 | 6.03 (2.82 - 12.89) | 1.33E-02 | 0.83 | 6.06 (2.84 - 12.92) | 1.11E-02 | 0.83 |
| *BPIFB4* | 2 | 6 | 6.7 (2.97 - 15.13) | 1.69E-02 | 0.85 | 8.61 (3.82 - 19.36) | 7.10E-04 | 0.88 |
| *SELENOS* | 1 | 4 | 9.95 (3.64 - 27.18) | 2.67E-02 | 0.9 | 0.67 (0.25 - 1.82) | *ns* | -0.48 |
| *ANO2* | 1 | 2 | 27.25 (6.38 - 116.32) | 2.91E-02 | 0.96 | 2.24 (0.56 - 9.04) | *ns* | 0.55 |
| *DAZL* | 1 | 2 | 26.11 (6.13 - 111.2) | 3.68E-02 | 0.96 | 1.36 (0.34 - 5.5) | *ns* | 0.27 |

* Non-Finnish European subpopulation in gnomAD genome v4.1.

^†^ All populations in gnomAD genome v4.1.

^‡^ Odds ratio with 95% confidence interval.

Abbreviations: CADD, Combined Annotation Dependent Depletion; NFE, Non-Finnish European; OR, Odds Ratio, CI, Confidence Interval; EF, Etiological Fraction; *ns*, non-significant.

Note: Genes are ordered by adjusted *P*-values (Bonferroni correction) of gnomADg (NFE).

#### **Supplementary Table 18: Genes enriched for high-confidence loss-of-function (LoF) and missense variants (CADD ≥ 20) associated with moderate depressive symptoms (10 ≤ PHQ-9 ≤ 14, n = 43).**

**Genes enriched with LoF variants**

| Gene | Number of variants | Number of individuals | gnomADg (NFE)* | | | gnomADg (global)^†^ | | |
| --- | --- | --- | --- | --- | --- | --- | --- | --- |
|  |  |  | **OR (95% CI)**^‡^ | **Adjusted-*P*** | **EF** | **OR (95% CI)** | **Adjusted-*P*** | **EF** |
| *C11orf16* | 1 | 2 | 73.2 (16.89 - 317.12) | 9.89E-07 | 0.99 | 0.7 (0.17 - 2.83) | *ns* | -0.44 |
| *FAM166B* | 1 | 2 | 66.81 (15.5 - 287.86) | 1.79E-06 | 0.99 | 54.09 (13.02 - 224.67) | 4.11E-06 | 0.98 |
| *HOXD12* | 1 | 2 | 61.46 (14.33 - 263.63) | 3.08E-06 | 0.98 | 131.17 (30.64 - 561.47) | 5.12E-09 | 0.99 |
| *CALHM2* | 1 | 2 | 59.13 (13.81 - 253.09) | 3.97E-06 | 0.98 | 44.89 (10.85 - 185.76) | 1.58E-05 | 0.98 |
| *C4orf48* | 1 | 3 | 21.71 (6.76 - 69.76) | 2.47E-05 | 0.95 | 10.73 (3.38 - 34.07) | 5.90E-03 | 0.91 |
| *ENDOG* | 1 | 2 | 19.41 (6.05 - 62.27) | 6.39E-05 | 0.95 | 16.88 (5.31 - 53.71) | 1.77E-04 | 0.94 |
| *PXMP2* | 2 | 3 | 14.91 (4.71 - 47.2) | 4.51E-04 | 0.93 | 16 (5.08 - 50.36) | 2.25E-04 | 0.94 |
| *MPO* | 1 | 4 | 6.9 (2.52 - 18.9) | 1.79E-02 | 0.86 | 10.52 (3.84 - 28.77) | 4.80E-04 | 0.9 |

**Genes enriched with missense variants**

| Gene | Number of variants | Number of individuals | gnomADg (NFE)* | | | gnomADg (global)^†^ | | |
| --- | --- | --- | --- | --- | --- | --- | --- | --- |
|  |  |  | **OR (95% CI)**^‡^ | **Adjusted-*P*** | **EF** | **OR (95% CI)** | **Adjusted-*P*** | **EF** |
| *EIF4G1* | 1 | 3 | 203.69 (55.81 - 743.4) | 1.61E-12 | 1 | 135 (40.82 - 446.46) | 1.77E-12 | 0.99 |
| *AGAP3* | 1 | 3 | 163.56 (46.15 - 579.77) | 5.61E-12 | 0.99 | 127.04 (38.54 - 418.79) | 3.32E-12 | 0.99 |
| *BEND3* | 1 | 6 | 4831.58 (575.11 - 40590.72) | 1.09E-11 | 1 | 10714.27 (1275.35 - 90011.4) | 2.48E-14 | 1 |
| *CYP4V2* | 1 | 3 | 51.81 (15.79 - 170.04) | 1.44E-07 | 0.98 | 12.02 (3.78 - 38.19) | 4.78E-02 | 0.92 |
| *KLKB1* | 1 | 3 | 51.81 (15.79 - 170.04) | 1.44E-07 | 0.98 | 12.02 (3.78 - 38.19) | 4.78E-02 | 0.92 |
| *WWC3* | 2 | 6 | 7.29 (3.83 - 13.85) | 2.61E-06 | 0.86 | 10.71 (5.64 - 20.33) | 8.20E-10 | 0.91 |
| *B4GALT7* | 1 | 3 | 38.22 (11.76 - 124.26) | 2.68E-06 | 0.97 | 59.5 (18.45 - 191.88) | 1.53E-08 | 0.98 |
| *INA* | 1 | 2 | 56.93 (13.32 - 243.21) | 9.46E-05 | 0.98 | 43.73 (10.57 - 180.87) | 3.54E-04 | 0.98 |
| *LZTS3* | 1 | 2 | 49.57 (11.68 - 210.45) | 2.35E-04 | 0.98 | 100.34 (23.72 - 424.36) | 7.27E-07 | 0.99 |
| *PARP4* | 1 | 2 | 49.56 (11.67 - 210.42) | 2.35E-04 | 0.98 | 64.33 (15.43 - 268.28) | 2.11E-05 | 0.98 |
| *KIAA0513* | 1 | 4 | 14.2 (5.16 - 39.07) | 5.40E-04 | 0.93 | 17.91 (6.53 - 49.1) | 3.97E-05 | 0.94 |
| *CCDC40* | 2 | 5 | 9.93 (4.06 - 24.31) | 9.63E-04 | 0.9 | 6.04 (2.48 - 14.71) | *ns* | 0.83 |
| *CEP250* | 1 | 2 | 39.67 (9.42 - 167.08) | 1.02E-03 | 0.97 | 50.84 (12.25 - 210.95) | 1.21E-04 | 0.98 |
| *CEP120* | 2 | 12 | 4.26 (2.41 - 7.5) | 1.06E-03 | 0.77 | 3.32 (1.89 - 5.85) | *ns* | 0.7 |
| *DCAF5* | 1 | 2 | 26.04 (6.26 - 108.29) | 1.43E-02 | 0.96 | 54.12 (13.03 - 224.79) | 7.60E-05 | 0.98 |
| *DLX4* | 1 | 2 | 25.93 (6.23 - 107.93) | 1.49E-02 | 0.96 | 24.73 (6.02 - 101.55) | 1.65E-02 | 0.96 |
| *PGM2L1* | 1 | 11 | 3.98 (2.16 - 7.34) | 1.82E-02 | 0.75 | 4.35 (2.36 - 8) | 4.63E-03 | 0.77 |
| *SRRM4* | 2 | 7 | 4.89 (2.4 - 9.97) | 2.39E-02 | 0.8 | 6.41 (3.15 - 13.04) | 5.89E-04 | 0.84 |
| *PKD1L3* | 1 | 3 | 13.3 (4.16 - 42.53) | 2.45E-02 | 0.92 | 24.52 (7.68 - 78.26) | 1.26E-04 | 0.96 |
| *C2CD3* | 2 | 10 | 3.7 (2.06 - 6.67) | 2.47E-02 | 0.73 | 2.07 (1.15 - 3.73) | *ns* | 0.52 |
| *ANK3* | 1 | 2 | 22.25 (5.37 - 92.26) | 3.68E-02 | 0.96 | 32.44 (7.88 - 133.59) | 2.80E-03 | 0.97 |

* Non-Finnish European subpopulation in gnomAD genome v4.1.

^†^ All populations in gnomAD genome v4.1.

^‡^ Odds ratio with 95% confidence interval.

Abbreviations: CADD, Combined Annotation Dependent Depletion; NFE, Non-Finnish European; OR, Odds Ratio, CI, Confidence Interval; EF, Etiological Fraction; *ns*, non-significant.

Note: Genes are ordered by adjusted *P*-values (Bonferroni correction) of gnomADg (NFE).

#### **Supplementary Table 19: Genes enriched for high-confidence loss-of-function (LoF) and missense variants (CADD ≥ 20) associated with severe depressive symptoms (15 ≤ PHQ-9 ≤ 27, n = 28).**

**Genes enriched with LoF variants**

| Gene | Number of variants | Number of individuals | gnomADg (NFE)* | | | gnomADg (global)^†^ | | |
| --- | --- | --- | --- | --- | --- | --- | --- | --- |
|  |  |  | **OR (95% CI)**^‡^ | **Adjusted-*P*** | **EF** | **OR (95% CI)** | **Adjusted-*P*** | **EF** |
| *NCOA3* | 2 | 2 | 17.53 (6.42 - 47.9) | 1.45E-06 | 0.94 | 4.93 (1.82 - 13.4) | *ns* | 0.8 |
| *TMEM209* | 1 | 3 | 12.41 (3.86 - 39.95) | 1.49E-03 | 0.92 | 14.65 (4.57 - 47.04) | 3.98E-04 | 0.93 |
| *GRAMD1A* | 1 | 2 | 17.17 (4.14 - 71.1) | 5.47E-03 | 0.94 | 29.12 (7.05 - 120.34) | 1.99E-04 | 0.97 |
| *CDHR4* | 1 | 2 | 17.13 (4.14 - 70.95) | 5.53E-03 | 0.94 | 11.62 (2.82 - 47.8) | 4.20E-02 | 0.91 |

**Genes enriched with missense variants**

| Gene | Number of variants | Number of individuals | gnomADg (NFE)* | | | gnomADg (global)^†^ | | |
| --- | --- | --- | --- | --- | --- | --- | --- | --- |
|  |  |  | **OR (95% CI)**^‡^ | **Adjusted-*P*** | **EF** | **OR (95% CI)** | **Adjusted-*P*** | **EF** |
| *BEND3* | 1 | 3 | 3646.47 (373.28 - 35621.28) | 2.08E-09 | 1 | 8086.25 (827.78 - 78991.52) | 1.20E-11 | 1 |
| *OR2B11* | 1 | 3 | 43.96 (13.47 - 143.47) | 4.32E-07 | 0.98 | 6.02 (1.88 - 19.29) | *ns* | 0.83 |
| *KCNQ1* | 1 | 2 | 55.58 (13.13 - 235.22) | 5.69E-05 | 0.98 | 67.15 (16.09 - 280.16) | 9.27E-06 | 0.99 |
| *FRMPD2* | 1 | 5 | 11.85 (4.71 - 29.8) | 1.78E-04 | 0.92 | 17.64 (7.02 - 44.32) | 1.20E-06 | 0.94 |
| *KRT12* | 1 | 3 | 22.51 (6.96 - 72.76) | 2.34E-04 | 0.96 | 31.63 (9.82 - 101.88) | 8.43E-06 | 0.97 |
| *MTCH2* | 1 | 2 | 48.94 (10.86 - 220.55) | 4.84E-04 | 0.98 | 39.45 (9.31 - 167.22) | 7.24E-04 | 0.97 |
| *CEP55* | 1 | 2 | 38.54 (9.19 - 161.56) | 7.00E-04 | 0.97 | 1.21 (0.3 - 4.97) | *ns* | 0.17 |
| *ANKRD18B* | 1 | 2 | 38.53 (9.19 - 161.53) | 7.01E-04 | 0.97 | 1.18 (0.29 - 4.83) | *ns* | 0.15 |
| *RGS22* | 1 | 2 | 35.66 (8.52 - 149.23) | 1.18E-03 | 0.97 | 46.93 (11.31 - 194.81) | 1.37E-04 | 0.98 |
| *PRSS55* | 1 | 2 | 35.14 (8.4 - 147.04) | 1.29E-03 | 0.97 | 34.22 (8.27 - 141.59) | 1.28E-03 | 0.97 |
| *SCN5A* | 1 | 2 | 29.48 (7.07 - 122.97) | 4.05E-03 | 0.97 | 1.19 (0.29 - 4.88) | *ns* | 0.16 |
| *NR2C2* | 1 | 2 | 23.64 (5.69 - 98.26) | 1.61E-02 | 0.96 | 38.7 (9.34 - 160.3) | 5.47E-04 | 0.97 |
| *HAL* | 1 | 3 | 12.81 (3.98 - 41.23) | 2.26E-02 | 0.92 | 21.15 (6.58 - 67.99) | 3.56E-04 | 0.95 |
| *SLC26A6* | 1 | 2 | 20.94 (5.04 - 86.91) | 3.33E-02 | 0.95 | 15.21 (3.69 - 62.64) | *ns* | 0.93 |
| *RTL9* | 1 | 6 | 4.58 (2.24 - 9.35) | 3.59E-02 | 0.78 | 7.33 (3.59 - 14.96) | 5.49E-05 | 0.86 |
| *QRFPR* | 2 | 4 | 4.63 (2.25 - 9.51) | 3.64E-02 | 0.78 | 6.43 (3.13 - 13.21) | 4.82E-04 | 0.84 |

* Non-Finnish European subpopulation in gnomAD genome v4.1.

^†^ All populations in gnomAD genome v4.1.

^‡^ Odds ratio with 95% confidence interval.

Abbreviations: CADD, Combined Annotation Dependent Depletion; NFE, Non-Finnish European; OR, Odds Ratio, CI, Confidence Interval; EF, Etiological Fraction; *ns*, non-significant.

Note: Genes are ordered by adjusted *P*-values (Bonferroni correction) of gnomADg (NFE).

#### **Supplementary Table 20: Splicing effect prediction by *SpliceAI*.**

| **Variant position** | **Gene and associated transcripts** | **Type** | **Score** | **Position** |
| --- | --- | --- | --- | --- |
| chr2:178441705:C>  CAGTTTCCATAAATGACTCTAGCC  TGCAAATTGTAGTATATTCTCTCTTA | *PRKRA* (ENSG00000180228.14), ENST00000325748.9 (MANE transcript) | Acceptor loss | 0.73 | -1 bp |
|  |  | Donor loss | 0.80 | -95 bp |
|  |  | Acceptor gain | 0.06 | +45 bp position within the inserted sequence |
|  |  | Donor gain | 0 |  |
| chr2:178441705:C>  CAGTTTCCATAAATGACTCTAGCC  TGCAAATTGTAGTATATTCTCTCT | *PRKRA* (ENSG00000180228.14), ENST00000325748.9 (MANE transcript) | Acceptor loss | 0.73 | -1 bp |
|  |  | Donor loss | 0.62 | -95 bp |
|  |  | Acceptor gain | 0.28 | +45 bp position within the inserted sequence |
|  |  | Donor gain | 0 |  |

#### **Supplementary Table 21: Splicing effect prediction by *Human Splicing Human Pro*.**

| **Variant position** | **Signal of new encryptic splice site** | **Gene and associated transcript** |
| --- | --- | --- |
| chr2:178441705:C>CAGTTTCCATAAATGACTCTAGCCTGCAAATTGTAGTATATTCTCTCTTA | No signal | *PRKRA*, all transcripts |
| chr2:178441705:C>CAGTTTCCATAAATGACTCTAGCCTGCAAATTGTAGTATATTCTCTCT | No signal | *PRKRA*, all transcripts |

#### **Supplementary Table 22: Binding affinity of protein-kinase interaction between PKR and wild-type (WT)/mutant PACT (~3 kB deletion at ILE-105).**

| **Feature** | **WT PACT-PKR** | **Mutant PACT-PKR** | **Interpretation** |
| --- | --- | --- | --- |
| ΔG (kcal/mol) | -16.7 | -20.2 | Mutant binds more strongly |
| Kd (M) | 5.7E-13 | 1.5E-15 | ~380x tighter binding |
| Charged-charged ICs | 16 | 8 | Fewer salt bridges in mutant |
| Polar-polar ICs | 12 | 22 | More hydrogen bonding |
| Polar-apolar ICs | 41 | 68 | More mixed (adaptive) interactions |
| Apolar-apolar ICs | 45 | 51 | More hydrophobic packing |
| NIS charged/apolar | Similar | Similar | Minimal surface change |

Abbreviations: IC, Intermolecular Contacts; NIS, Non-Interacting Surface.

### **Supplementary Figure**

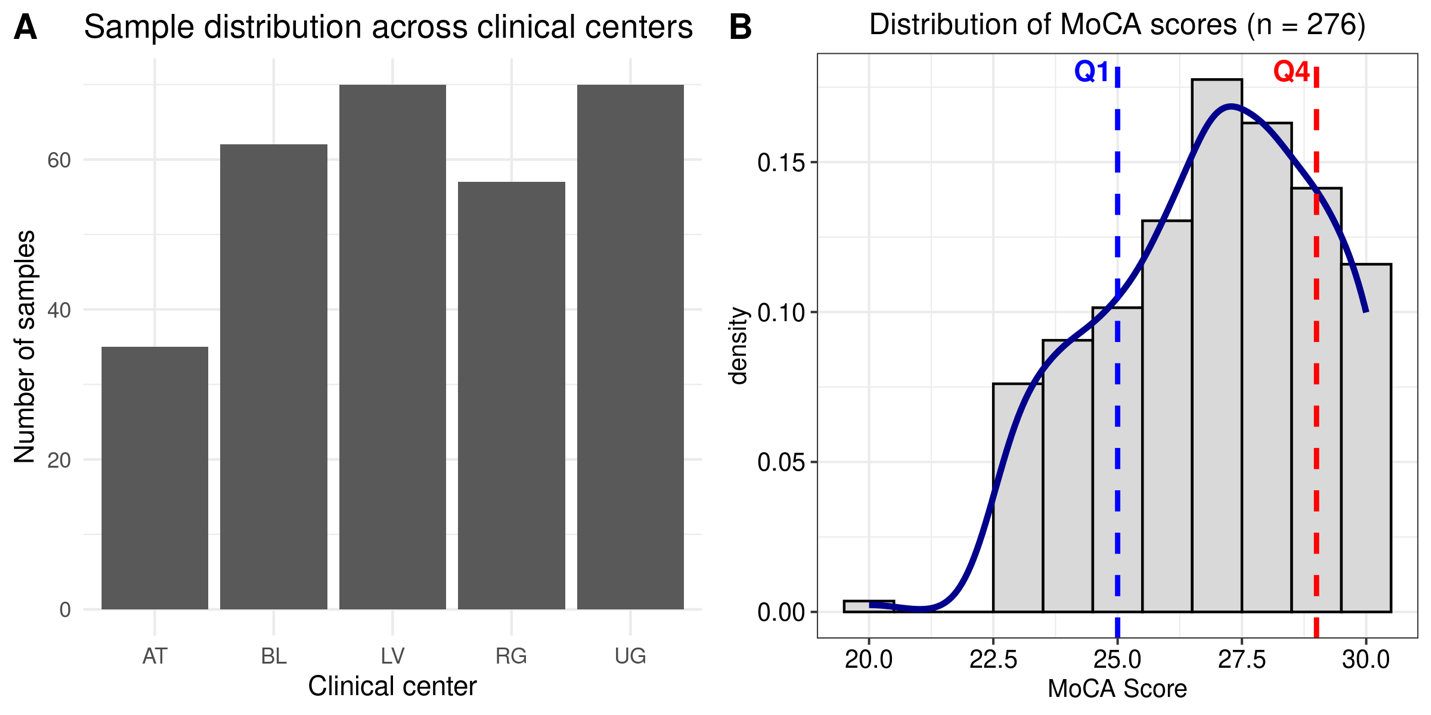

#### **Supplementary Fig. 1 Distribution of sample collection sites and MoCA scores in the UNITI cohort.**

(A) Bar plot shows sample distribution across five clinical centers. AT, Athens; BL, Berlin; LV, Leuven; RG, Regensburg; UG, Granada.

(B) Histogram shows the distribution of MoCA scores (x-axis) as density (y-axis, range 0-1). The first quartile (Q1, 25%) and fourth quartile (Q4, 75%) are marked with dotted blue and red lines, respectively. 18/294 samples have missing MoCA scores.

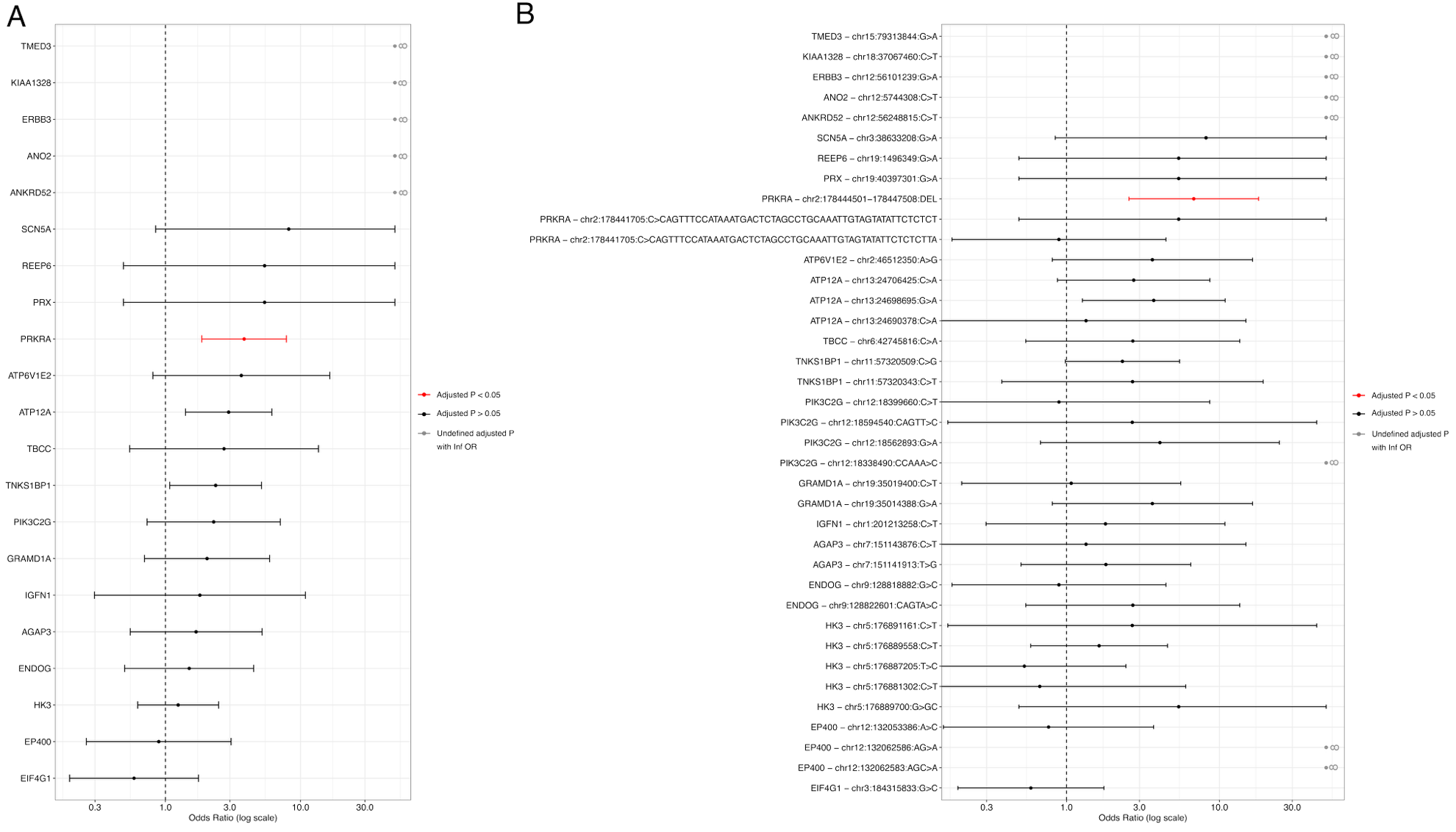

#### **Supplementary Fig. 2 Gene burden (A) and single-variant association analysis (B) between MCI and non-MCI group in 21 synaptic genes previously identified as enriched for MCI vs external controls.**

Log-scaled odd ratios of gene-level and single-variant associations for missense and LoF variants in 21 synaptic genes previously identified as enriched for MCI vs. external controls, shown here for comparison between MCI vs. non-MCI individual (internal cohort). Significant enriched genes and variants are highlighted in red. *P*-values were corrected using BH method.

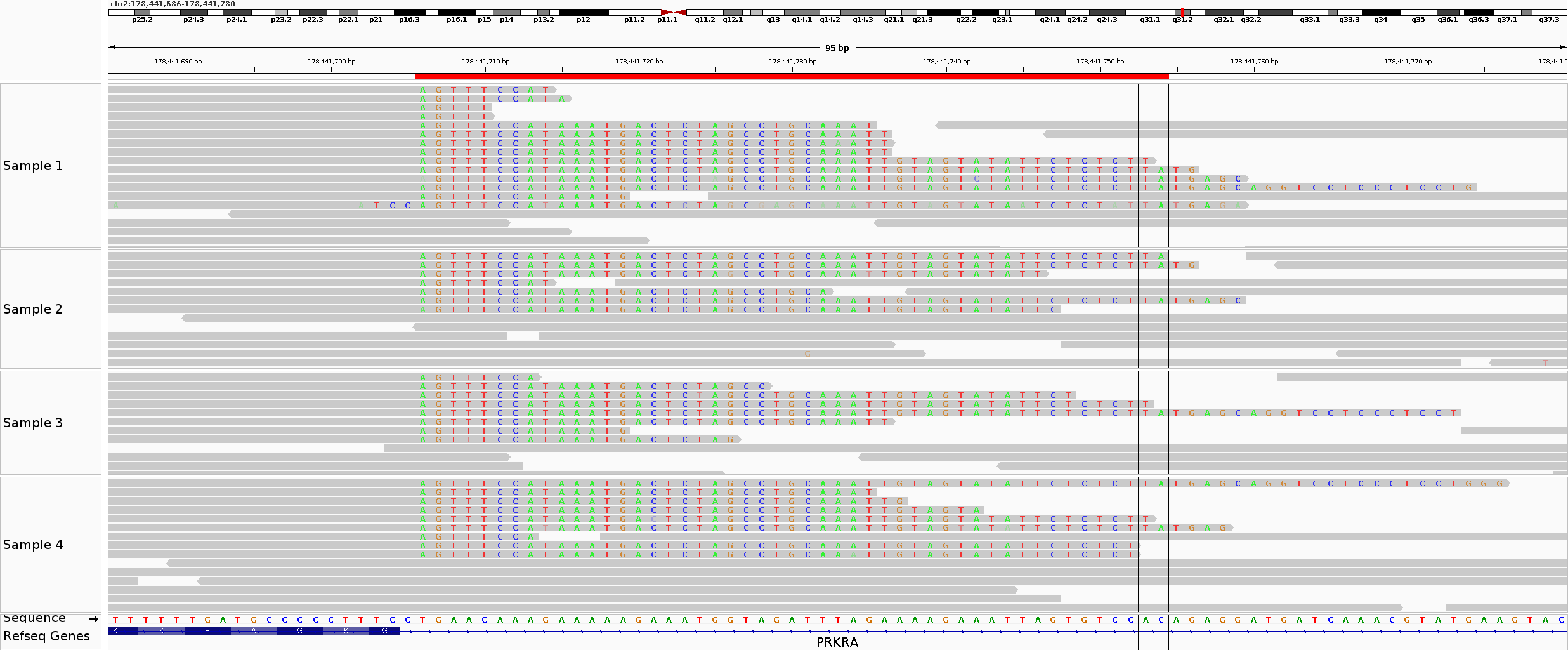

#### **Supplementary Fig. 3 IGV alignment view of two LoF variants (short insertion) in *PRKRA*.**

Insertions regions are bounded by solid blacklines and marked in light green (chr2:178441705:C>CAGTTTCCATAAATGACTCTAGCCTGCAATTGTAGTATATTCTCTCTTA) in samples 1,2 and light blue (chr2:178441705:C>CAGTTTCCATAAATGACTCTAGCCTGCAATTGTAGTATATTCTCTCT) in samples 3,4. The black boxes indicate soft clips, corresponding to mismatched bases at the ends of the reads. Aligned reads supported by soft-clipped bases which are different from reference bases indicates insertion regions

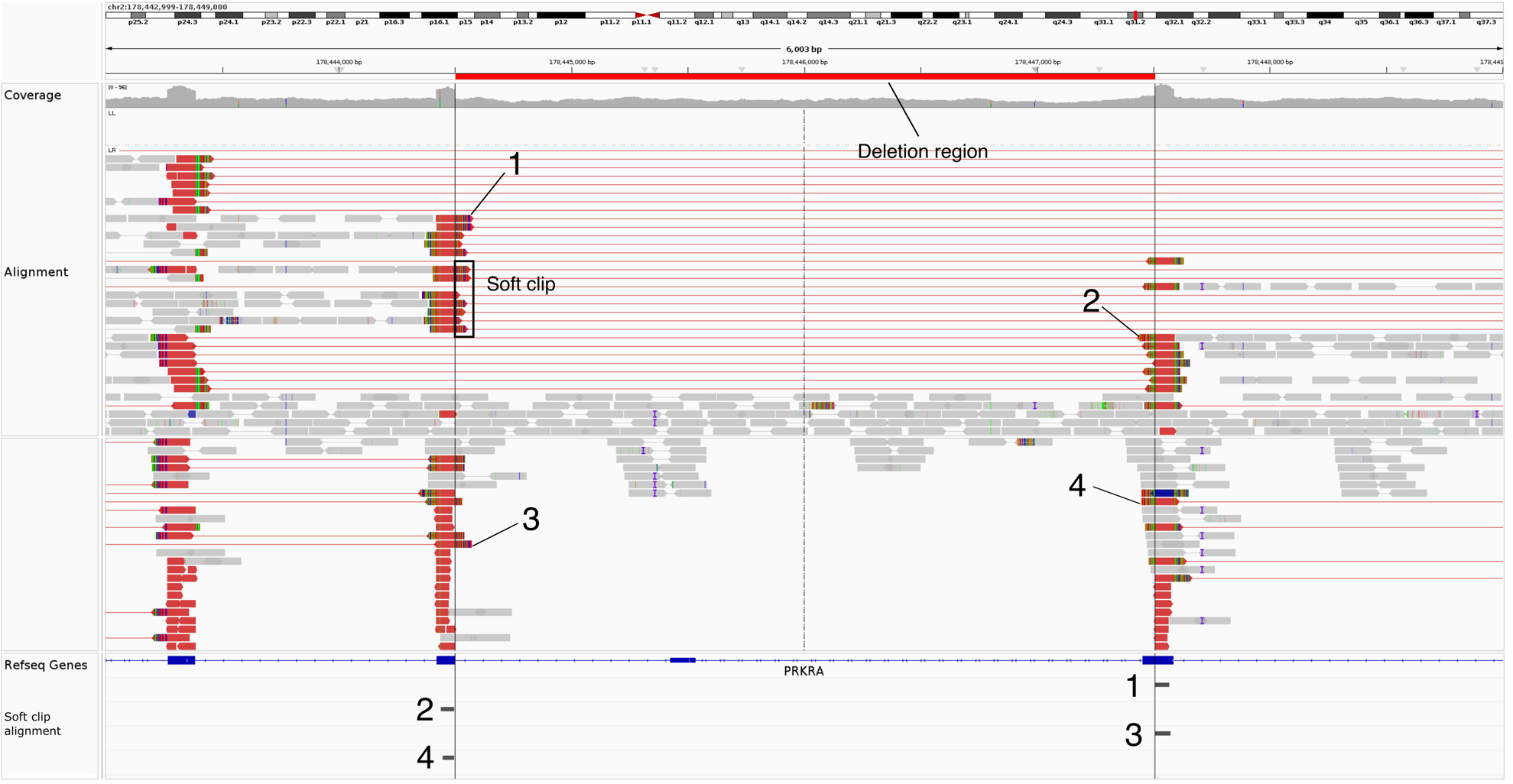

#### **Supplementary Fig. 4 IGV alignment view of the ~3kb deletion region in *PRKRA* in example sample 1.**

Deletion region is bounded by two solid black lines. Alignment is viewed in pairs, grouped by pair orientations, and coloured by insert size. Red or blue colour of the alignment indicates the insert size was larger or smaller, respectively, than the expected insert size. Numbers 1-4 denotes examples of soft-clipped bases matching to other regions that locate at the boundaries of the deletion region. The dotted black line represents the central axis of the view. The black boxes indicate soft clips, corresponding to mismatched bases at the ends of the reads.

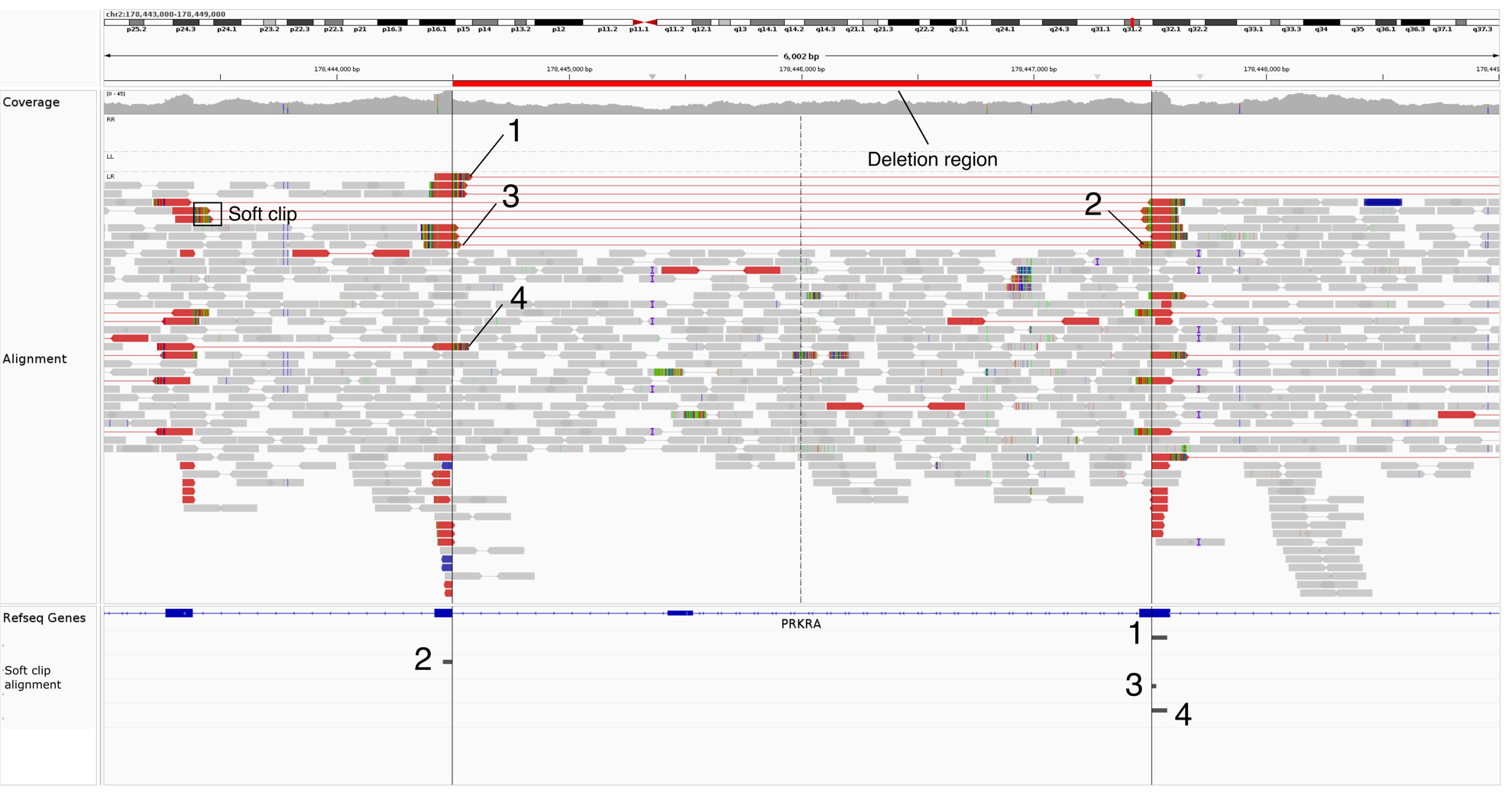

#### **Supplementary Fig. 5 Alignment view of the ~3kb deletion region in *PRKRA* in example sample 2.**

Deletion region is bounded by two solid black lines. Alignment is viewed in pairs, grouped by pair orientations, and coloured by insert size. Red or blue colour of the alignment indicates the insert size was larger or smaller, respectively, than the expected insert size. Numbers 1-4 denotes examples of soft-clipped bases matching to other regions that locate at the boundaries of the deletion region. The dotted black line represents the central axis of the view. The black boxes indicate soft clips, corresponding to mismatched bases at the ends of the reads.

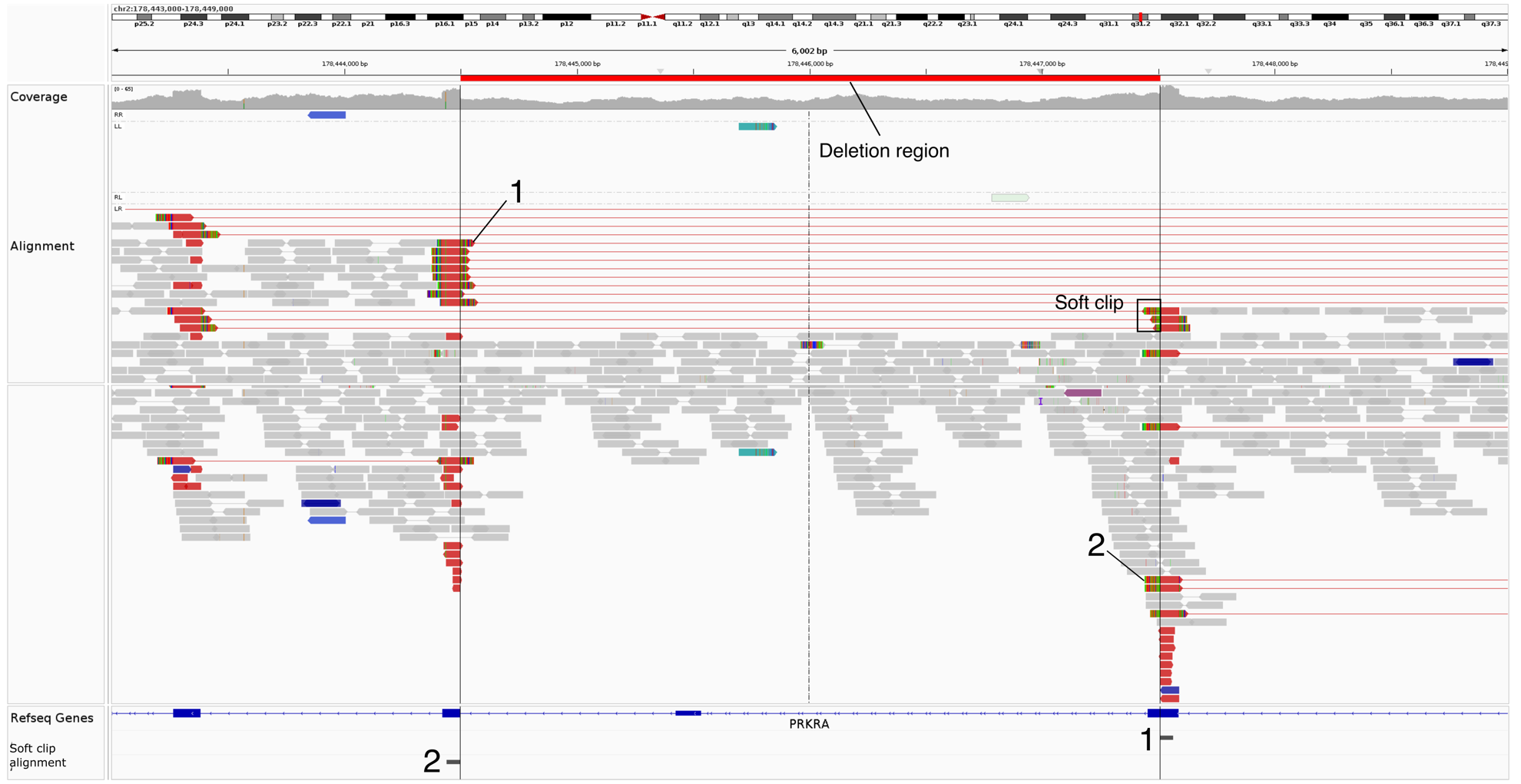

**Supplementary Fig. 6 Alignment view of the ~3kb deletion region in PRKRA in example sample 3.**

Deletion region is bounded by two solid black lines. Alignment is viewed in pairs, grouped by pair orientations, and coloured by insert size. Red or blue colour of the alignment indicates the insert size was larger or smaller, respectively, than the expected insert size. Numbers 1-4 denotes examples of soft-clipped bases matching to other regions that locate at the boundaries of the deletion region. The dotted black line represents the central axis of the view. The black boxes indicate soft clips, corresponding to mismatched bases at the ends of the reads.

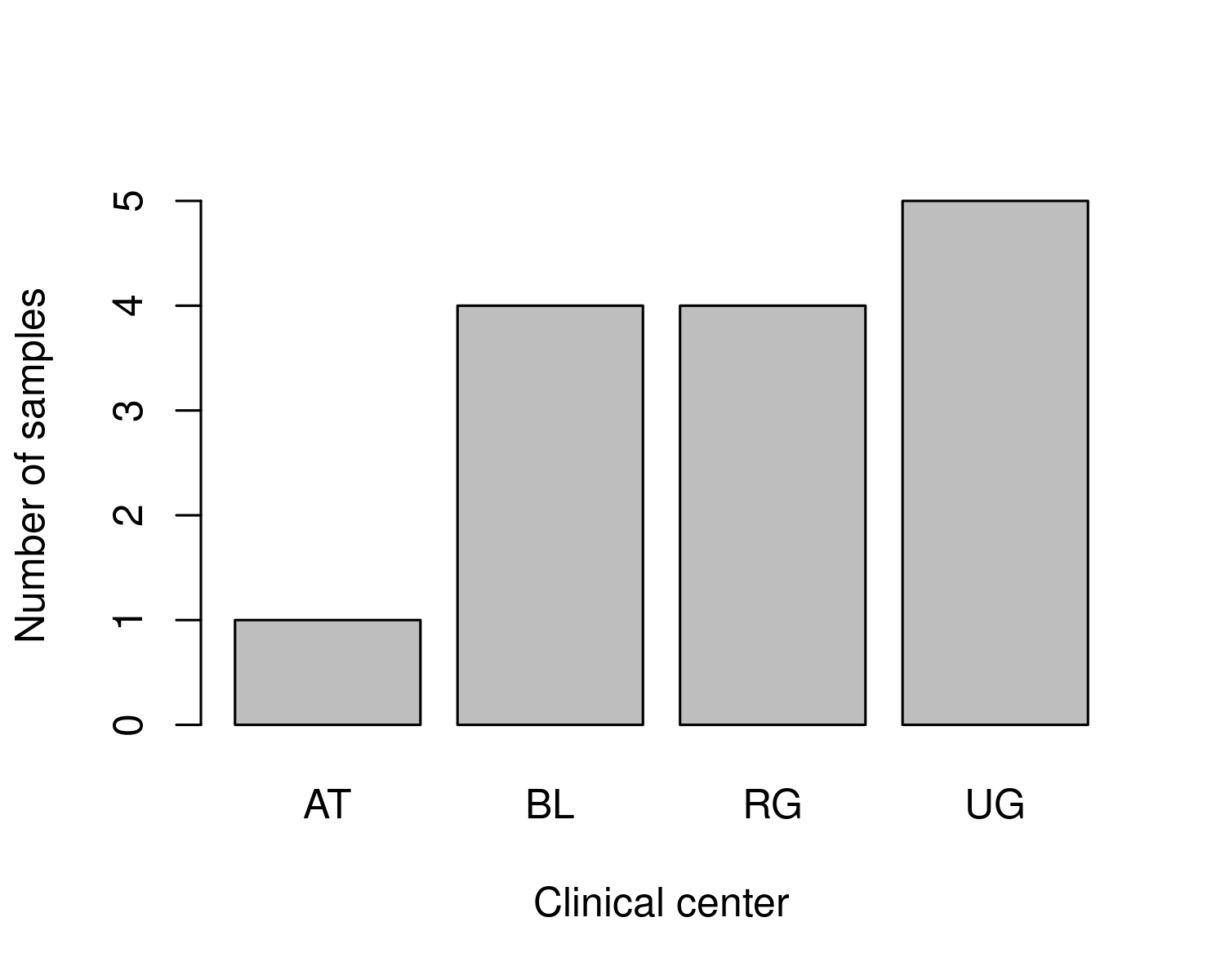

#### **Supplementary Fig. 7 Distribution of sample collection sites in carriers of deletion in *PRKRA*.**

Bar plot shows sample distribution across five clinical centers. AT, Athens; BL, Berlin; RG, Regensburg; UG, Granada.

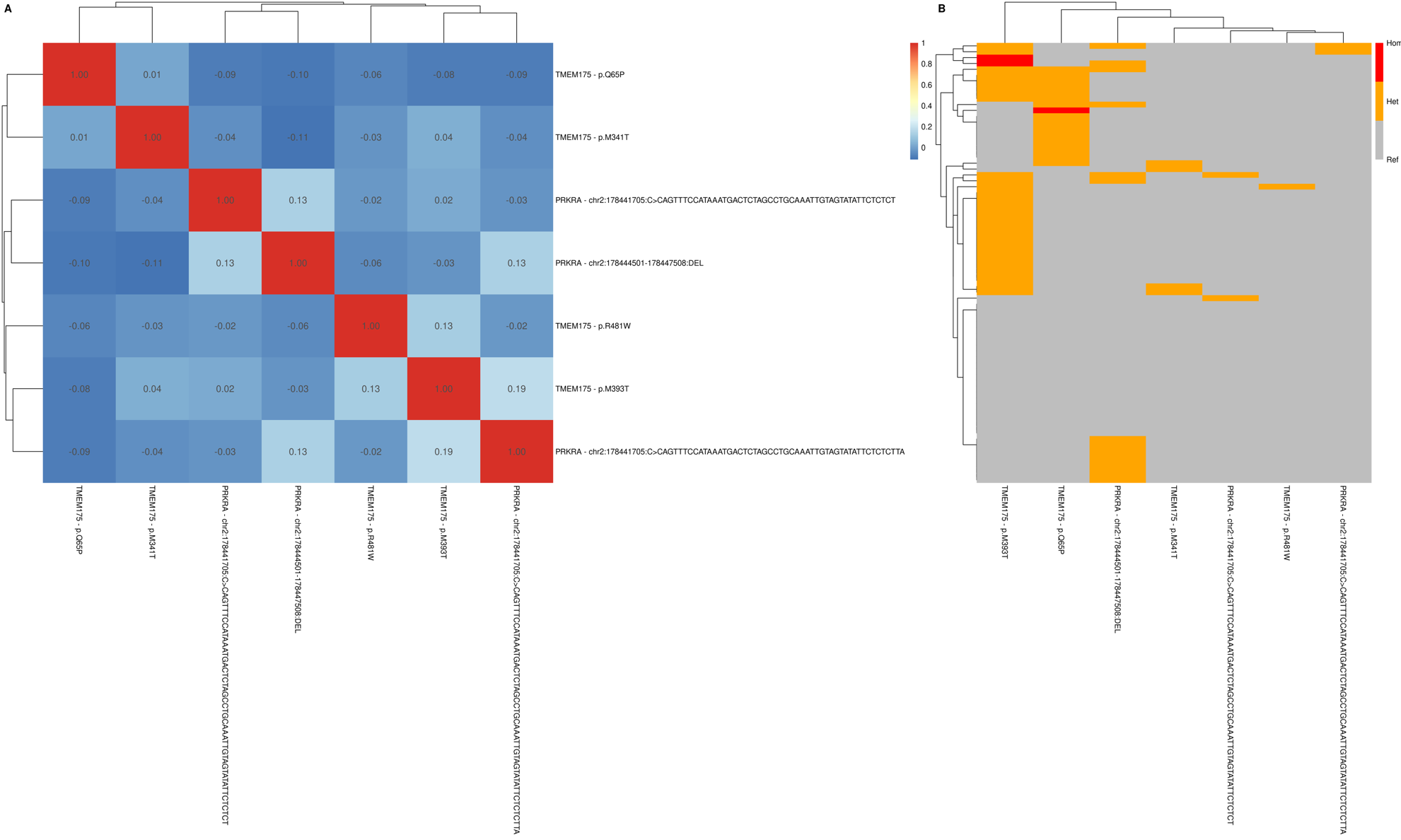

#### **Supplementary Fig. 8 Association between variants in *PRKRA* among MCI individuals and Parkinson’s disease-associated variants in *TMEM175*.**

Heatmap showing the Pearson’s correlation between variants in PRKRA and Parkinson’s disease risk variants in *TMEM175* (A) and co-occurrences of these variants in MCI individuals (B).

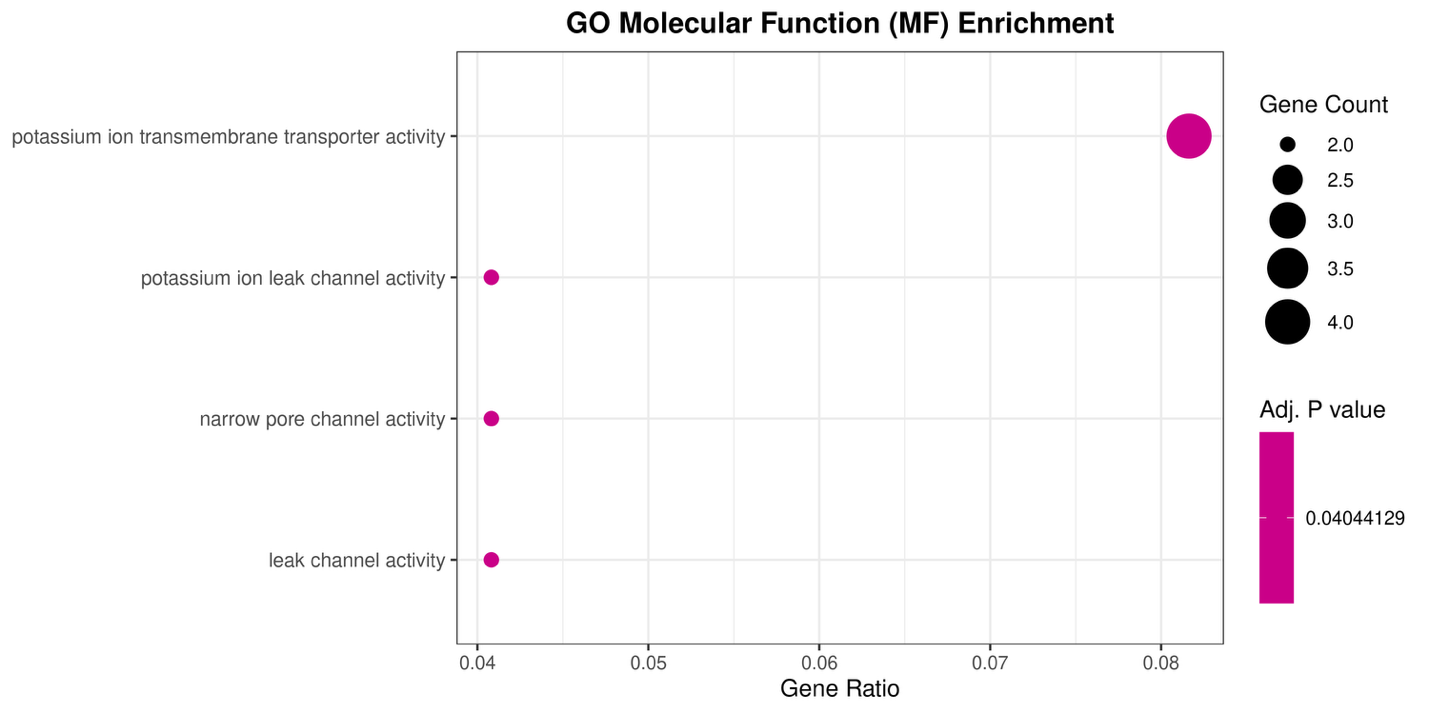
**Supplementary Fig. 9 Gene ontology (GO) enrichment of Molecular Function (MF) terms for the MCI gene burden set.**

Dot plot shows the molecular functions (y-axis) enriched among genes in the MCI gene burden set, with the corresponding gene ratio on the x-axis. Dot colour represents the adjusted *P* value (BH correction) and dot size indicates the number of genes associated with each MF.

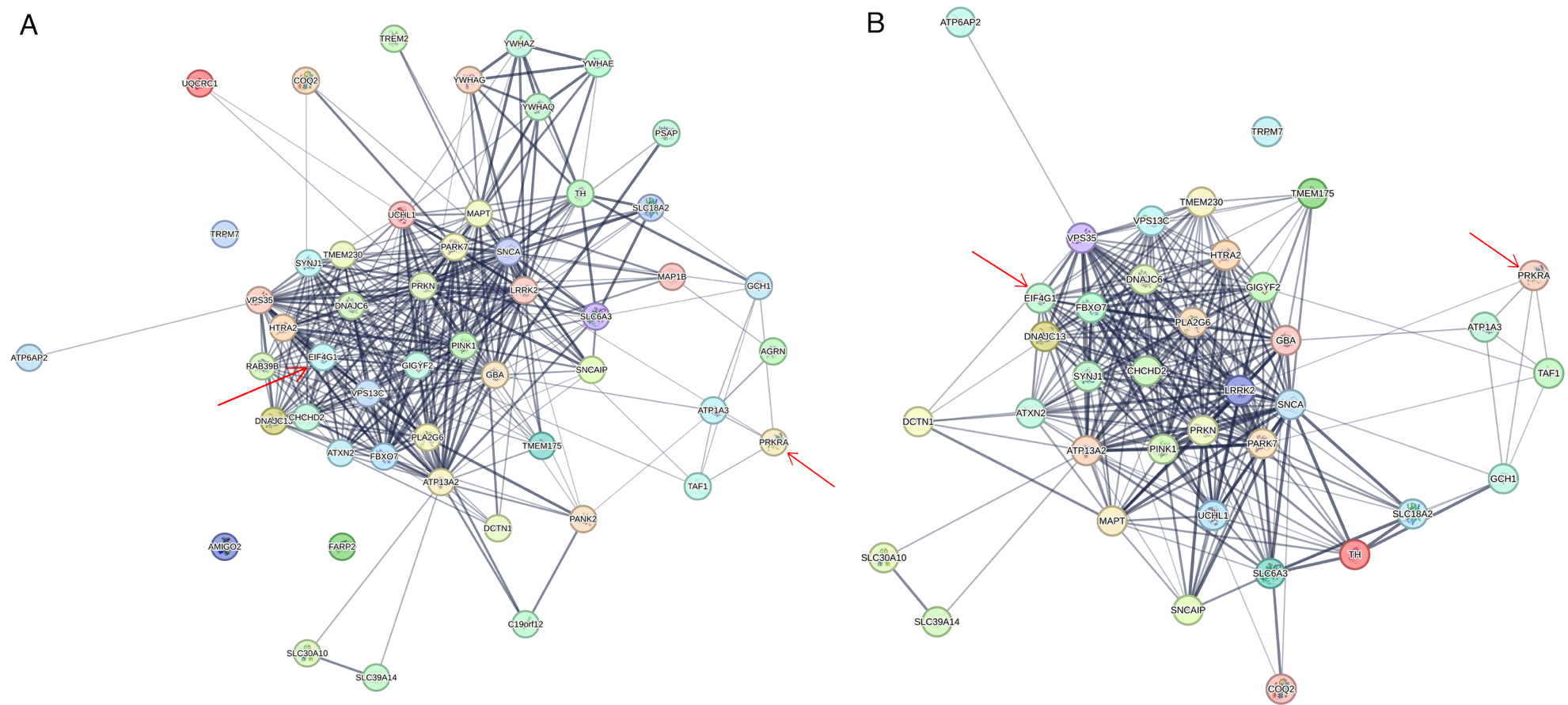

#### **Supplementary Fig. 10 Protein-protein association network of Parkinson’s disease-related genes.**

(A) Disease-gene associations from the Disease Ontology database (DOID: 14330).

(B) Protein annotations from UniProt (KW-0908).

The networks were constructed using seven evidence channels in STRING (text mining, neighbourhood, experiments, gene fusion, databases, co-occurrence, co-expression) and include *PRKRA* and *EIF4G1* (indicated by red arrows), which were identified in the MCI gene burden set. The thickness of network edges represents the strength of supporting evidence for each protein-protein association.

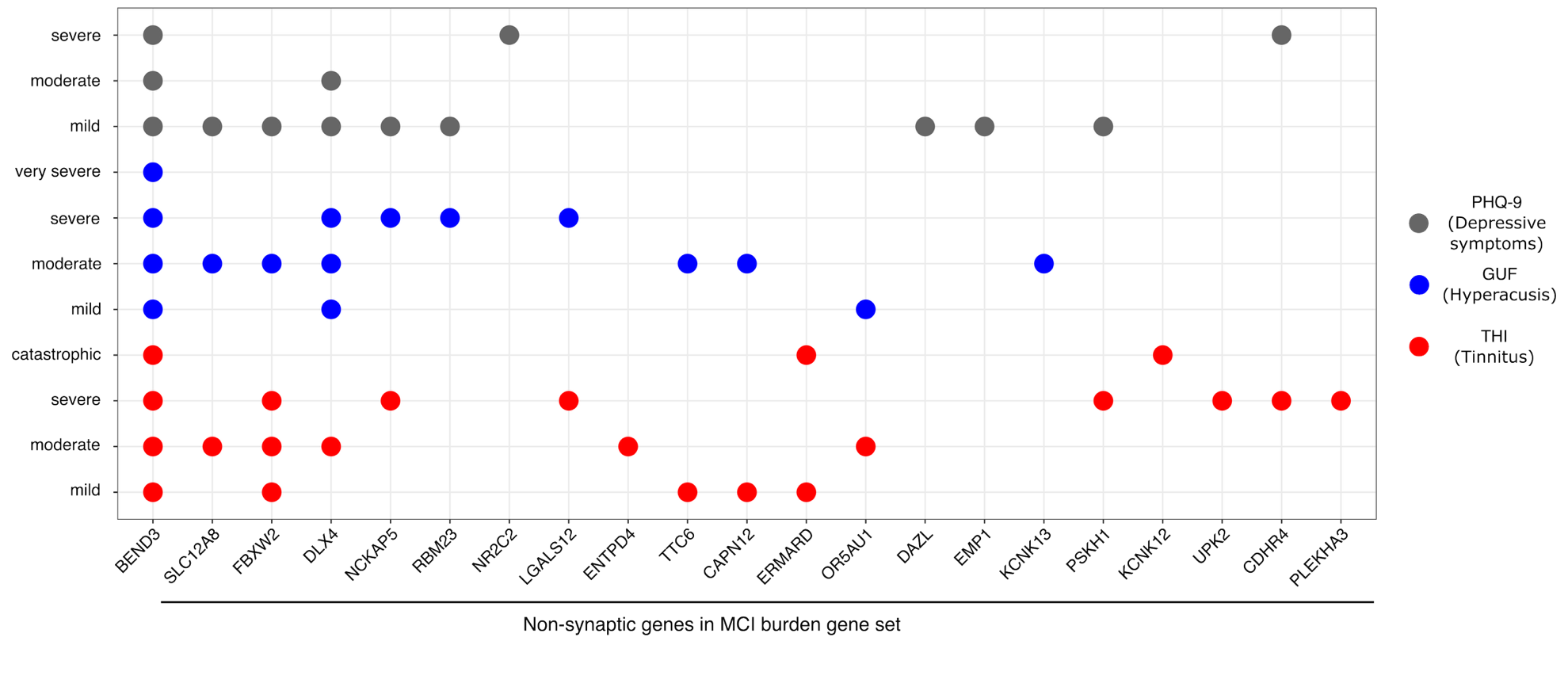

#### **Supplementary Fig. 11 Non-synaptic genes in the MCI gene burden set (x-axis) across severity levels of tinnitus, hyperacusis, and depressive symptoms (y-axis).**

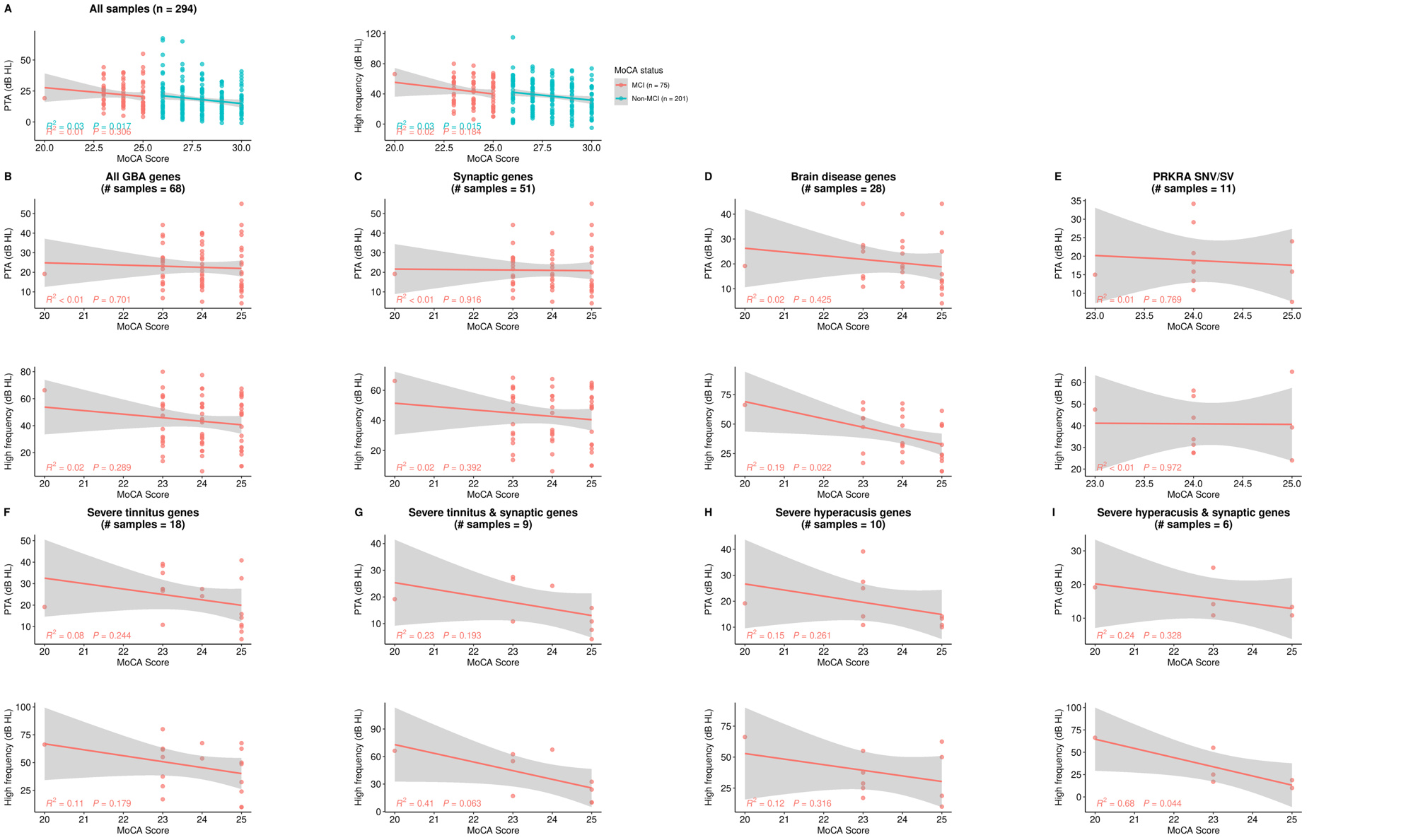

#### **Supplementary Fig. 12 Association between pure tone average (PTA) air-conducted hearing thresholds, high-frequency HL (HFHL), and MoCA score in representative gene sets.**

Scatterplot showing the correlation between MoCA score (x-axis) and PTA/HF (dB HL, y-axis) of patients in

**(A)** 294 UNITI individuals (75 MCI and 201 non-MCI);

**(B)** 68 MCI individuals carrying rare missense and loss-of-function variants in 52 burden genes;

**(C)** 51 MCI individuals carrying rare missense and loss-of-function variants in 21 synaptic burden genes;

**(D)** 28 MCI individuals carrying rare missense and loss-of-function variants in 12 burden genes associated with brain diseases;

**(E)** 11 MCI individuals carrying rare missense and loss-of-function variants (including structural variants) in *PRKRA* gene, associated with MCI and mild-to-moderate tinnitus;

**(F)** 20 MCI individuals carrying rare missense and loss-of-function variants in 3 synaptic genes (*GRAMD1A*, *ANKRD52*, *ENDOG*) and 8 non-synaptic genes (*NCKAP5*, *LGALS12*, *ERMARD*, *PSKH1*, *KCNK12*, *UPK2*, *CHDR4*, *PLEKHA3*) shared between MCI and severe/catastrophic tinnitus;

**(G)** 9 MCI individuals carrying rare missense and loss-of-function variants in 3 synaptic genes (*GRAMD1A*, *ANKRD52*, *ENDOG*) shared between MCI and severe-to-catastrophic tinnitus;

**(H)** 10 MCI individuals carrying rare missense and loss-of-function variants in 2 synaptic genes (*ANKRD52*, *SCN5A*) and 2 non-synaptic genes (*NCKAP5*, *RBM23*) shared between MCI and severe-to-very-severe hyperacusis;

**(I)** 6 MCI individuals carrying rare missense and loss-of-function variants in 2 synaptic genes (*ANKRD52*, *SCN5A*) shared between MCI and severe-to-very-severe hyperacusis. Sample size for regression plots are indicated in each panel title. GBA, gene burden analysis; MCI, mild cognitive impairment.

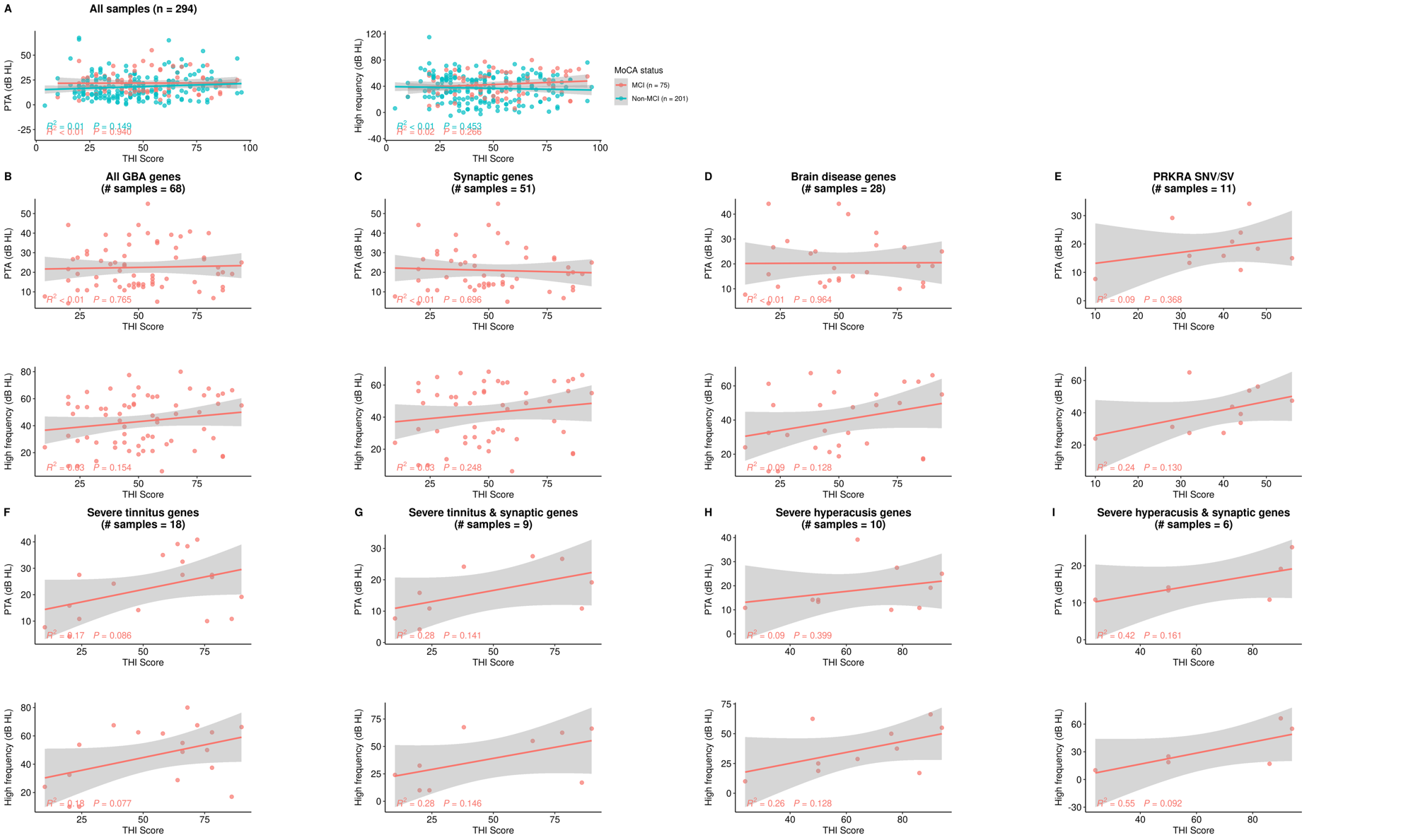

#### **Supplementary Fig. 13 Association between pure tone average (PTA) air-conducted hearing thresholds, high-frequency HL (HFHL), and THI score in representative gene sets.**

Scatterplot showing the correlation between THI score (x-axis) and PTA/HF (dB HL, y-axis) of patients in

**(A)** 294 UNITI individuals (75 MCI and 201 non-MCI);

**(B)** 68 MCI individuals carrying rare missense and loss-of-function variants in 52 burden genes;

**(C)** 51 MCI individuals carrying rare missense and loss-of-function variants in 21 synaptic burden genes;

**(D)** 28 MCI individuals carrying rare missense and loss-of-function variants in 12 burden genes associated with brain diseases;

**(E)** 11 MCI individuals carrying rare missense and loss-of-function variants (including structural variants) in *PRKRA* gene, associated with MCI and mild-to-moderate tinnitus;

**(F)** 20 MCI individuals carrying rare missense and loss-of-function variants in 3 synaptic genes (*GRAMD1A*, *ANKRD52*, *ENDOG*) and 8 non-synaptic genes (*NCKAP5*, *LGALS12*, *ERMARD*, *PSKH1*, *KCNK12*, *UPK2*, *CHDR4*, *PLEKHA3*) shared between MCI and severe/catastrophic tinnitus;

**(G)** 9 MCI individuals carrying rare missense and loss-of-function variants in 3 synaptic genes (*GRAMD1A*, *ANKRD52*, *ENDOG*) shared between MCI and severe-to-catastrophic tinnitus;

**(H)** 10 MCI individuals carrying rare missense and loss-of-function variants in 2 synaptic genes (*ANKRD52*, *SCN5A*) and 2 non-synaptic genes (*NCKAP5*, *RBM23*) shared between MCI and severe-to-very-severe hyperacusis;

**(I)** 6 MCI individuals carrying rare missense and loss-of-function variants in 2 synaptic genes (*ANKRD52*, *SCN5A*) shared between MCI and severe-to-very-severe hyperacusis. Sample size for regression plots are indicated in each panel title. GBA, gene burden analysis; MCI, mild cognitive impairment.

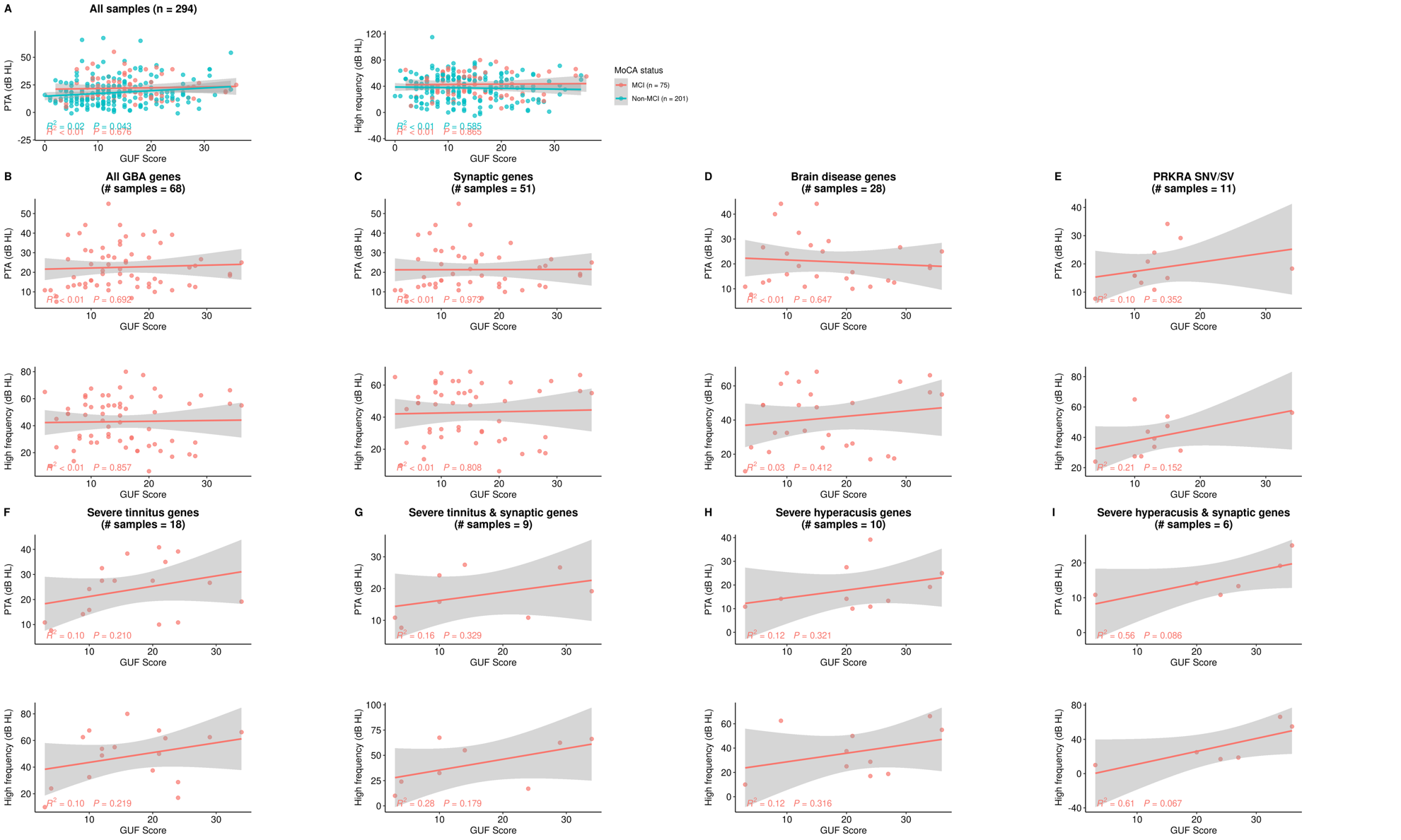

#### **Supplementary Fig. 14 Association between pure tone average (PTA) air-conducted hearing thresholds, high-frequency HL (HFHL), and GÜF score in representative gene sets.**

Scatterplot showing the correlation between GÜF score (x-axis) and PTA/HF (dB HL, y-axis) of patients in

**(A)** 294 UNITI individuals (75 MCI and 201 non-MCI);

**(B)** 68 MCI individuals carrying rare missense and loss-of-function variants in 52 burden genes;

**(C)** 51 MCI individuals carrying rare missense and loss-of-function variants in 21 synaptic burden genes;

**(D)** 28 MCI individuals carrying rare missense and loss-of-function variants in 12 burden genes associated with brain diseases;

**(E)** 11 MCI individuals carrying rare missense and loss-of-function variants (including structural variants) in *PRKRA* gene, associated with MCI and mild-to-moderate tinnitus;

**(F)** 20 MCI individuals carrying rare missense and loss-of-function variants in 3 synaptic genes (*GRAMD1A*, *ANKRD52*, *ENDOG*) and 8 non-synaptic genes (*NCKAP5*, *LGALS12*, *ERMARD*, *PSKH1*, *KCNK12*, *UPK2*, *CHDR4*, *PLEKHA3*) shared between MCI and severe/catastrophic tinnitus;

**(G)** 9 MCI individuals carrying rare missense and loss-of-function variants in 3 synaptic genes (*GRAMD1A*, *ANKRD52*, *ENDOG*) shared between MCI and severe-to-catastrophic tinnitus;

**(H)** 10 MCI individuals carrying rare missense and loss-of-function variants in 2 synaptic genes (*ANKRD52*, *SCN5A*) and 2 non-synaptic genes (*NCKAP5*, *RBM23*) shared between MCI and severe-to-very-severe hyperacusis;

**(I)** 6 MCI individuals carrying rare missense and loss-of-function variants in 2 synaptic genes (*ANKRD52*, *SCN5A*) shared between MCI and severe-to-very-severe hyperacusis. Sample size for regression plots are indicated in each panel title. GBA, gene burden analysis; MCI, mild cognitive impairment.

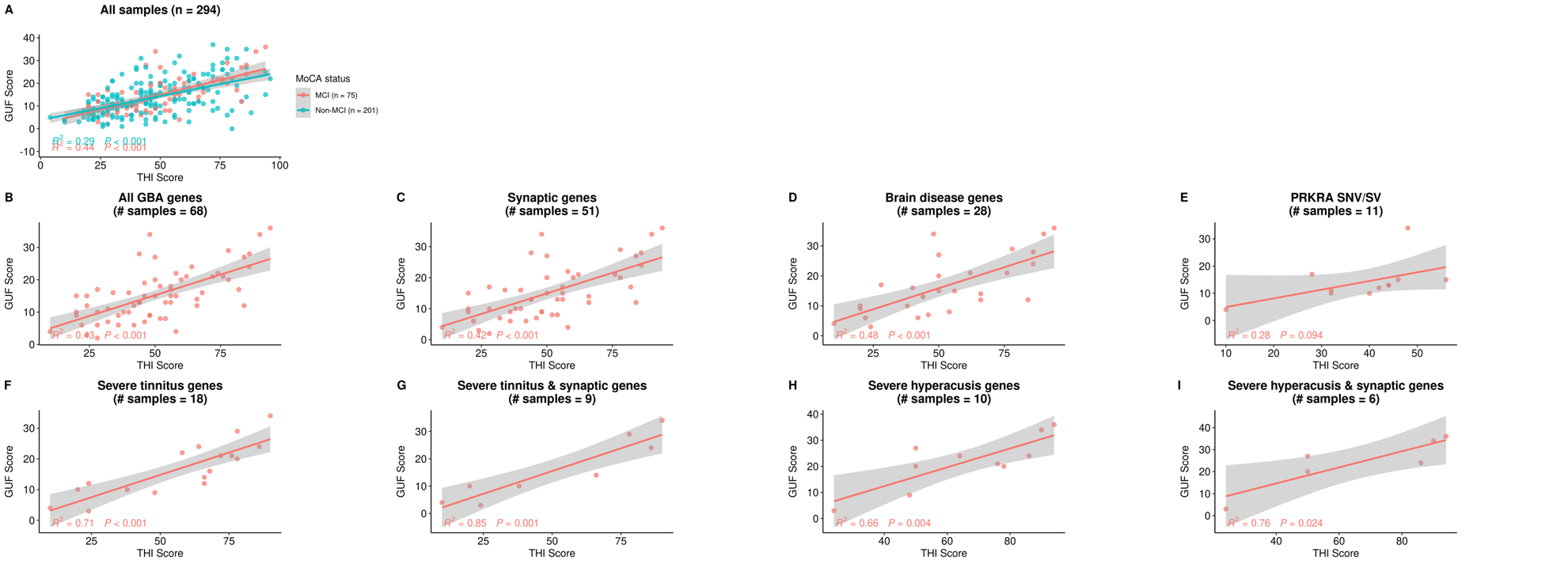

#### **Supplementary Fig. 15 Association between THI and GÜF scores in representative gene sets.**

Scatterplot showing the correlation between THI score (x-axis) and GÜF (y-axis) of patients in

**(A)** 294 UNITI individuals (75 MCI and 201 non-MCI);

**(B)** 68 MCI individuals carrying rare missense and loss-of-function variants in 52 burden genes;

**(C)** 51 MCI individuals carrying rare missense and loss-of-function variants in 21 synaptic burden genes;

**(D)** 28 MCI individuals carrying rare missense and loss-of-function variants in 12 burden genes associated with brain diseases;

**(E)** 11 MCI individuals carrying rare missense and loss-of-function variants (including structural variants) in *PRKRA* gene, associated with MCI and mild-to-moderate tinnitus;

**(F)** 20 MCI individuals carrying rare missense and loss-of-function variants in 3 synaptic genes (*GRAMD1A*, *ANKRD52*, *ENDOG*) and 8 non-synaptic genes (*NCKAP5*, *LGALS12*, *ERMARD*, *PSKH1*, *KCNK12*, *UPK2*, *CHDR4*, *PLEKHA3*) shared between MCI and severe/catastrophic tinnitus;

**(G)** 9 MCI individuals carrying rare missense and loss-of-function variants in 3 synaptic genes (*GRAMD1A*, *ANKRD52*, *ENDOG*) shared between MCI and severe-to-catastrophic tinnitus;

**(H)** 10 MCI individuals carrying rare missense and loss-of-function variants in 2 synaptic genes (*ANKRD52*, *SCN5A*) and 2 non-synaptic genes (*NCKAP5*, *RBM23*) shared between MCI and severe-to-very-severe hyperacusis;

**(I)** 6 MCI individuals carrying rare missense and loss-of-function variants in 2 synaptic genes (*ANKRD52*, *SCN5A*) shared between MCI and severe-to-very-severe hyperacusis. Sample size for regression plots are indicated in each panel title. GBA, gene burden analysis; MCI, mild cognitive impairment.

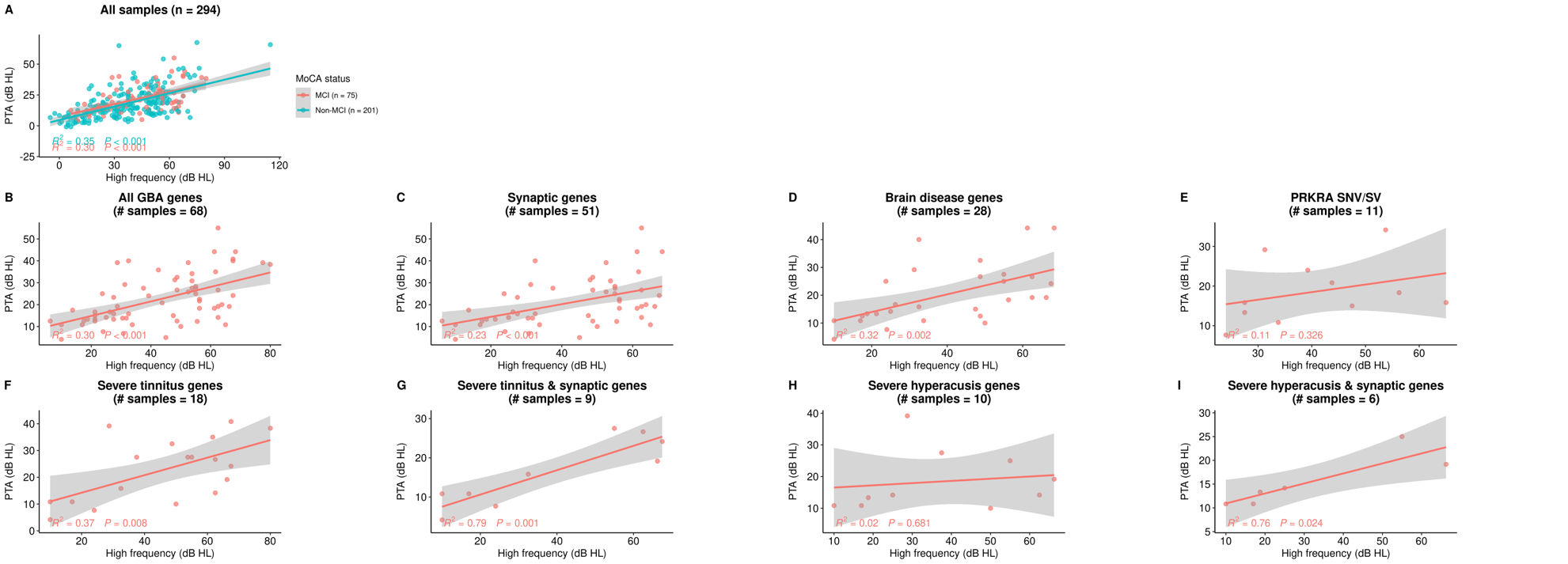

#### **Supplementary Fig. 16 Association between pure tone average (PTA) air-conducted hearing thresholds and high-frequency HL (HFHL) in representative gene sets.**

Scatterplot showing the correlation between PTA (dB HL, y-axis) and HF (dB HL, x-axis) of patients in

**(A)** 294 UNITI individuals (75 MCI and 201 non-MCI);

**(B)** 68 MCI individuals carrying rare missense and loss-of-function variants in 52 burden genes;

**(C)** 51 MCI individuals carrying rare missense and loss-of-function variants in 21 synaptic burden genes;

**(D)** 28 MCI individuals carrying rare missense and loss-of-function variants in 12 burden genes associated with brain diseases;

**(E)** 11 MCI individuals carrying rare missense and loss-of-function variants (including structural variants) in *PRKRA* gene, associated with MCI and mild-to-moderate tinnitus;

**(F)** 20 MCI individuals carrying rare missense and loss-of-function variants in 3 synaptic genes (*GRAMD1A*, *ANKRD52*, *ENDOG*) and 8 non-synaptic genes (*NCKAP5*, *LGALS12*, *ERMARD*, *PSKH1*, *KCNK12*, *UPK2*, *CHDR4*, *PLEKHA3*) shared between MCI and severe/catastrophic tinnitus;

**(G)** 9 MCI individuals carrying rare missense and loss-of-function variants in 3 synaptic genes (*GRAMD1A*, *ANKRD52*, *ENDOG*) shared between MCI and severe-to-catastrophic tinnitus;

**(H)** 10 MCI individuals carrying rare missense and loss-of-function variants in 2 synaptic genes (*ANKRD52*, *SCN5A*) and 2 non-synaptic genes (*NCKAP5*, *RBM23*) shared between MCI and severe-to-very-severe hyperacusis;

**(I)** 6 MCI individuals carrying rare missense and loss-of-function variants in 2 synaptic genes (*ANKRD52*, *SCN5A*) shared between MCI and severe-to-very-severe hyperacusis. Sample size for regression plots are indicated in each panel title. GBA, gene burden analysis; MCI, mild cognitive impairment.

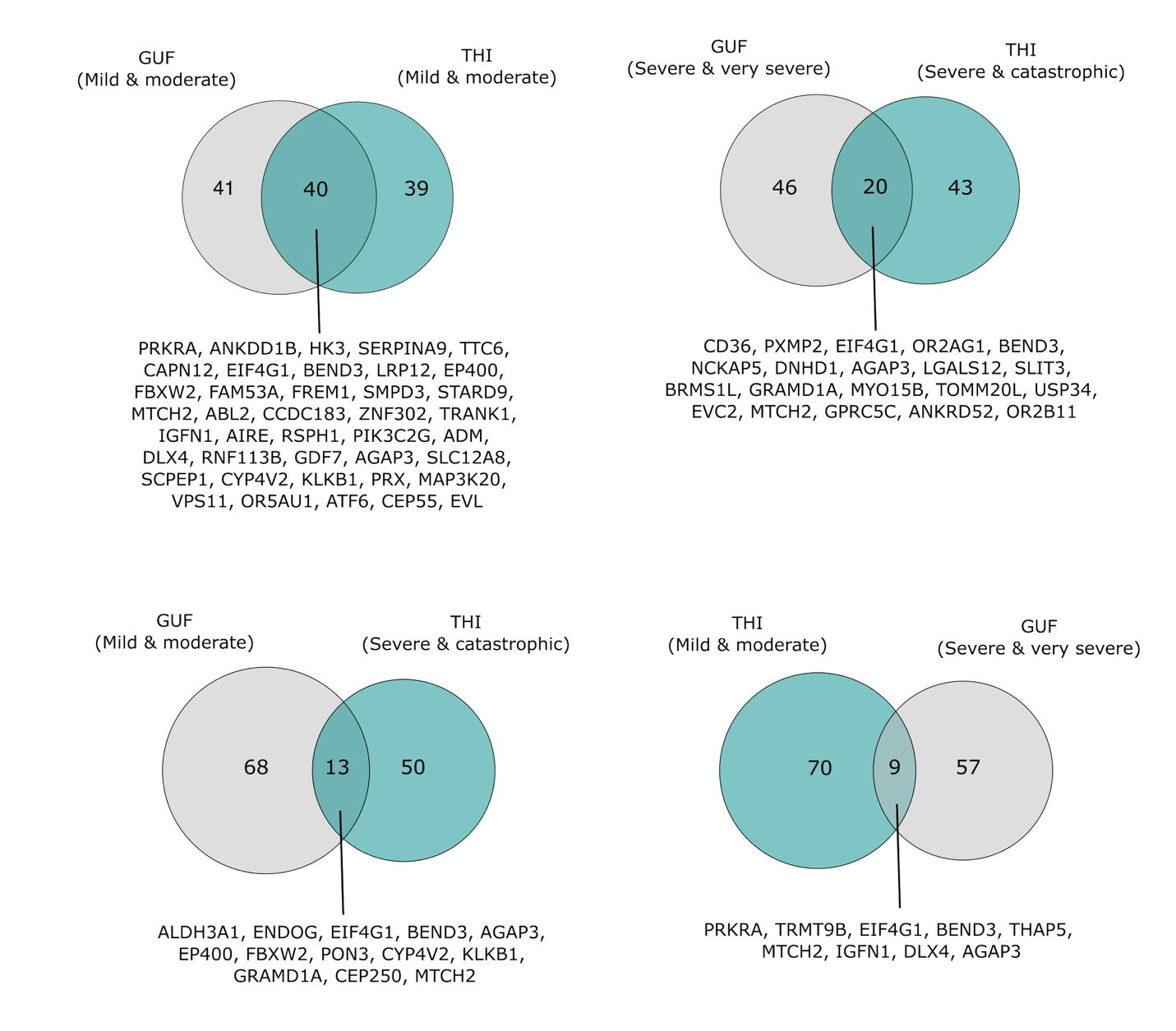

#### **Supplementary Fig. 17 Genes shared between tinnitus and hyperacusis according to GUF and THI scores severity.**

Venn diagrams show overlaps between mild-to-moderate tinnitus and mild-to-moderate hyperacusis (top-left), severe-to-very-severe hyperacusis and severe-to-catastrophic tinnitus (top-right), mild-to-moderate hyperacusis and severe-to-catastrophic tinnitus (bottom-left), severe-to-very-severe hyperacusis and mild-to-moderate tinnitus (bottom-right).

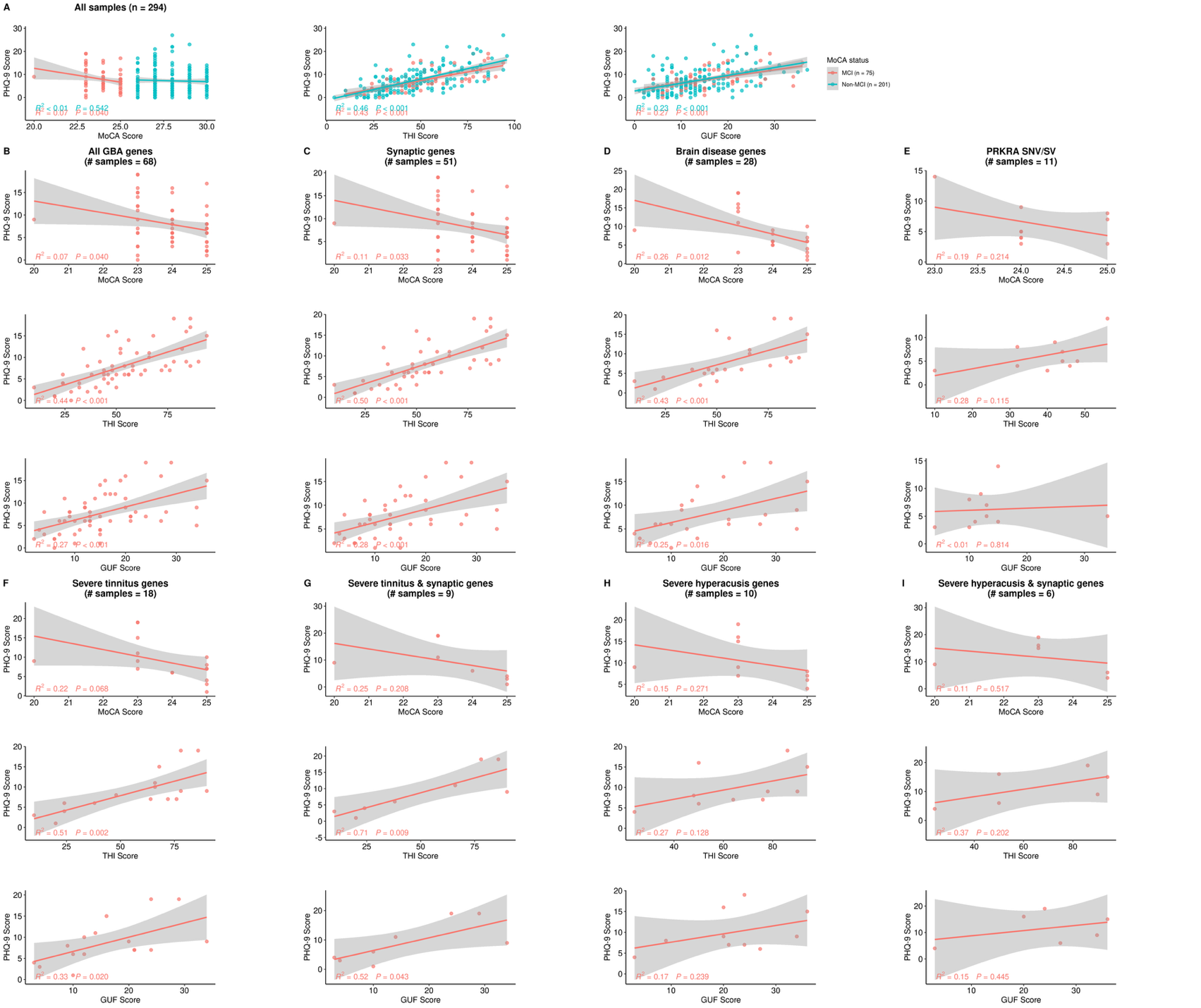

#### **Supplementary Fig. 18 Association between PHQ-9 score and MoCA/THI/GÜF scores in representative gene sets.**

Scatterplot showing the correlation between PHQ-9 score (y-axis) and MoCA/THI/GÜF (x-axis) of patients in

**(A)** 294 UNITI individuals (75 MCI and 201 non-MCI);

**(B)** 68 MCI individuals carrying rare missense and loss-of-function variants in 52 burden genes;

**(C)** 51 MCI individuals carrying rare missense and loss-of-function variants in 21 synaptic burden genes;

**(D)** 28 MCI individuals carrying rare missense and loss-of-function variants in 12 burden genes associated with brain diseases;

**(E)** 11 MCI individuals carrying rare missense and loss-of-function variants (including structural variants) in *PRKRA* gene, associated with MCI and mild-to-moderate tinnitus;

**(F)** 20 MCI individuals carrying rare missense and loss-of-function variants in 3 synaptic genes (*GRAMD1A*, *ANKRD52*, *ENDOG*) and 8 non-synaptic genes (*NCKAP5*, *LGALS12*, *ERMARD*, *PSKH1*, *KCNK12*, *UPK2*, *CHDR4*, *PLEKHA3*) shared between MCI and severe/catastrophic tinnitus;

**(G)** 9 MCI individuals carrying rare missense and loss-of-function variants in 3 synaptic genes (*GRAMD1A*, *ANKRD52*, *ENDOG*) shared between MCI and severe-to-catastrophic tinnitus;

**(H)** 10 MCI individuals carrying rare missense and loss-of-function variants in 2 synaptic genes (*ANKRD52*, *SCN5A*) and 2 non-synaptic genes (*NCKAP5*, *RBM23*) shared between MCI and severe-to-very-severe hyperacusis;

**(I)** 6 MCI individuals carrying rare missense and loss-of-function variants in 2 synaptic genes (*ANKRD52*, *SCN5A*) shared between MCI and severe-to-very-severe hyperacusis. Sample size for regression plots are indicated in each panel title. GBA, gene burden analysis; MCI, mild cognitive impairment.

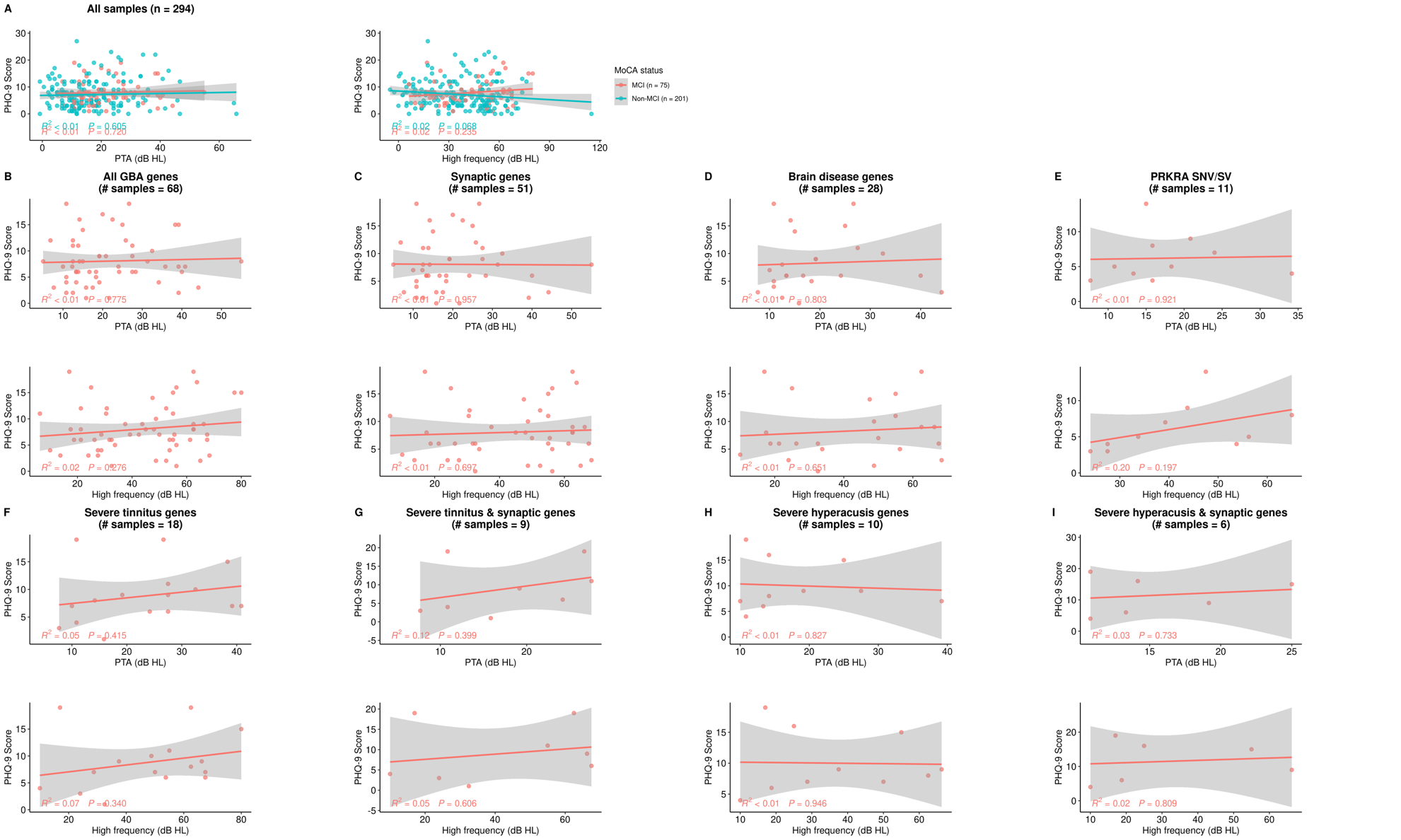

#### **Supplementary Fig. 19 Association between PHQ-9 score and PTA/High-frequency HL (HFHL) in representative gene sets.**

Scatterplot showing the correlation between PHQ-9 score (y-axis) and PTA/HF (db HL, x-axis) of patients in

**(A)** 294 UNITI individuals (75 MCI and 201 non-MCI);

**(B)** 68 MCI individuals carrying rare missense and loss-of-function variants in 52 burden genes;

**(C)** 51 MCI individuals carrying rare missense and loss-of-function variants in 21 synaptic burden genes;

**(D)** 28 MCI individuals carrying rare missense and loss-of-function variants in 12 burden genes associated with brain diseases;

**(E)** 11 MCI individuals carrying rare missense and loss-of-function variants (including structural variants) in *PRKRA* gene, associated with MCI and mild-to-moderate tinnitus;

**(F)** 20 MCI individuals carrying rare missense and loss-of-function variants in 3 synaptic genes (*GRAMD1A*, *ANKRD52*, *ENDOG*) and 8 non-synaptic genes (*NCKAP5*, *LGALS12*, *ERMARD*, *PSKH1*, *KCNK12*, *UPK2*, *CHDR4*, *PLEKHA3*) shared between MCI and severe/catastrophic tinnitus;

**(G)** 9 MCI individuals carrying rare missense and loss-of-function variants in 3 synaptic genes (*GRAMD1A*, *ANKRD52*, *ENDOG*) shared between MCI and severe-to-catastrophic tinnitus;

**(H)** 10 MCI individuals carrying rare missense and loss-of-function variants in 2 synaptic genes (*ANKRD52*, *SCN5A*) and 2 non-synaptic genes (*NCKAP5*, *RBM23*) shared between MCI and severe-to-very-severe hyperacusis;

**(I)** 6 MCI individuals carrying rare missense and loss-of-function variants in 2 synaptic genes (*ANKRD52*, *SCN5A*) shared between MCI and severe-to-very-severe hyperacusis. Sample size for regression plots are indicated in each panel title. GBA, gene burden analysis; MCI, mild cognitive impairment.

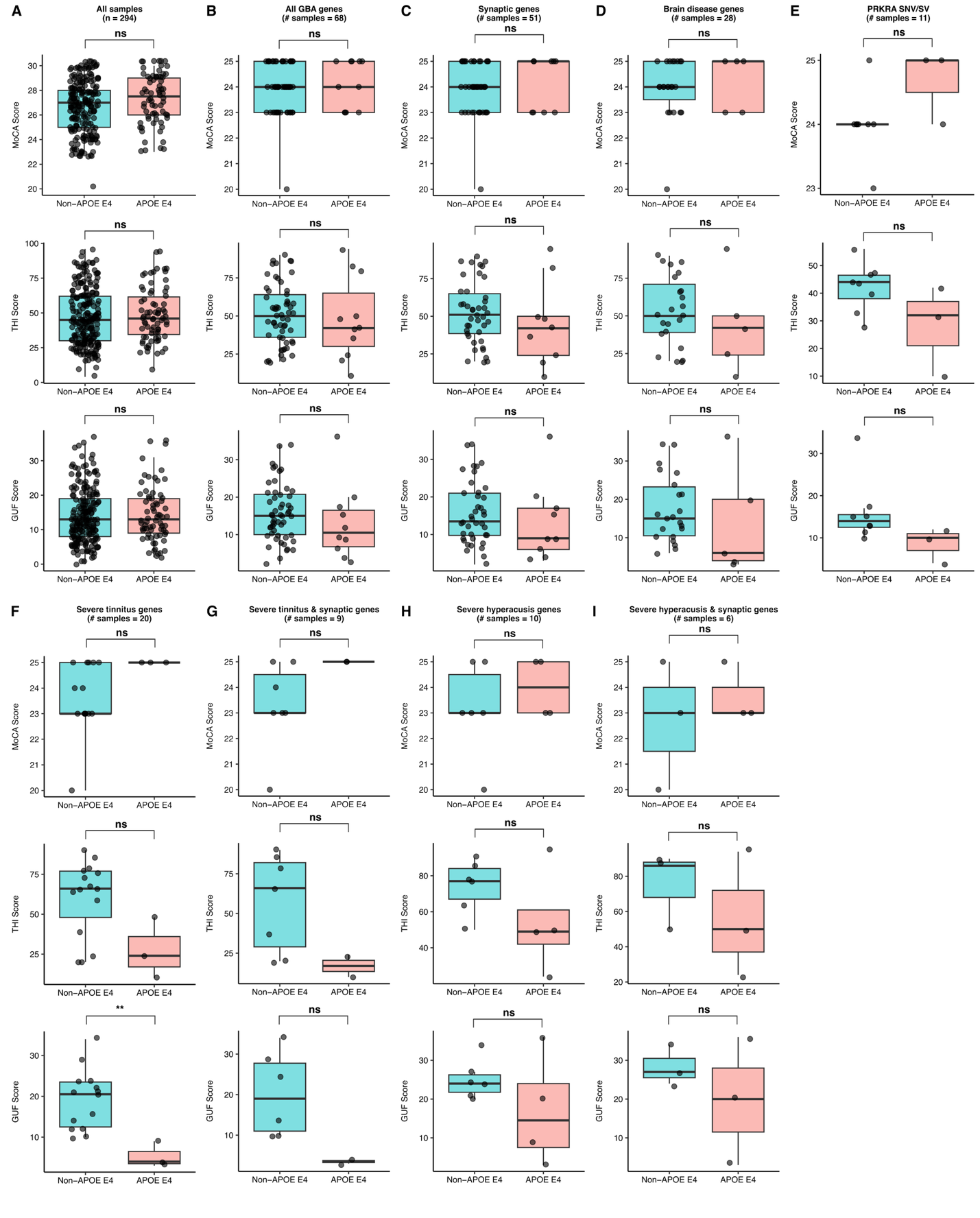

#### **Supplementary Fig. 20 Relationship between *APOE* ε4 genotypes and THI, GÜF, and MoCA score in representative gene sets.**

Boxplot showing the correlation between *APOE* ε4 genotypes and MoCA, THI/GÜF score of patients in

**(A)** 294 UNITI individuals;

**(B)** 68 MCI individuals carrying rare missense and loss-of-function variants in 52 burden genes;

**(C)** 51 MCI individuals carrying rare missense and loss-of-function variants in 21 synaptic burden genes;

**(D)** 28 MCI individuals carrying rare missense and loss-of-function variants in 12 burden genes associated with brain diseases;

**(E)** 11 MCI individuals carrying rare missense and loss-of-function variants (including structural variants) in *PRKRA* gene, associated with MCI and mild-to-moderate tinnitus;

**(F)** 20 MCI individuals carrying rare missense and loss-of-function variants in 3 synaptic genes (*GRAMD1A*, *ANKRD52*, *ENDOG*) and 8 non-synaptic genes (*NCKAP5*, *LGALS12*, *ERMARD*, *PSKH1*, *KCNK12*, *UPK2*, *CHDR4*, *PLEKHA3*) shared between MCI and severe/catastrophic tinnitus;

**(G)** 9 MCI individuals carrying rare missense and loss-of-function variants in 3 synaptic genes (*GRAMD1A*, *ANKRD52*, *ENDOG*) shared between MCI and severe-to-catastrophic tinnitus;

**(H)** 10 MCI individuals carrying rare missense and loss-of-function variants in 2 synaptic genes (*ANKRD52*, *SCN5A*) and 2 non-synaptic genes (*NCKAP5*, *RBM23*) shared between MCI and severe-to-very-severe hyperacusis;

**(I)** 6 MCI individuals carrying rare missense and loss-of-function variants in 2 synaptic genes (*ANKRD52*, *SCN5A*) shared between MCI and severe-to-very-severe hyperacusis. Sample size for each boxplot are indicated in each panel title. Statistical significance was assessed using a two-sided Wilcoxon test. ns, non-significant. GBA, gene burden analysis; MCI, mild cognitive impairment.

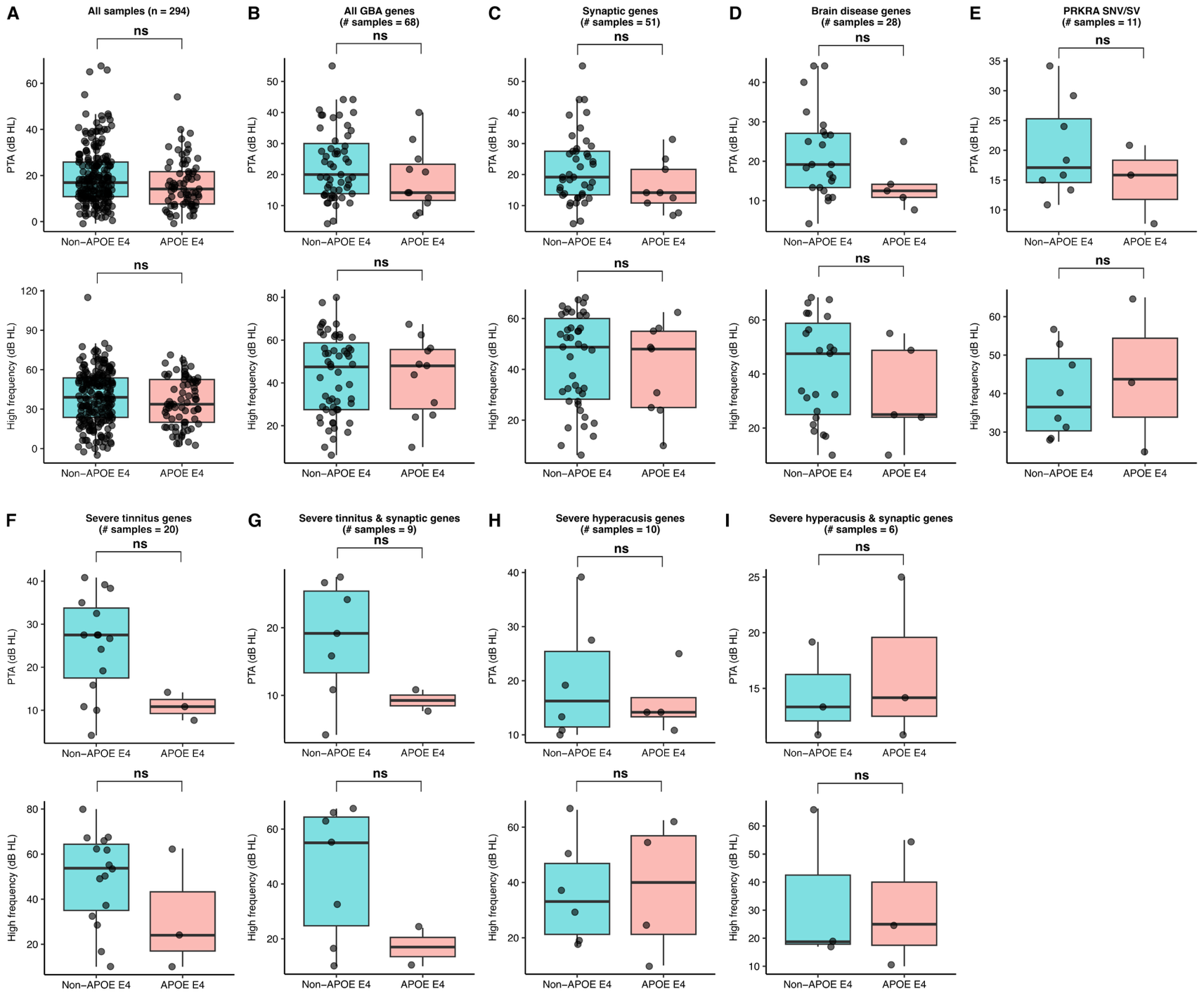

#### **Supplementary Fig. 21 Relationship between *APOE* ε4 genotypes and PTA/High-frequency HL in representative gene sets.**

Boxplot showing the correlation between *APOE* ε4 genotypes and PTA/High-frequency HL (dB) of patients in

**(A)** 294 UNITI individuals;

**(B)** 68 MCI individuals carrying rare missense and loss-of-function variants in 52 burden genes;

**(C)** 51 MCI individuals carrying rare missense and loss-of-function variants in 21 synaptic burden genes;

**(D)** 28 MCI individuals carrying rare missense and loss-of-function variants in 12 burden genes associated with brain diseases;

**(E)** 11 MCI individuals carrying rare missense and loss-of-function variants (including structural variants) in *PRKRA* gene, associated with MCI and mild-to-moderate tinnitus;

**(F)** 20 MCI individuals carrying rare missense and loss-of-function variants in 3 synaptic genes (*GRAMD1A*, *ANKRD52*, *ENDOG*) and 8 non-synaptic genes (*NCKAP5*, *LGALS12*, *ERMARD*, *PSKH1*, *KCNK12*, *UPK2*, *CHDR4*, *PLEKHA3*) shared between MCI and severe/catastrophic tinnitus;

**(G)** 9 MCI individuals carrying rare missense and loss-of-function variants in 3 synaptic genes (*GRAMD1A*, *ANKRD52*, *ENDOG*) shared between MCI and severe-to-catastrophic tinnitus;

**(H)** 10 MCI individuals carrying rare missense and loss-of-function variants in 2 synaptic genes (*ANKRD52*, *SCN5A*) and 2 non-synaptic genes (*NCKAP5*, *RBM23*) shared between MCI and severe-to-very-severe hyperacusis;

**(I)** 6 MCI individuals carrying rare missense and loss-of-function variants in 2 synaptic genes (*ANKRD52*, *SCN5A*) shared between MCI and severe-to-very-severe hyperacusis. Sample size for regression plots are indicated in each panel title. Statistical significance was assessed using a two-sided Wilcoxon test. ns, non-significant. GBA, gene burden analysis; MCI, mild cognitive impairment.

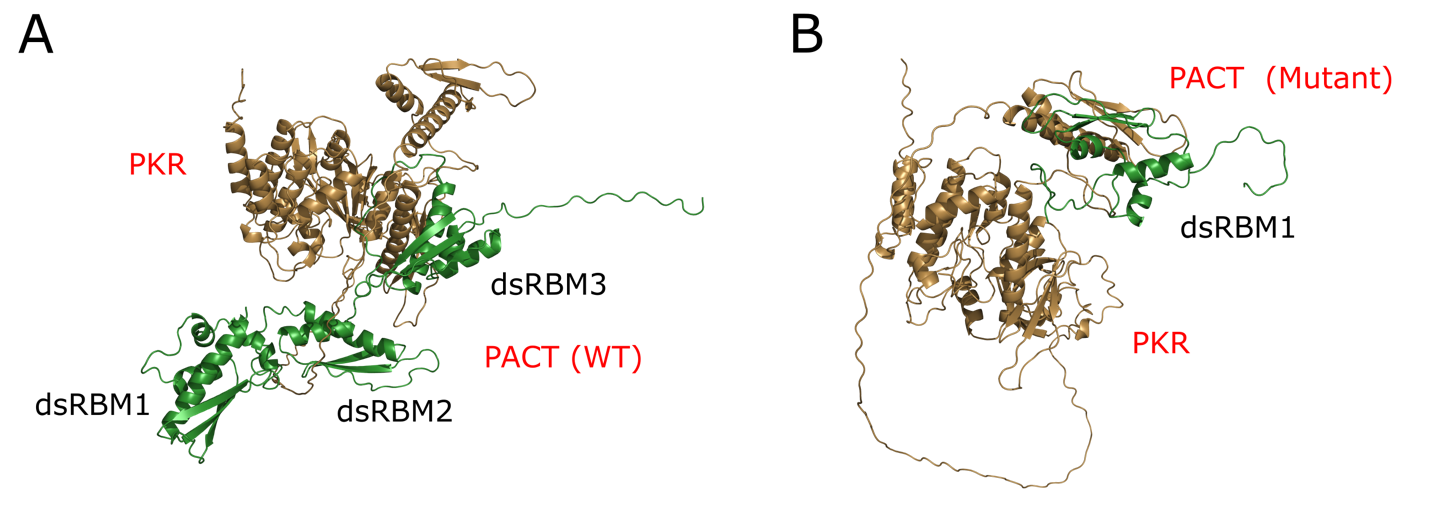

#### **Supplementary Fig. 22 Docking of wild-type (WT) and mutant PACT-PKR interaction**

WT PACT-PKR interaction (A) and mutant PACT-PKR interaction (B). Mutant PACT is a model of deletion at ILE-105. PACT (encoded by *PRKRA*) and PKR (encoded by *EIF2AK2*) are coloured in green and brown, respectively.
